# Suppression, delayed containment, or no intervention: optimal strategies for pandemic control

**DOI:** 10.1101/2024.12.04.24318460

**Authors:** Steinar Holden, Gunnar Rø, Ingrid Hjort

## Abstract

Future pandemics will require policymakers to balance reductions in infections and mortality against the societal costs of interventions. Using an integrated epidemiological–economic framework combining an age-structured SEIR model with endogenous behavioral responses, healthcare capacity constraints, Test–Trace–Isolate–Quarantine (TTIQ), and a unified measure of societal costs, we find that optimal intervention strategies cluster into three distinct regimes: *Suppression*, *No/negligible intervention*, or *Delayed containment*, where initial spread is followed by a short period of stringent interventions and subsequent control through TTIQ. By contrast, prolonged mitigation (*Flatten-the-curve*) strategies are optimal only in a narrow set of scenarios. The three-regime structure is robust across a broad parameter space of 4 320 scenarios, spanning transmissibility, disease severity, valuation of health losses, healthcare over-capacity costs, and vaccine timing. This pattern reflects non-convex trade-offs governing pandemic control: intermediate interventions often incur both substantial infection-related costs and substantial intervention costs. The framework provides a general approach for evaluating pandemic preparedness and intervention strategies.

## 1 Introduction

A novel respiratory pandemic confronts policymakers with a fundamental trade-off: reducing transmission through non-pharmaceutical interventions (NPIs) may substantially lower mortality and morbidity, but also imposes economic and welfare costs by restricting social interaction and economic activity. During the COVID-19 pandemic, countries adopted markedly different responses, ranging from rapid suppression to sustained mitigation or relatively limited intervention. Although a large literature has demonstrated that NPIs can reduce transmission [1–9], there remains little consensus on when strict interventions are socially justified and whether intermediate mitigation strategies, such as flattening-the-curve approaches, provide an effective compromise between health and societal costs [10–12].

COVID-19 has receded as an immediate policy concern, yet future pandemics remain a major global risk and preparedness continues to be a major public-health priority [13, 14]. Understanding how intervention strategies should depend on pathogen characteristics and societal conditions therefore remains an important challenge for pandemic preparedness.

One reason for the lack of consensus on intervention strategies is the inherent complexity of pandemic dynamics. Transmission evolves nonlinearly and depends on pathogen characteristics, country conditions (population density, contact and mobility patterns, baseline health), behavioral responses, and interventions. Moreover, the choice of intervention strategy also depends on how societies value heterogeneous consequences such as mortality and morbidity, broader welfare losses from NPIs, economic disruption, and distributional effects.

A growing epidemiological–economic (“epi–econ”) literature studies optimal pandemic interventions by integrating disease transmission models with economic costs and behavioral responses [15–27]. Broad assessments by [16, 27] find that suppression, mitigation or no/limited intervention may be optimal under different conditions. By contrast, [28, 29] show that it can be optimal initially to allow infections to spread and subsequently impose a sharp intervention rather than beginning with strong containment. [17] shows that contact tracing is much more efficient than general contact-reducing interventions when prevalence is low. Because results are sensitive to policy instruments and the specific epidemiological setting and parameters, and key studies advocate different strategies, it remains difficult to extract general guidance for intervention strategies in a future pandemic.

We develop an integrated epidemiological–economic framework to characterize optimal intervention strategies across a broad range of pandemic scenarios. The model combines an age-structured SEIR framework with endogenous voluntary social distancing, healthcare capacity constraints, Test–Trace–Isolate– Quarantine (TTIQ), and a unified measure of societal costs including monetized health losses, economic disruption, and welfare losses from reduced social interaction. While most previous studies compare a limited number of policy scenarios, our analysis focuses on general properties of the policy problem by systematically exploring a broad parameter space spanning transmissibility, severity, valuation of health losses, healthcare capacity costs, and vaccine timing. We also explore alternative behavioral responses. The broad and flexible approach encompasses several findings from earlier studies while also identifying novel solutions.

Our central finding is that optimal pandemic responses collapse into three distinct regimes: *Suppression*, *No/Negligible intervention*, and *Delayed containment*. The three-regime pattern reflects a structural non-convexity in the trade-offs governing pandemic control. Intermediate interventions often incur both substantial infection-related costs and substantial intervention costs, making them dominated by strategies that either suppress transmission sufficiently to keep incidence low or avoid costly interventions altogether. In our simulations, *Flatten-the-curve* strategies are optimal only in a narrow set of scenarios, primarily when healthcare overcapacity costs are very high and vaccination is substantially delayed.

This paper makes three contributions. First, we show that in a broad and flexible parameter scan, optimal intervention strategies collapse into three distinct regimes, *Suppression*, *No/Negligible intervention* and *Delayed containment*. The three-regime finding is consistent with [16, 27], but one of their regimes, mitigation, is replaced by a delayed containment strategy drawing upon [28, 29]. Second, we identify a strategy, *Delayed containment*, which combines delayed interventions found in [28–31] with a testing-and-isolation-policy (TTIQ) studied by [17]. The novel feature lies in the combination. Initial spread increases population immunity and thereby reduces the reproduction potential of the pathogen, after which a stringent intervention reduces incidence. For a pathogen with transmissibility too high to be controlled by TTIQ alone, greater population immunity reduces the effective reproduction number sufficiently for TTIQ to maintain control after a stringent intervention has brought incidence to a low level. Delayed containment can therefore keep cumulative incidence below the herd-immunity threshold that would prevail in the absence of interventions, at much lower cost than continued general contact-reducing interventions. Our parameter scan demonstrates the quantitative relevance within a broad comparison of alternative strategies.

Third, we provide a flexible epi–econ framework integrating behavioral responses and healthcare constraints that can be used to evaluate pandemic preparedness and intervention policies under alternative pathogen and societal conditions. We show that it is crucial to take the extent and type of voluntary social distancing into account when deciding on government interventions. We also show how the framework can be used to quantify the value of specific preparedness measures and how their benefits depend on the overall intervention strategy.

Our analysis highlights features that can complicate pandemic policymaking. The interaction between epidemic dynamics, interventions, and individual behavior, alongside disparate consequences for health, welfare, and economic activity, may generate substantial disagreement about appropriate policies. Such disagreement can encourage intermediate compromise policies. However, our results suggest that intermediate policies may be costly if they combine substantial intervention costs with substantial health losses.

## 2 Materials and methods

We use a deterministic metapopulation SEIR-type model with nine age groups. Infections progress through latent, presymptomatic, asymptomatic or symptomatic infectious states and may result in hospital admission, intensive-care treatment, recovery or death, with age-specific transition rates and probabilities calibrated to COVID-19 in Norway. Transmission between age groups is governed by an age-specific contact matrix. The complete system of compartments, differential equations and parameter values is reported in the S1 Appendix. The model belongs to a family of metapopulation models used for COVID-19 analysis at the Norwegian Institute of Public Health (NIPH) [32, 33], similar to models used for other countries, see eg. [34, 35]. An earlier version of the model was used by an expert group giving advice to the Norwegian authorities [36].

We extend the model to include contact reduction from government interventions and individuals’ voluntary social distancing (VSD), as well as valuation of the health, economic and welfare consequences. The evolution of the pandemic depends on the initial state and overall transmissibility of the disease, as well as government interventions and VSD, measured by indexes on the unit interval: *G*(*t*) *∈* [0, 1] measures government interventions and *V ^i^*(*t*) *∈* [0, 1] measures VSD per age group *i*. Individuals decide *V ^i^*(*t*) by weighing the welfare loss from social distancing against the expected health loss that can be avoided, see further description below. Figure 1 provides a conceptual overview of the model.

**Figure 1:**
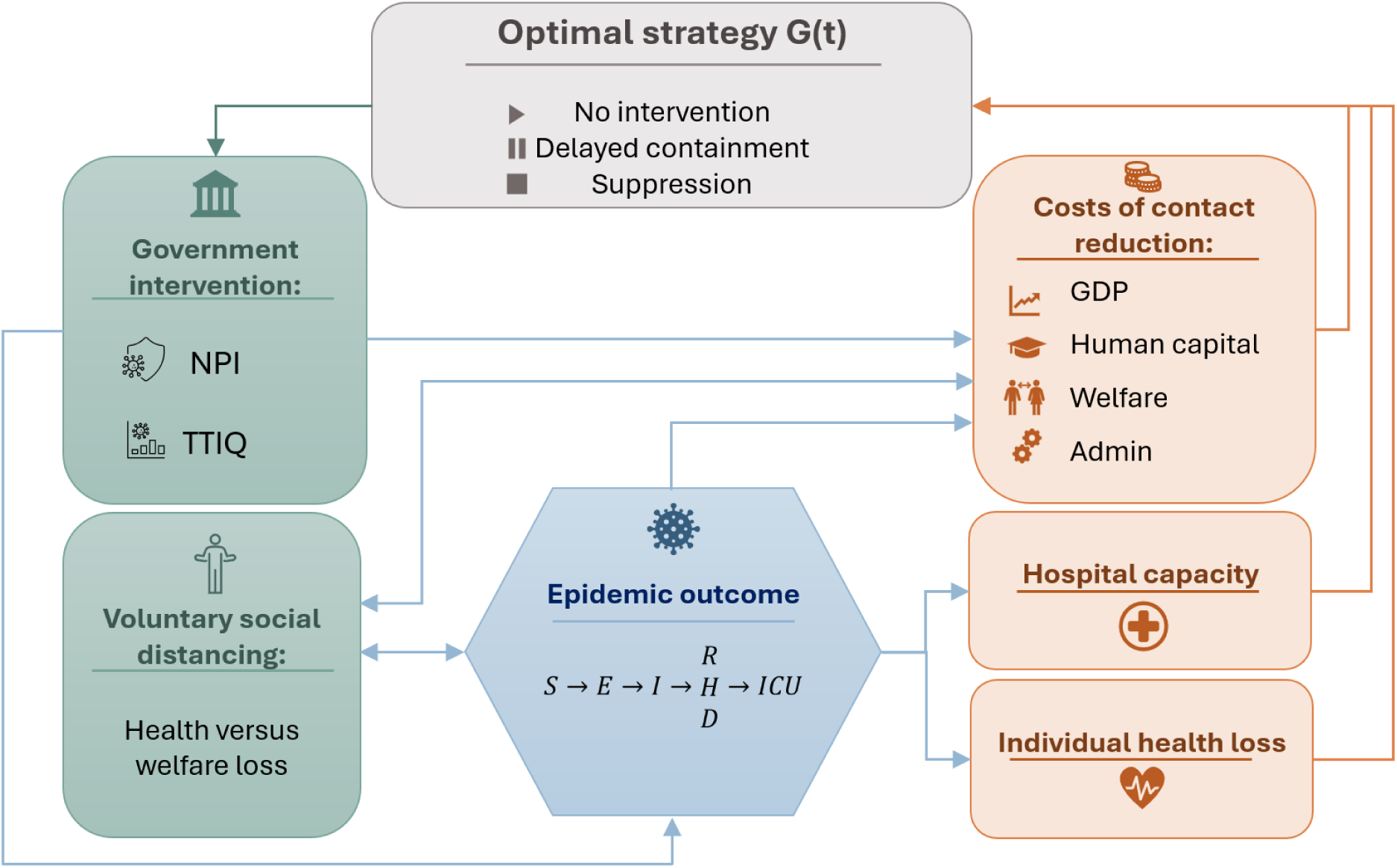
Conceptual overview of the integrated epi-econ framework. The framework links interventions and voluntary behavior to epidemic outcomes and societal costs, with policy chosen to minimize total cost.

For a given path of government interventions over time, *G*(*t*), and all model parameters included in a vector *θ*, the model, *M*, provides both the complete disease state, *S*(*t*) and the amount of voluntary social distancing, *V ^i^*(*t*):

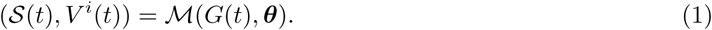

The optimal intervention strategy is the path of possibly time-varying level of interventions, *G*(*t*), that minimizes the total societal costs of the pandemic and interventions.

The policymaker chooses the time path of interventions *G*(*t*) to minimize total societal costs over the duration of the pandemic. The optimization problem is non-linear and non-convex because interventions affect epidemic dynamics through multiple interacting channels.

The baseline calibration of intervention and health costs is informed by the experiences during the COVID-19 pandemic in Norway. However, the objective of the analysis is not to evaluate a single historical episode, but to characterize general properties of optimal pandemic responses across a broad range of epidemiological and societal conditions.

### 2.1 Interventions and individual responses in a pandemic

The spread of the virus is affected by various types of government interventions like Test-Trace-Isolate-Quarantine (TTIQ) and other non-pharmaceutical interventions (NPIs) aimed at general contact reduction, as well as voluntary social distancing (VSD), where individuals choose to limit their social interactions to lower the risk of infection for themselves and others [5, 21–23, 37–39]. [32] found that the lockdown in Norway in March/April 2020 led to a substantial and statistically significant reduction in the reproduction number *R*, with a point estimate of 85% and regional estimates varying from 69% to 93%. [40] report suggestive evidence that transmission was reduced by 85 % in Denmark in the spring 2020. We assume that the maximum feasible reduction in the contact rate, from combined NPIs, TTIQ and VSD, is a reduction in *R* by 90%.

Government interventions and voluntary distancing will have partially overlapping effects. For example, if individuals avoid stores to reduce the risk of being infected, there will be little additional effect from a legal restriction closing stores. VSD can also complement government interventions when private responses are heterogeneous or social norms and coordination failures leave some high-contact behavior uncorrected. Many studies seek to empirically distinguish the effects of government interventions from those of VSD [5, 38, 41, 42]. Findings vary widely, and in a review of the literature, [43] find that the effects attributed to lockdown measures range from 12% to 60%.

We assume that the overall reduction in the contact rate 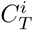 for age group *i* from interventions *G* and voluntary distancing *V ^i^*is:

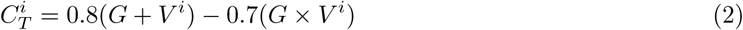

The coefficients of equation 2 are chosen so that either channel alone can reduce contacts by up to 80%, while combined interventions and VSD can reduce contacts by up to 90%, implying substantial but incomplete overlap. We assume that the effect of *G* and *V ^i^* on the contact rate is constant over time, see further discussion below.

The index for government interventions *G* should be thought of as a generic intervention stringency index. We interpret the first part of the index as TTIQ, which is cost-effective relative to more stringent lockdown measures when infection rates are low [17, 44]. Specifically, we interpret *G* in [0, 0.25] as a system of TTIQ that reduces effective transmission through testing, contact tracing, isolation, and quarantine, which in the model is represented as up to a 20% reduction in effective contacts if *V ^i^* = 0. [4] finds a small but statistically significant effect on *R* across OECD countries, and [45] conclude based on UK data that well-implemented contact tracing can provide up to a 15% reduction in *R*. Stronger effects of TTIQ are found by [46] for Britain and [44] for Australia.

Further interventions, for *G ∈* [0.25, 1], are assumed to be generic contact-reducing NPIs like workplace closures, school guidelines, cancellation of public events, public transport restrictions and restrictions on gathering sizes that lower the virus’s effective reproduction rate.

### 2.2 Voluntary social distancing (VSD)

Various approaches to VSD have been used in the epi-econ literature, from empirically based relationships based on mobility measures [16] or the death rate from the virus [4], to assumptions of full information rational expectations [27]. We assume that individuals across age groups weigh their welfare costs of staying at home over a given period against their expected health costs from infection risk associated with normal social activity. As individuals take into account their own health risk, there is much stronger social distancing among older age groups, consistent with evidence during the pandemic, see eg. [47] and [48].

The average welfare cost of voluntarily staying at home is set at 285 NOK per day (around 28 USD), based on a Swedish choice experiment conducted in mid-April 2020 [49]. As the experiment shows considerable individual variation, costs are assumed to be uniformly distributed across individuals from 0 to 570 NOK per day (see *SI Appendix*, section C for details). The expected health costs from continuing with normal social activity are calculated on the basis of the contemporaneous probability of infection, multiplied by age-specific QALY-loss from infection. This could be interpreted as a reduced-form proxy for publicly perceived infection risk, which in practice is likely to be formed from reported cases, hospitalizations, personal experience, and public-health communication. We use a logistic transformation to ensure that the individual’s choice responds gradually to changes in the relative utility of social distancing. This transformation captures that individuals may abstain from actions involving high risk, yet they may take some risk in other situations.

The VSD parameters are calibrated to match a reduction in peak incidence and hospital admissions of approximately 50% and 75%, respectively, in a SARS-CoV-2-like epidemic (*ES* = 15) without government intervention. There is considerable uncertainty; the literature documents substantial VSD, but quantitative evidence varies and the effect may also depend on the specific country and situation [5, 21–23, 37–39]. For robustness, we also consider no VSD as well as extended VSD where individuals also take into account the adverse health effects of their behavior on others (altruism).

The VSD reduces the contact-rate at the start of the simulations, so the initial reproduction numbers are defined conditional on this behavioral response, cf. *SI Appendix*, A.5.

### 2.3 The cost of interventions

The calibration of intervention costs is based on observed GDP losses during the interventions the winter of 2020–21. These interventions were as strict as those imposed in March 2020 [50], but adaptation, learning and fiscal support reduced the negative impact on the economy [51]. Measured by the GDP gap, the reduction was equal to 4.6% of mainland GDP, which corresponds to 12.7 bnNOK (billion Norwegian kroner) per month. Including the tax-financing costs (i.e. the efficiency loss from increased taxes due to higher government expenditure), estimated by [36] at 2 bnNOK per month, the total economic cost amounts to about 15 bnNOK per month.

Contact-reducing measures also result in welfare losses, which are calibrated using the choice experiment in [49], ensuring consistency between individuals’ and the decision maker’s evaluation of welfare costs and health costs. Assuming an average welfare loss of 285 NOK per person per day for the maximum reduction in the contact rate of 90%, yields about 47 bnNOK per month for Norway’s population of 5.4 mn. The total intervention costs at maximum intervention, both economic and welfare, are 62 bnNOK per month, about 22% of monthly mainland GDP in 2021, the base year for our calculations.

To make the intervention costs a continuous function of interventions and social distancing, we define an auxiliary quadratic function:

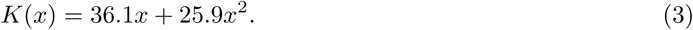

Note that *K*(1) = 62, equal to the costs from maximum contact reduction, while *K*(0.5) = 24.5, implying a convex cost function where 50% of the maximal reduction in the contact rate costs 40% of the total costs for the maximum reduction (as 24.5*/*62 *≈* 0.4).

The overall cost from contact reduction for age group *i* depends on government interventions *G* and the age-specific VSD *V ^i^*, applying the same overlap structure used for transmission:

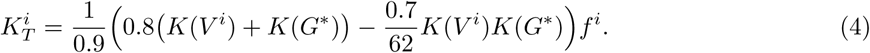

*G^∗^* = max(0, (*G −* 0.25)*/*0.75) is a measure of ordinary NPIs, as the costs of TTIQ, *G* in [0, 0.25], is calculated separately. Furthermore, *f^i^* is the fraction of the population in age group *i*. The total cost from contact reduction measured in bnNOK per month is then given by:

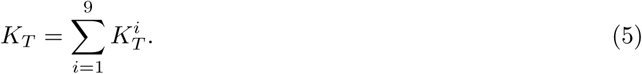

We assume that each change in intervention level *ΔG* entails a cost, up to a maximum cost of 3 bnNOK for large changes in *G*. These additional costs represent information, decision and implementation costs associated with changing interventions.

While the effect of TTIQ is modeled as a reduction in effective transmission as explained above, the costs of TTIQ are calculated separately based on the assumed number of detected cases. The costs are then calibrated based on the funding for Norwegian municipalities of 3 bnNOK in 2021 as extraordinary payments, earmarked as discretionary funds for infection control and the TTIQ-strategy. These costs are added to the cost of contact reduction. We assume half (1.5 bnNOK) covers initial setup (1/3) and daily fixed costs (2/3), while the rest corresponds to administrative expenses slightly below 3000 NOK per infected. In addition, there are economic and welfare effects for individuals in isolation or quarantine. The costs are assumed to be proportional to the extent of TTIQ, implying that for eg. *G* = 0.125, only half of these costs are included.

### 2.4 The cost of the disease

The disease may also involve large health losses in terms of symptomatic infections, hospitalizations, treatment in the Intensive Care Unit (ICU), post acute sequelae (long COVID-type symptoms) and deaths. Health losses are calibrated to the estimates of loss of QALYs from COVID-19 in [36], with a value of 1.4 mnNOK per QALY, consistent with Norwegian cost-benefit guidance for the Value of a Statistical Life Year (VSLY) in 2021 [52]. This corresponds to about 140 000 USD and is roughly five times per capita household consumption in Norway in 2021. For comparison, [53] and [54] set VSLY in the US to 6.25 and 6 times annual per capita consumption, respectively.

We assume a 20% mortality rate for patients with COVID-19 in the ICU at normal capacity, based on [55]. Based on [56], mortality increases linearly with overload, so that each additional percentage point of occupancy above capacity increases ICU mortality by 0.7% of its baseline level, capped at a maximum of 75% mortality. This increase applies uniformly to COVID-19 and non-COVID-19 patients due to triage considerations. The average death in the ICU is assumed to involve a loss of 10.4 QALYs, based on the assumption that ICU mortality follows a similar exponential increase with age as overall COVID-19 mortality in Norway, weighted by the age distribution of those who have received ICU treatment for COVID-19 in Norway, using public data from [57]. To summarize: Each admission to the ICU when it is operating above capacity is associated with an additional loss of 10.4(0.2 *∗* 0.7%*X*) QALY (up to 75% mortality), where *X* denotes the ICU occupancy above capacity, measured in percentage points.

### 2.5 Vaccine and immunity

Based on the rapid development of vaccines during the COVID-19 pandemic, we assume that a vaccine will also be developed for a new pathogen. In the baseline case, an effective vaccine becomes available after nine months, but we also consider longer times to vaccination, up to 1500 days. We capture the vaccination in a simplistic way by imposing *R*_eff_ = 0.7 after vaccination. We include a cool-down period of 150 days after vaccination, to capture the full health cost from infections before vaccination.

In line with most of the literature, we assume that individuals who have been infected by the virus will have immunity to re-infection for the time span until vaccination. Furthermore, we assume that the virus cannot be eliminated through strict interventions, including TTIQ. For countries with considerable cross-border trade and mobility, it would be difficult to adopt an elimination strategy without cooperation from neighboring countries. However, if elimination of the virus is feasible, it is likely to be a good strategy if the disease burden is high, as argued by [58] and analyzed by [16, 17].

For simplicity, we assume that there is no improvement in medical treatment before the arrival of the vaccine.

### 2.6 Optimal Strategy

The epi-econ literature typically explores the optimal strategy defined as the policy which maximizes welfare or minimizes costs in the society [15, 16, 21, 22]. Other objective functions used include minimizing a measure of economic loss subject to keeping infections within the capacity of the hospitals [59], minimizing total incidence of the disease subject to bounds on the intensity and/or total interventions [30], or minimizing peak prevalence of the disease [60].

We define the optimal strategy as the intervention path *G*(*t*) that minimizes total societal costs, *L_T_* in equation [7]. As the model is deterministic, there is no uncertainty about the evolution of the epidemic given the path of interventions *G*(*t*). By using the infectious disease model, *M*, each strategy *G*(*t*) gives a specific disease outcome-path, *S*(*t*), and an associated amount of voluntary social distancing per age group, *V ^i^*(*t*). The vector ***θ*** denotes the set of all the parameters in the model. The optimal strategy is given by

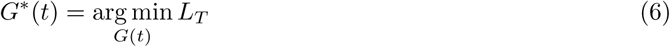

where

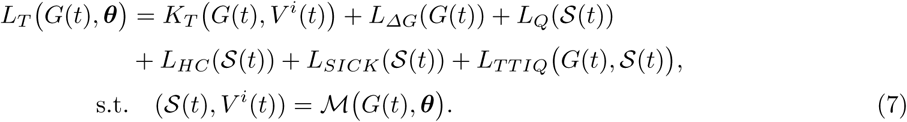

*K_T_* is the total costs from contact reduction given by Equation 5. *L_ΔG_* is the cost of changing the intervention level, *L_Q_* is the health loss from COVID-19 disease, *L_HC_* is the health loss from hospitals operating above capacity, *L_SICK_* is the economic and welfare cost when infected, and *L_TTIQ_* represents the cost of the TTIQ program including both administrative costs and economic and welfare costs associated with quarantine and isolation. All components are measured in monetary units.

We parameterize *G*(*t*) as a piecewise constant function and numerically find the optimal values of these constants. This is a challenging optimization due to the large number of parameters, non-convexity of the objective function, the need for derivative-free optimization and the comparatively slow runtime of the model. Because the objective function is non-convex, we compare solutions obtained using several parameterisations, derivative-free optimisation procedures, and starting paths representing qualitatively different strategies, including no intervention, sustained mitigation or suppression, and delayed stringent intervention. The reported optimal strategy is the lowest-cost solution obtained across these procedures. The complete optimisation procedure, including change points, algorithms and starting values, is described in *SI Appendix*. We assume that the epidemic starts with a small number of infected individuals (900), and that there is a limited inflow of infections from abroad (1 per day in baseline).

## 3 Results: Baseline

We first characterize optimal intervention strategies in the baseline parameterization. We consider a novel SARS-CoV-2-like respiratory pathogen with two key attributes: transmissibility, captured by the initial reproduction number *R ∈* [1.2, 3.0], and effective severity, measured by an index *ES ∈* [1, 20] that scales age-specific hospitalization and fatality risk proportionally. The duration and QALY-loss from infection is also increasing in effective severity, cf. *SI Appendix*, B1. *ES* = 1 is calibrated to the early Omicron wave with a high degree of immunity from vaccines and previous infections, while *ES ≈* 15 corresponds approximately to the original Wuhan variant in an immunologically näıve population.

Across the parameter space defined by transmissibility and effective severity, optimal policies cluster into three qualitatively distinct regimes: *Suppression*, *Delayed containment*, and *No/negligible intervention* (Figure 2.A)

**Figure 2:**
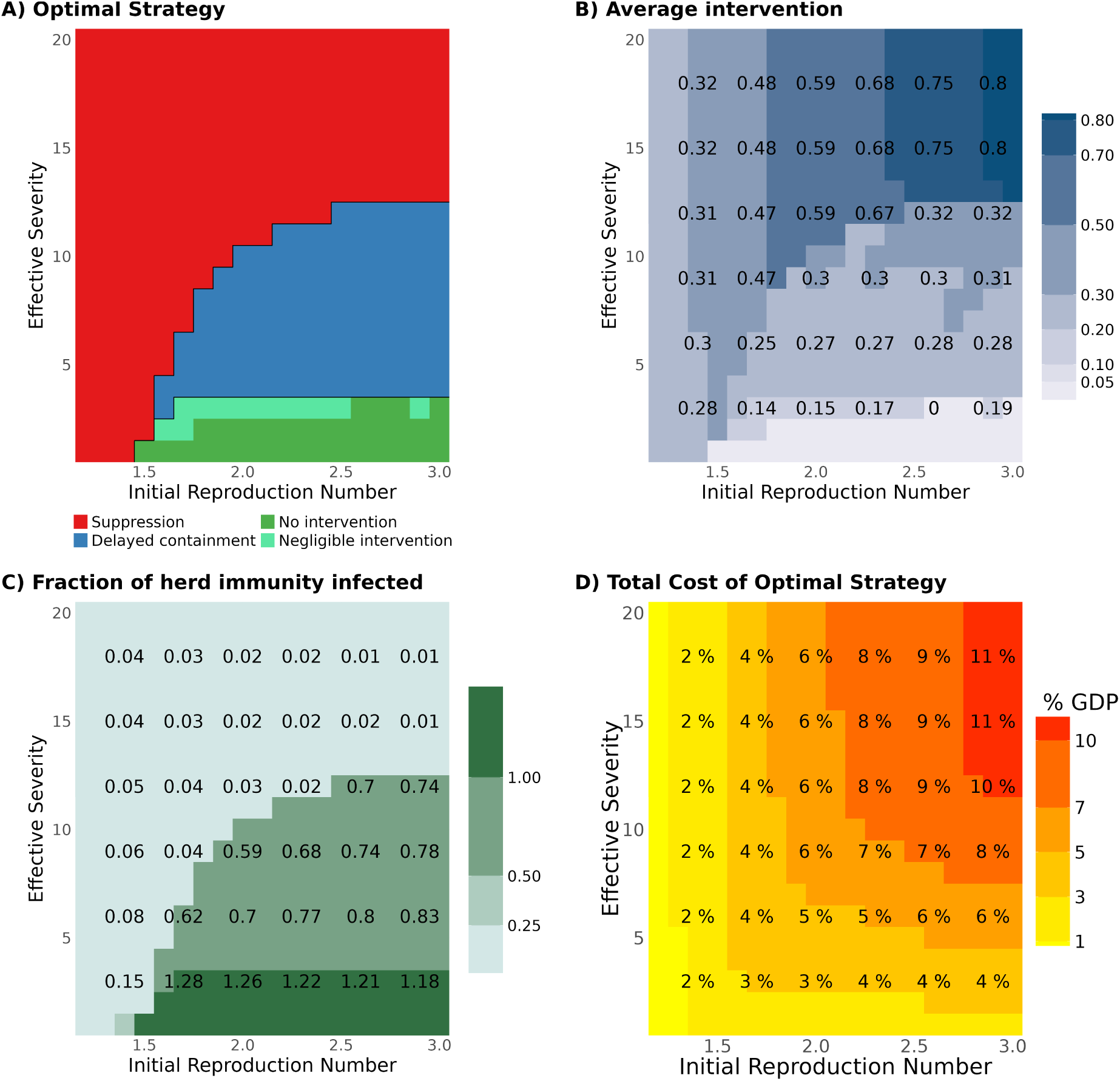
Optimal strategies in baseline. Each pixel represents one of the 19*·*20 = 380 possible (*R, ES*)-combinations, where *R ∈* [1.2, 3] and *ES ∈* [1, 20]. **Panel A:** Optimal strategy type. **Panel B:** Average intervention level. **Panel C:** The fraction of total infections under the optimal strategy compared to the herd immunity level (FHI). **Panel D:** The total cost, *L_T_*, as a percent of annual GDP.

For sufficiently high severity, the optimal strategy is *Suppression* (red area). In this regime, interventions reduce the effective reproduction number to around or below one, keeping incidence very low. *Suppression* is also optimal when transmissibility is relatively low (low *R*), because the epidemic can then be controlled at low cost through Test-Trace-Isolate-Quarantine (TTIQ).

For intermediate severity, the optimal strategy is characterized by *Delayed containment* (blue area), where infections are initially allowed to increase, followed by a short period of strict interventions which reduces transmission and prevents epidemic overshoot. Then low incidence can be maintained using TTIQ. As TTIQ is cost-efficient when infection levels are low, and sufficient to control the pandemic when the effective reproduction number is near one, this strategy keeps total infections well below the herd immunity threshold at fairly low costs.

For low severity, *No intervention* (green area) is optimal because the health loss from the epidemic is not sufficient to warrant the use of costly interventions. For some parameter values, the optimal strategy involves TTIQ part of the time. These are indicated by light green and termed *Negligible intervention*, defined as a strategy with a reduction of cumulative infections of less than 5% compared to infections under no interventions. Since TTIQ carries relatively low costs, this strategy may be optimal even if its epidemiological effect is limited.

Figure 2.B shows the associated average intervention until vaccination. Under *Suppression*, the required intervention level increases with *R*, and for *R* close to one, *Suppression* can be achieved by use of TTIQ only. The intervention level changes smoothly with *R* and *ES*, except at the boundaries between different strategy types, where discrete jumps occur.

Figure 2.C shows the sharp differences in outcomes across strategies, measured by the fraction of infections relative to the number of infections necessary to reach herd immunity (FHI). *Suppression* reduces the number of infections to a very low level, but not to zero. *Delayed containment* keeps infections at levels typically 15% to 35% below the herd immunity threshold, by use of TTIQ after a short period of strict interventions. Under *No/negligible intervention*, epidemic overshoot can push cumulative infections 15% to 30% above the herd immunity threshold, implying substantially higher health costs.

Finally, figure 2.D displays total costs as a share of GDP under the optimal strategy. Total costs are continuous in (*R, ES*) even though the optimal intervention level is discontinuous at strategy boundaries. Overall, optimal policies generally involve either sufficiently stringent intervention to suppress transmission or little general intervention. *Delayed containment* consists of a combination, where a period of no intervention is followed by a sharp intervention and subsequent control through TTIQ. Intermediate mitigation policies (Flatten-the-curve) are not optimal, as they involve both substantial health costs and substantial intervention costs, making them inferior to one of the more “extreme” strategies.

Our finding of distinct optimal strategy types and discontinuities in optimal interventions at parameter boundaries is consistent with previous work by [16, 27, 61]. The discontinuities are consistent with the existence of Skiba points [61], where qualitatively different strategies yield identical total costs. As emphasized by [16], this implies that otherwise similar countries may optimally choose very different strategies if they lie on opposite sides of a strategy boundary.

However, the classification of strategy types we obtain differs from the existing literature. [16, 27] identify suppression, mitigation (flatten-the-curve), or no intervention as relevant optimal regimes under different conditions. In contrast, we find that *Flatten-the-curve* is not optimal in any (*R, ES*)-scenario in the baseline case, while *Delayed containment* is optimal in many cases. The *Delayed containment* -strategy resembles results in [28–31], but these studies do not consider the subsequent transition to TTIQ. The combination of a delayed strict intervention and subsequent TTIQ implies that for a pathogen with transmissibility too high to be controlled by TTIQ alone, the initial spread reduces the effective reproduction number so that, after a sharp intervention, TTIQ alone can control the disease. Cumulative incidence can therefore be kept below the conventional herd immunity threshold at low cost until vaccination.

### 3.1 Intervention paths

Figure 3.A presents optimal intervention paths for selected (*R, ES*)-combinations. The top row with high severity (*ES* = 18) shows *Suppression* strategies, where interventions become stricter (higher *G*) for higher *R*. Because vaccine arrival is known, interventions are relaxed shortly before vaccination to reduce intervention costs.

**Figure 3:**
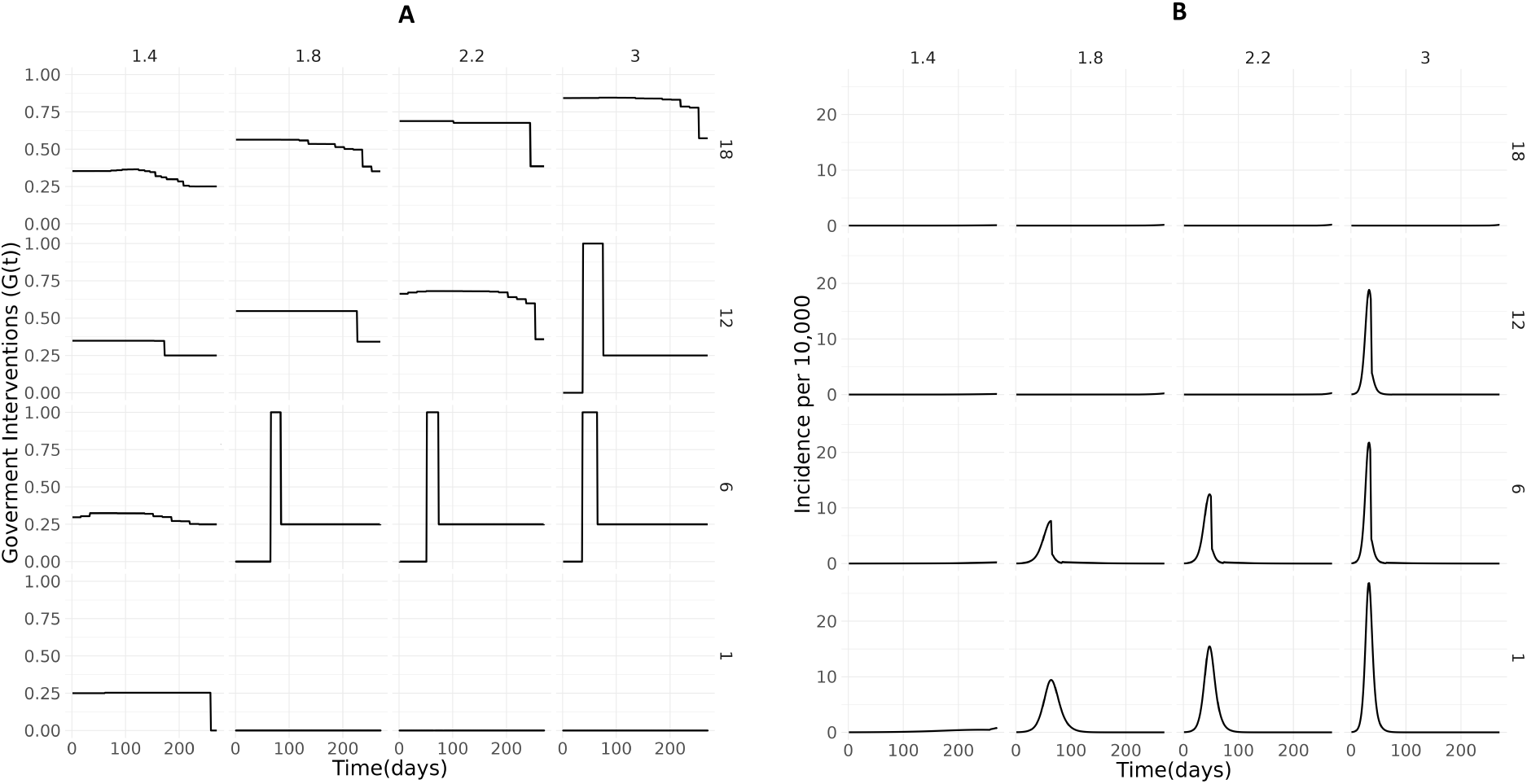
The baseline case: A. Optimal *G*(*t*)-path. B. Corresponding incidence over time. For a subset of initial reproduction numbers *R* = *{*1.4, 1.8, 2.2, 3*}* (columns) and severity levels *ES* = *{*1, 6, 12, 18*}* (rows), that gives 16 combinations.

For intermediate severity (*ES* = 12 and *ES* = 6), *Suppression* is optimal for low *R*, while *Delayed containment* is chosen for higher *R*. Under *Delayed containment*, infections initially increase before a sharp intervention reduces transmission. Afterwards, TTIQ is used at maximum (*G* = 0.25) to maintain low incidence, allowing the sharp intervention to end before herd immunity is reached.

The bottom row shows that for low severity (*ES* = 1), *Suppression* through constant TTIQ is sufficient when *R* = 1.4, while *No intervention* is optimal for higher values of *R*.

## 4 Results: Broad parameter scan

In this section, we examine how the optimal strategy depends on parameter values in a broad parameter scan along five dimensions: transmissibility, effective severity, the valuation of health losses relative to intervention costs, the time until effective vaccination, and the excess costs associated with an overburdened health sector. The aim is to identify robust qualitative policy regimes across a broad span of plausible future pandemic characteristics. High overcapacity-cost scenarios are included to explore the relevance of *Flatten-the-curve* strategies, since these are designed to protect the health sector. Table 1 presents the parameter values used in the scan, yielding 4320 = 9 *·* 10 *·* 4 *·* 3 *·* 4 parameter combinations.

**Table 1:** Parameter values for broad parameter scan.

| Parameter | Values |
| --- | --- |
| Initial reproduction number | $R \in \{1.4, 1.6, 1.8, 2.0, 2.2, 2.4, 2.6, 2.8, 3.0\}$ |
| Effective severity | $ES \in \{1, 3, 5, 7, 9, 11, 13, 15, 17, 19\}$ |
| Time to effective vaccine availability | $T \in \{270, 540, 1000, 1500\}$ days |
| Scaling baseline value of a QALY $s_Q \times COST_Q$ | $s_Q \in \{0.5, 1, 2\}$ |
| Scaling overcapacity costs $s_o \times L_{HC}$ | $s_o \in \{0.5, 1, 2, 5\}$ |

To classify optimal strategies, we use a parsimonious set of quantitative criteria that groups intervention paths and epidemiological outcomes with similar characteristics. The thresholds are informed by the pronounced clustering of the optimized paths documented in Section D.1 of the *SI Appendix*. They should be interpreted as classification rules rather than as complete definitions of the strategies.

*No intervention* refers to strategies with no interventions, while *Negligible intervention* refers to strategies where cumulative infections are reduced by less than 5% relative to no intervention. *Suppression* strategies are defined by sufficient interventions that keep infections low, but also allow some relaxation close to vaccine availability. Specifically, we require average relative intervention (ARI) to exceed 0.65 and relative max incidence (*RMI*) to remain below 0.3. *ARI* denotes the average intervention level relative to the intervention required for *R*_eff_ = 1, whereas *RMI* denotes the maximum incidence compared to maximum incidence under no interventions.

Conceptually, *Delayed containment* consists of limited initial intervention, followed by a delayed and short period of stringent intervention that sharply reduces incidence. The optimized paths displaying this pattern form a distinct cluster. We classify a path as *Delayed containment*, when interventions start no earlier than day 15 and the maximum intervention level (*MINT*) exceeds 0.3. The rule does not guarantee that the strict intervention is followed by containment through TTIQ, but in the simulations this is true in all the *Delayed containment* solutions. A less strict variant using only TTIQ is classified separately as *TTIQ-only*, where interventions also start after day 15 but peak at 0.25, corresponding to full TTIQ use.

There is also a limited number of optimal strategies that do not satisfy these requirements. We categorize a strategy as *Flatten-the-curve* if interventions prevent high peak incidence rates without preventing widespread cumulative infections, consistent with [16, 27]. Specifically, we define these as strategies with *RMI <* 0.5 and the fraction infected relative to the herd immunity threshold (FHI) above 0.60. (FHI is defined as the share of the population that would need immunity at *t* = 0 to obtain *R*_eff_ = 1). Strategies that do not fall into any of the other categories are grouped in *Other*.

Figure 4 shows the optimal strategies across the parameter space in Table 1. Panels A-B-C present results for various QALY-values relative to the baseline case. Note that the strategy choice depends on the relative valuation of health effects to intervention costs. Thus, a 100% increase in intervention costs - economic and welfare - has the same effect as a 50% reduction in the monetary value of a QALY. The four columns within each panel vary hospital overload costs, while the rows vary the time until effective vaccination from 270 to 1500 days.

**Figure 4:**
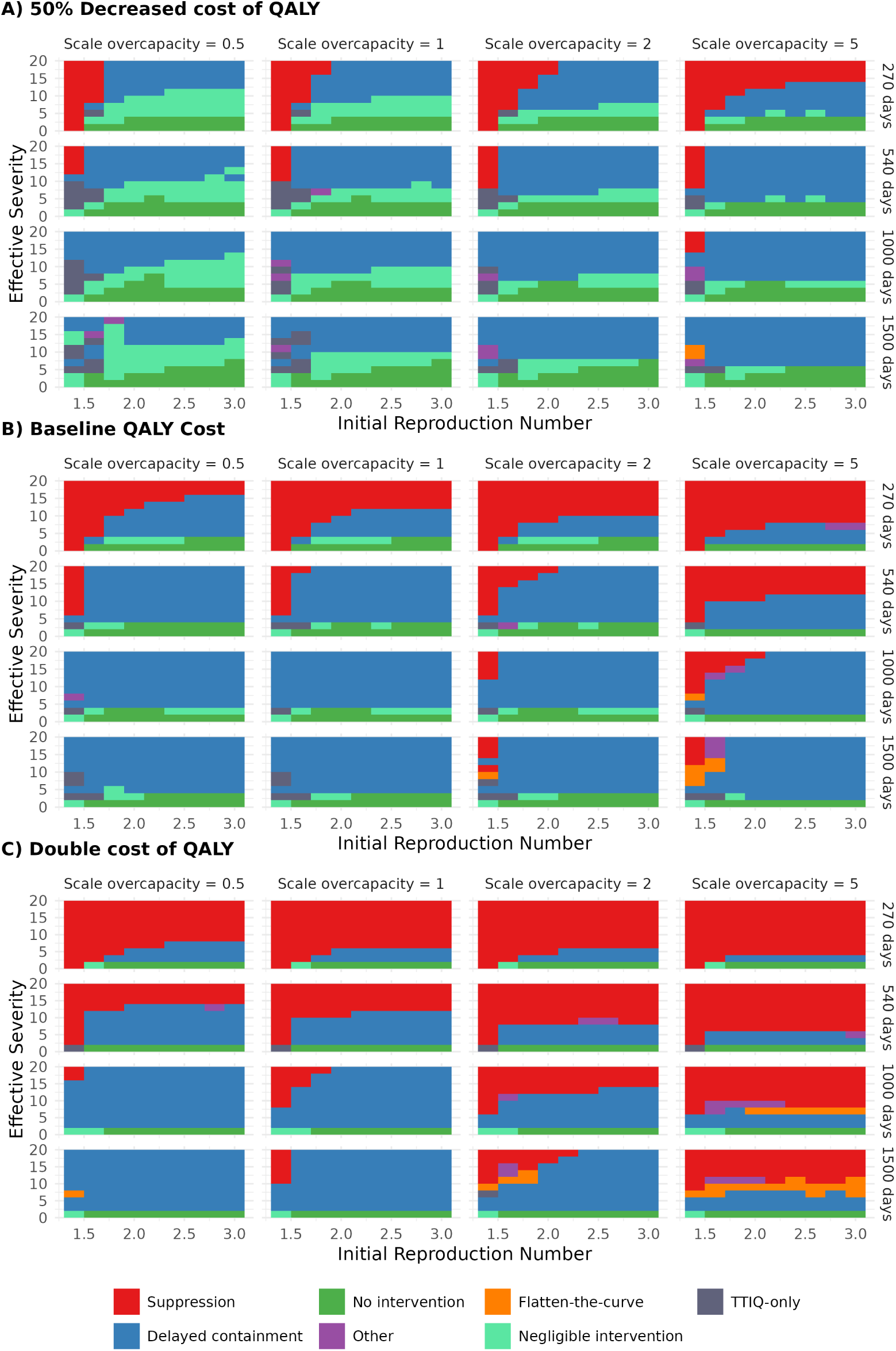
Parameter space scan. Optimal strategies for all parameter combinations in Table 1. Each scenario (pixel) is classified into strategy class based on the following rule: *No intervention*: ARI = 0; *Negligible intervention*: Reduction in infections relative to no intervention is less than 5%; *Suppression*: ARI *>* 0.65 and RMI *<* 0.3; *Delayed containment* : No interventions before day 15 and the maximum intervention level (*MINT*) exceeds 0.3; *TTIQ–only*: No interventions before day 15 and max intervention *≈* 0.25; *Flatten–the–curve*: FHI *>* 0.6 and RMI *<* 0.5; *Other* : None of the above.

Broadly, the qualitative results are in line with the results from the baseline calibration. Across all 4320 scenarios, *Delayed containment* is the optimal regime in more than half, and combined, *Suppression*, *No/negligible intervention*, and *Delayed containment/TTIQ-only*, account for more than 98% of optimal strategies. *Flatten-the-curve* and *Other* each account for less than 1% (Table S20 in *SI Appendix*).

*Suppression* is typically optimal when health costs are high, caused by high severity and/or a high QALY-valuation/high hospital overload costs. *Suppression* may also be optimal for low *R*, where the pandemic can be controlled at low cost through TTIQ. Delayed vaccination makes *Suppression* less attractive, since suppression costs increase roughly proportionally with duration, while *Delayed containment* and *No/negligible intervention* incur health costs mainly during a limited period with high infection rates.

When health costs are low, due to low severity and/or low QALY-valuation, *No/negligible intervention* dominates, while *Delayed containment* is typically optimal for intermediate health costs. Higher hospital overload costs increase the gains from intervention, shifting the thresholds between strategies downwards, ie. changing *Delayed containment* to *Suppression* and *No/negligible intervention* to *Delayed containment*. *Flatten-the-curve* is optimal for a small number of parameter combinations where late vaccination makes suppression too costly and high costs of overcapacity make it important to use interventions to protect the health sector. For the large majority of parameter combinations, *Flatten-the-curve* is less attractive, because it typically combines high health costs due to many infections with high intervention costs to protect the health sector.

Section D.7 in *SI Appendix* shows that the qualitative pattern remains unchanged without TTIQ: optimal strategies are typically either *Suppression*, delayed stringent intervention (without the subsequent TTIQ), or *No/negligible intervention*, while *Flatten-the-curve* appears only in a few cases with long duration and high overcapacity costs.

Section D.8.1 in the *SI Appendix* illustrates the costs of policy mistakes using predetermined strategies. For parameter combinations where *Delayed containment* is optimal, maintaining ICU admissions below capacity at all times may increase total costs substantially, in extreme cases by more than 600%. By contrast, when *Suppression* is optimal, ICU admissions usually remain below capacity without additional restrictions.

### 4.1 The effect of strategy type on infections

Figure 5 shows cumulative infections, measured by the fraction of herd immunity (*FHI*), across strategy types. *Suppression* keeps infections low, generally resulting in *FHI <* 0.25. *Delayed containment* allows temporary spread before strict intervention followed by TTIQ keeps incidence below the herd immunity level, and in most cases *FHI <* 0.9. Under *No/negligible intervention*, epidemic overshoot is substantial and *FHI* always exceeds one. *SI Appendix*, Sections D.1 and D.3 provide more information on the effects of different strategy types on infections and interventions.

**Figure 5:**
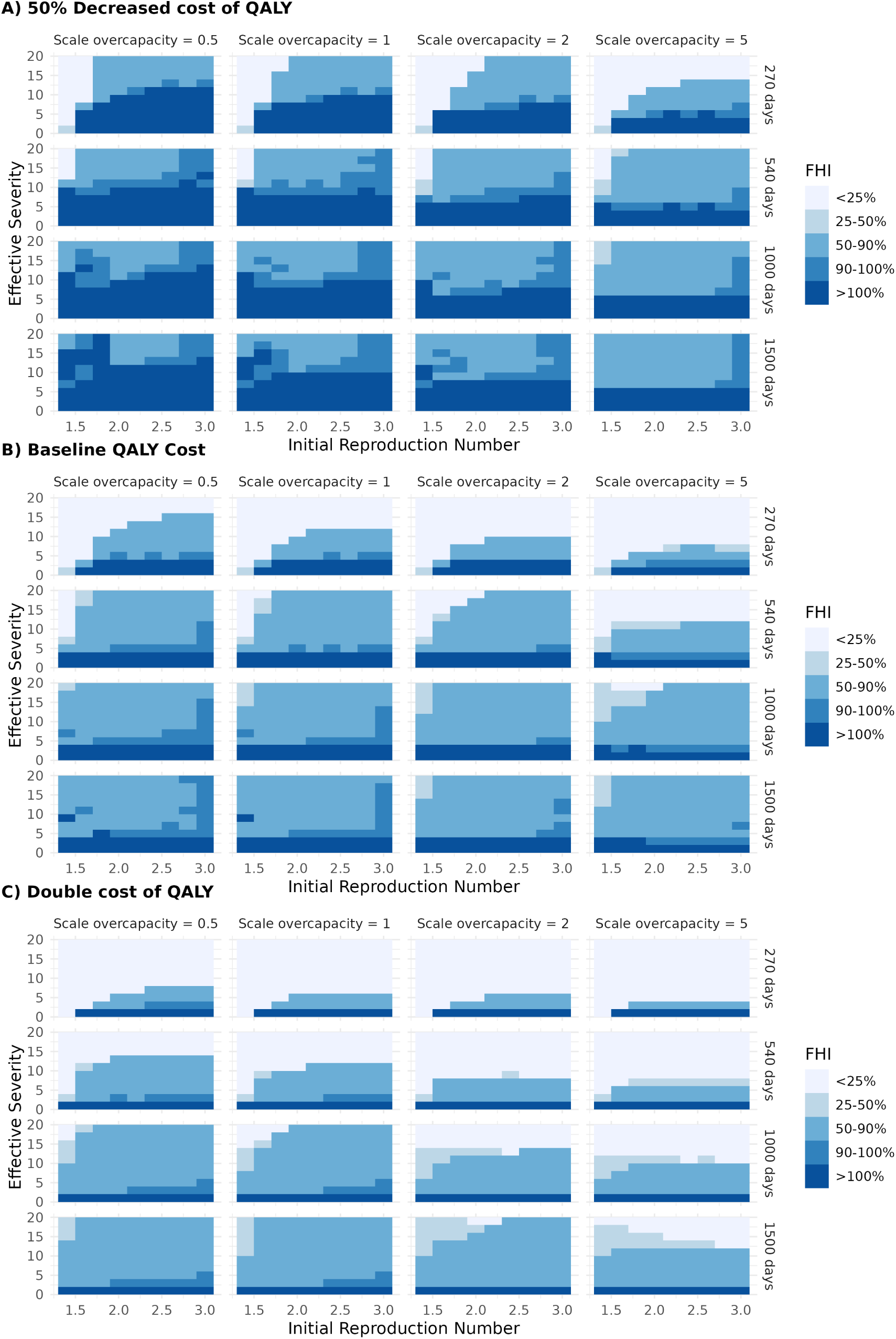
Parameter scan. Optimal strategies: Fraction of herd immunity *FHI* for the optimal strategies in Figure 4.

## 5 The effects of voluntary social distancing (VSD)

Figure 6 shows the effects of stronger VSD representing a form of altruism, where individuals take into account both their own health risk and the expected health costs imposed on others through transmission. Panel I shows that the stronger VSD under altruism reduces the use of government interventions, implying that *TTIQ-only* strategies are often optimal.

**Figure 6:**
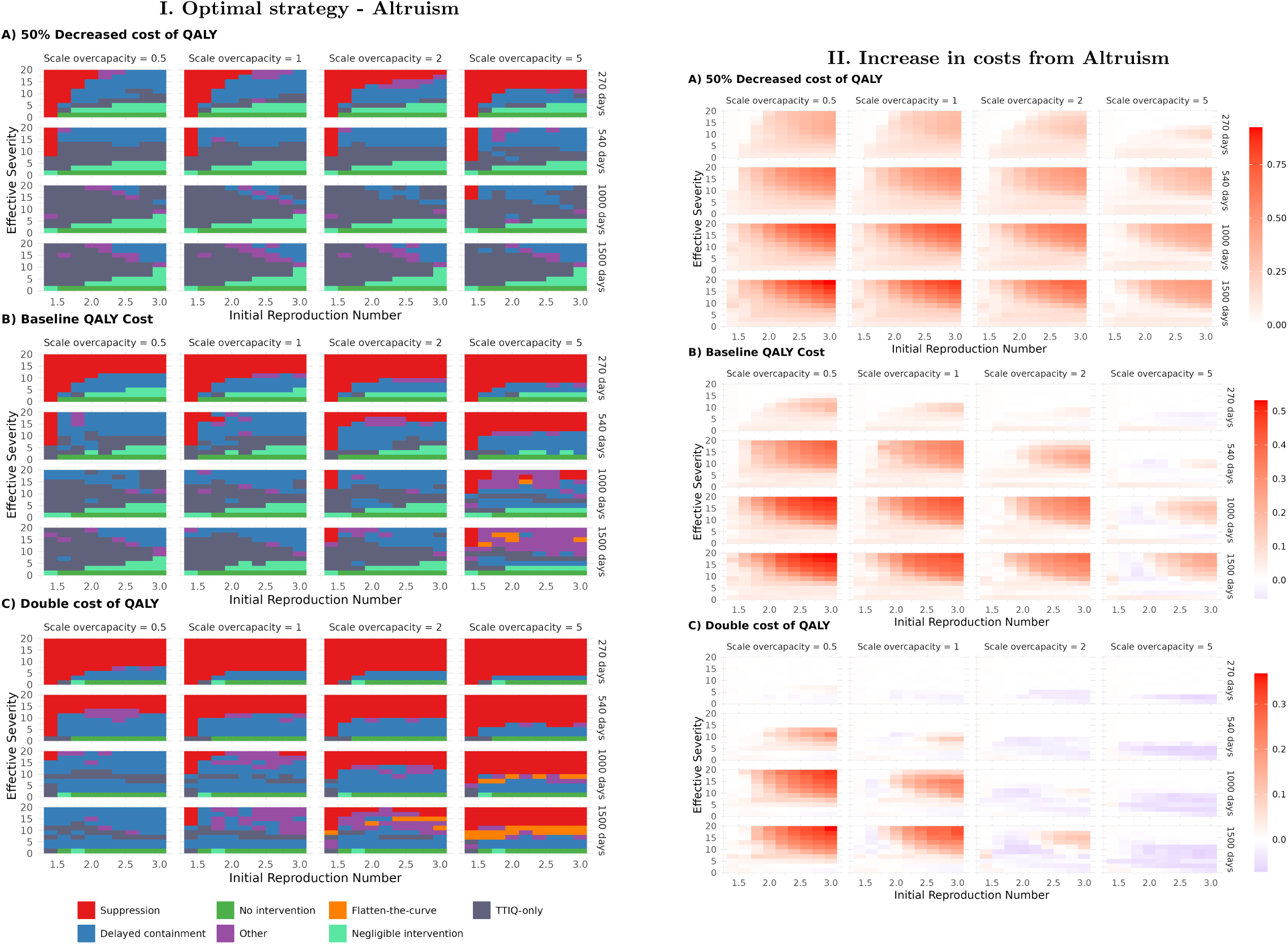
Altruism. Left: Optimal strategy with altruism. Right: Increase in cost with altruism, relative to baseline VSD. Red cells indicate higher costs with altruism, while blue cells indicate lower costs. Strategy types are classified as in Figure 4.

Figure 6 panel II shows that altruism, as it is represented in our framework, leads to higher total costs than baseline VSD in most scenarios. Altruism causes younger individuals to reduce contacts, slowing transmission and prolonging the epidemic wave. In some scenarios, this may change the optimal strategy from *Delayed containment* to *No/negligible intervention*, implying higher overall infection rates. In other scenarios, it may increase infection risk among older groups for unchanged optimal strategy, by shifting infections from younger to older age groups. Altruism may also reduce welfare through lower social interaction. When health costs are very high, due to high QALY values and high overcapacity costs, altruism may reduce total costs by lowering peak incidence and reducing healthcare overload.

Figure 7 shows the effects if there were no VSD. This generally leads to stricter interventions, where *Suppression* is used in more parameter combinations, and *No/negligible intervention* is largely replaced by *Delayed containment* or *Flatten-the-curve*. The reason is that without VSD, periods of high incidence involve substantially larger health costs among vulnerable older groups.

**Figure 7:**
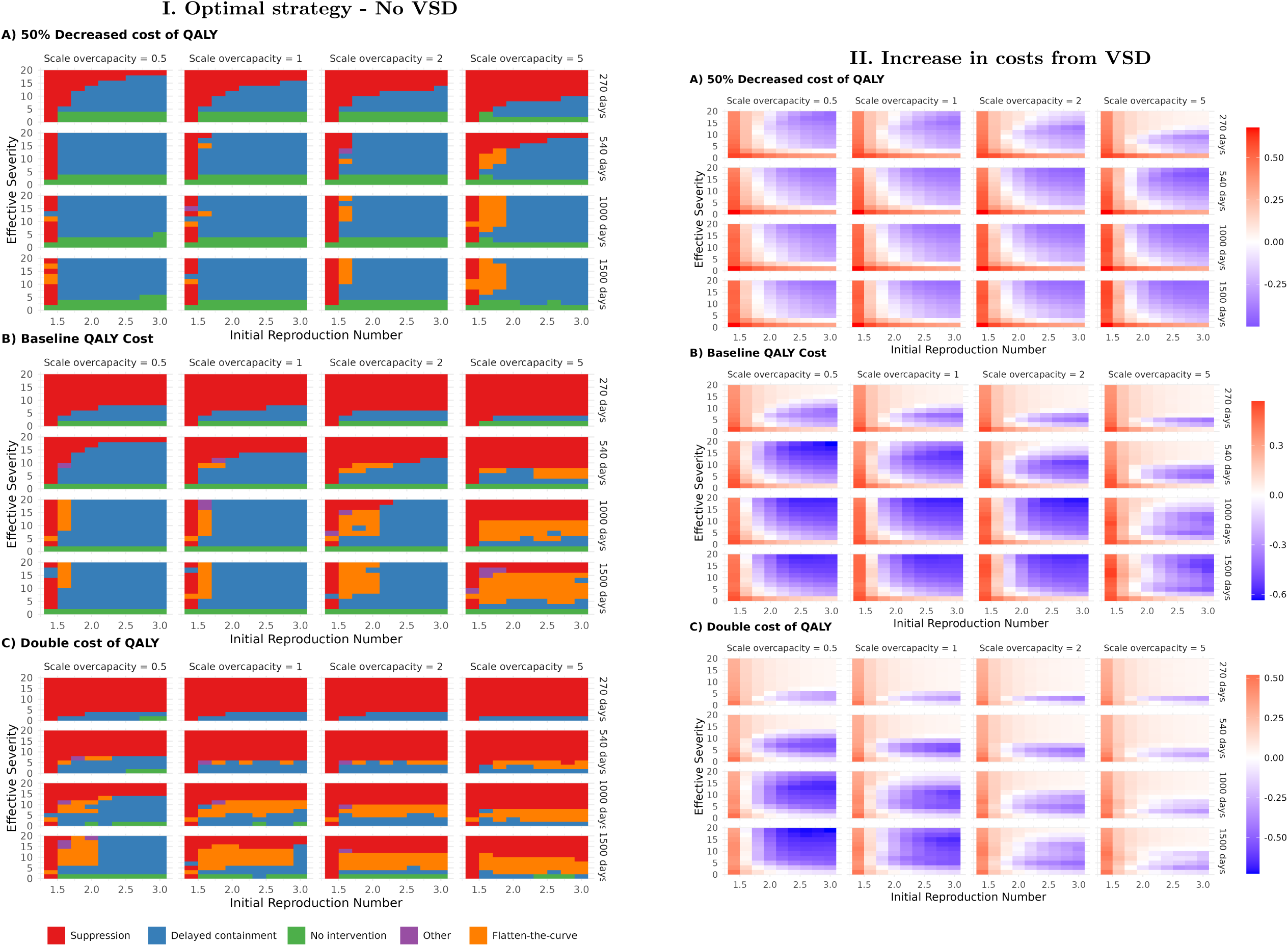
No voluntary social distancing (VSD). Left: Optimal strategy with no VSD. Right: Increase in costs with VSD, relative to costs without VSD. Red cells indicate higher costs with VSD, while blue cells indicate lower costs with VSD.

The experiences from COVID-19 where contact activity fell substantially also in periods and areas without government restrictions, provide strong evidence for the importance of VSD [5, 21–23, 37, 39]. This implies that the baseline specification with VSD is more policy relevant than the specification with no VSD. Thus, these results suggest that *Flatten-the-curve* may seem attractive if one neglects VSD, even if *Delayed containment* or *No/negligible intervention* yield lower total costs once behavioral responses are taken into account.

Figure 7, panel II, shows that VSD has opposing effects on total costs, consistent with theoretical contributions emphasizing both positive and negative externalities of precautionary behavior [18, 22, 29, 62]. For low *R*-values, the epidemic can be controlled at relatively low cost through TTIQ, so additional voluntary contact reduction generates welfare losses not fully offset by health gains. For higher *R*-values, where costly interventions are needed to contain infections, VSD reduces total costs in most parameter combinations.

The mixed effects of VSD on total costs reflect that voluntary behavior and government interventions have different strengths. VSD adapts to individual risk, with vulnerable groups reducing contacts more strongly, whereas government interventions can account for broader equilibrium effects. When *Delayed containment* or *No/negligible intervention* is optimal, voluntary reduction in contacts among younger groups may increase total costs because it slows the accumulation of population immunity while imposing additional welfare costs.

Taken together, Figure 6 and Figure 7 show that behavioral responses are crucial for the choice of optimal strategy. If information and guidance can influence behavioral responses in the population at large, or for specific groups, this would give a potential for substantial gains. The model can be used to inform such guidance.

## 6 Using the model to evaluate preparedness and policy measures

In this section we return to the baseline case with effective vaccination after 270 days and baseline assumptions on the value of a QALY and overcapacity costs (*s_Q_*= 1 and *s_o_*= 1). We show how the model can be used to quantify the gains from reduced health and/or intervention costs from specific policy changes. In decisions on investments or policy interventions, these estimates should be compared to estimates of the costs of undertaking the policy.

The left panels of Figure 8 show the optimal strategy after each policy change, while the right panels show the associated effect on total costs.

**Figure 8:**
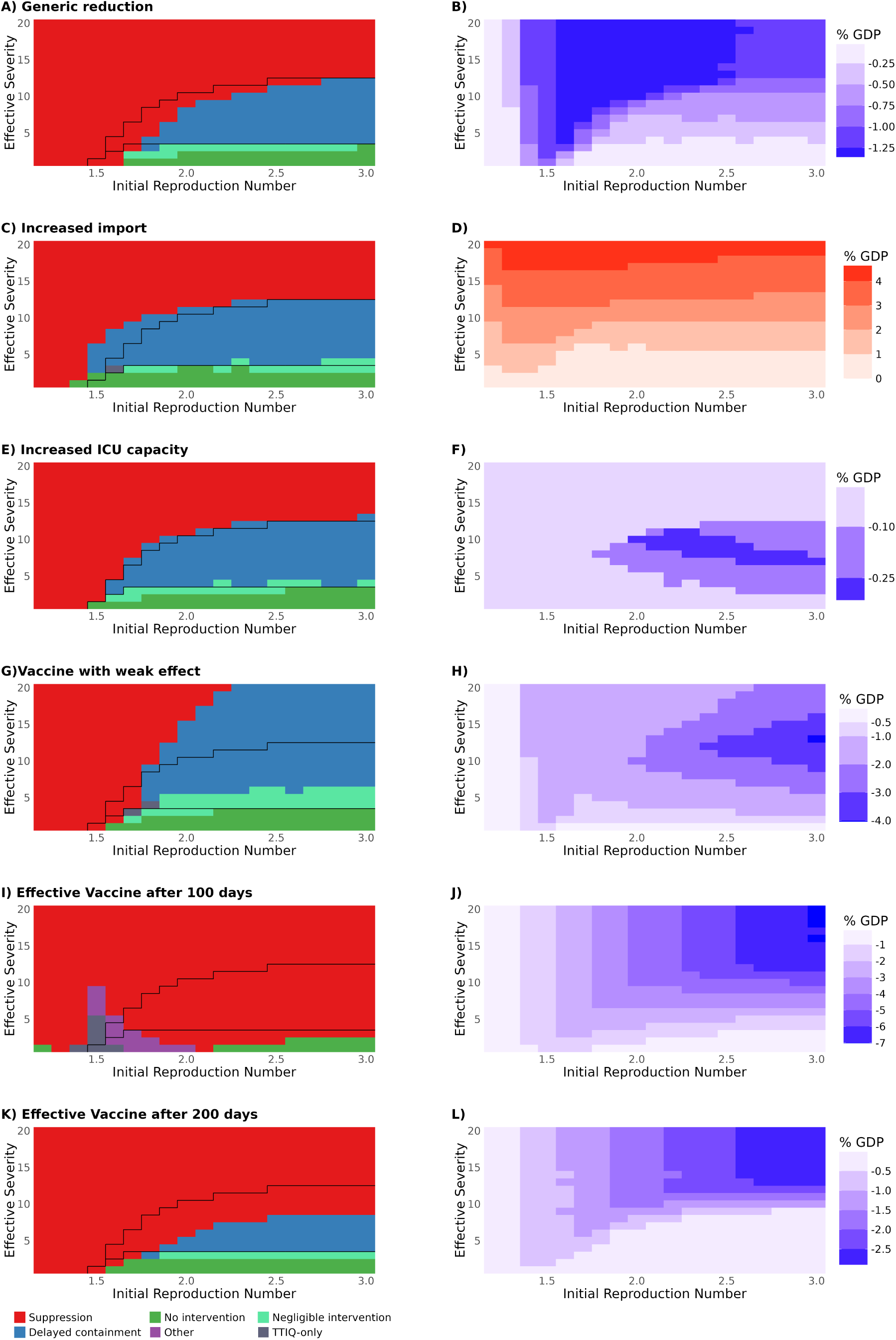
Effect of preparedness and policy measures on optimal strategy and total costs. The left panels show the optimal strategy as a function of *R* and *ES* for different policy deviations from the baseline case. The black line indicates the strategy boundary in the baseline case. The right panels show the associated costs or gains compared to the baseline case, with red indicating higher costs and blue indicating savings. **Panel A-B:** 10% reduction in transmission. **Panel C-D:** Increased import of infections. **Panel E-F:** Increased ICU capacity. **Panel G-H:** Vaccination with weak effect. **Panel I-J:** Effective vaccine available after 100 days. **Panel K-L:** Effective vaccine available after 200 days.

Measures that reduce transmission, such as improved ventilation or mask use, are particularly valuable under suppression strategies because they lower the intensity and duration of costly interventions required to maintain low incidence (Figures 8.A–B). By contrast, the same measures have more limited effects when widespread transmission is already allowed.

Figure 8.C and 8.D show that increased daily inflow of infected individuals makes *Suppression* less attractive, as stricter interventions are required to offset imported infections. In contrast, when *No intervention* is optimal, additional imports have little effect on total costs. This illustrates that strict border controls can be valuable as part of a suppression strategy, while the effect is likely to be much smaller without other contact-reducing interventions, consistent with [63].

Similarly, increases in healthcare capacity mainly reduce societal costs in scenarios where incidence becomes sufficiently high for healthcare constraints to bind (Figures 8.E–F). Under suppression strategies, healthcare capacity constraints are often less important because incidence remains low.

Figures 8.G and 8.H show that a partially effective vaccine (e.g. one targeted to older strains) that reduces susceptibility to infection and the risk of severe outcomes, reduces health costs substantially for variants with high transmissibility and moderate-to-high severity.

The gain from earlier availability of vaccination depends strongly on the optimal strategy (Figures 8.I–L). This increases the attractiveness of suppression because the duration of stringent interventions required to bridge the period until vaccination becomes shorter.

Overall, these analyses show that the effectiveness of individual policy measures depends critically on the broader intervention strategy within which they are implemented. This implies that preparedness measures cannot be evaluated independently of the intervention strategy they are planned to support.

## 7 Discussion

Using an integrated epi–econ framework, we show that optimal intervention strategies cluster into three distinct categories: *Suppression*, *Delayed containment* and *No/negligible intervention*. By contrast, prolonged mitigation strategies aimed at flattening the curve are optimal only in a narrow set of scenarios. This pattern reflects a non-convexity in the trade-offs governing pandemic control. Interventions that only partially reduce transmission generate considerable economic and welfare costs, but the reduction in health loss is often limited.

Our findings provide a unifying perspective on previously divergent results in the literature. Broad analyses have emphasized suppression, mitigation, and limited intervention, while optimal-control studies have shown the possible value of delayed stringent intervention. Work on testing and tracing has shown why targeted measures are particularly valuable at low prevalence. In our broad and flexible parameter scan, we identify optimal strategy types that integrate elements from all these approaches, as well as novel combinations.

Our finding of distinct optimal strategy types and discontinuities in optimal interventions at parameter boundaries is consistent with previous work by [16, 27]. However, the classification of strategy types differs, as [16, 27] find that the optimal strategies are *Suppression*, *Mitigation/Flatten-the-curve* and *No intervention*. In contrast, we find that *Mitigation/Flatten-the-curve* is only optimal in a narrow set of scenarios, while *Delayed containment* is optimal in many cases. The Delayed containment strategy resembles results in [28–31], with the additional feature that the strict intervention period is followed by TTIQ.

The *Delayed containment* strategy emerges as a particularly important intermediate regime. For a pathogen with transmissibility too high to be controlled by TTIQ alone, limited initial spread increases population immunity and thereby reduces its reproduction potential. A subsequent stringent intervention brings incidence down. When incidence is low and the effective reproduction number is sufficiently close to one, TTIQ can maintain control, keeping cumulative infections well below the conventional herd-immunity level, at substantially lower cost than continued general contact-reducing interventions. When the cost of maintaining *Suppression* until vaccination is too high, *Delayed containment* can substantially reduce health costs relative to *No/negligible intervention*, while avoiding the prolonged intervention costs of *Suppression*. However, the feasibility of a *Delayed containment* strategy depends critically on strong pandemic surveillance and timely information on epidemic progression and population immunity. Moreover, the early phase with increasing infections and limited interventions may pose important challenges in terms of pressure on hospitals as well as communication and trust in the population.

Behavioral responses play an important role in shaping optimal interventions. Evidence from the COVID-19 pandemic shows that voluntary behavioral responses partly substitute for government interventions and contribute to reducing transmission even in the absence of stringent policies [21, 22, 37, 39, 64]. At the same time, behavioral responses may also generate welfare losses associated with reduced social and economic activity, implying that the net effect on societal costs is ambiguous and depends on epidemiological conditions. Perhaps surprisingly, we find that altruism, as it is represented in our framework, increases costs in most scenarios because it involves reduced social interaction and slows transmission among low-risk groups, thereby prolonging the epidemic wave. Consistent with [48], we find that it is crucial to take the extent and type of VSD into account when deciding on government interventions. Guidelines on individual behavior should be tailored to the epidemiological context, and the model can be used to design guidance and quantify potential cost reductions.

The cost-minimizing strategies identified here may be politically difficult to sustain. Under *suppression*, interventions impose visible economic and welfare costs while few people are ill. In contrast, under *no interventions*, health losses and pressure on hospitals may generate fear and criticism, while the benefits of avoiding restrictions are less visible. Although a calibrated model cannot resolve these political and ethical tensions, it can clarify the trade-offs associated with alternative strategies.

The analysis has several limitations. First, we abstract from uncertainty and assume that key epidemiological characteristics, including vaccine timing, are known. In practice, policymakers operate under substantial uncertainty, particularly during the early phase of a pandemic. Uncertainty may affect both the timing and robustness of optimal interventions. However, the underlying mechanisms and non-convexity of the trade-offs are likely to remain relevant even under extensive uncertainty. If the characteristics of a virus are unknown, but there is enough information to establish a meaningful probability distribution for its transmissibility and severity, the model can be used to calculate the expected costs of different strategies, allowing authorities to choose the one with the lowest expected cost, conditional on the other assumptions of the model. The model can also be used to explore the robustness of a specific policy to alternative assumptions regarding the virus and the effect of interventions.

Second, the model is calibrated to a high-income country with a well-educated population, generally good housing conditions, good access to telecommunications, low inequality and a high level of trust. These conditions may facilitate pandemic control [65]. As a result, both the effectiveness and the costs of interventions in our model may differ in other settings. We also restrict attention to general contact-reducing interventions (NPIs) and TTIQ, with no differentiation across population groups. In a pandemic with large differences in risk across age-groups, there may be gains from differentiating interventions and protective measures across age groups [20, 24, 66]. The effects of such policies can be explored in our framework and this is an important avenue for future work.

Third, to focus on the choice of overall intervention stringency, we do not distinguish between different types of interventions. There is strong empirical evidence that interventions work [1–4, 6–9, 67], but the evidence on the effects of specific interventions is more uncertain and often conflicting, as illustrated by the discussion of the literature in [4]. To make concrete prescriptions for specific interventions, the model must be supplemented with estimates for the effects and costs of the various types of interventions.

Fourth, we assume that individuals can be infected at most once during the time horizon considered. Reinfections and immune escape may become important in longer-lasting pandemics or for pathogens with rapidly evolving variants [68, 69]. Extending the framework to incorporate reinfections and endogenous pathogen evolution would therefore be a useful direction for future research.

Fifth, we have neglected seasonal effects and regional differences in the spread of the virus. Such effects could be incorporated in the model, and this might be important for practical use in a real pandemic. Likewise, we assume that disease severity and the effectiveness and costs of interventions remain constant over time. This ignores changes in pathogen characteristics, improvements in medical treatment, and evolving evidence on intervention effectiveness as we observed during COVID-19 [8, 51, 70–72]. Consequently, the model abstracts from an important mechanism: if the expected cost of infection declines over time because of improved treatment, vaccination, or lower virulence, delaying infections becomes more valuable, potentially increasing the optimal duration of early suppression before interventions are relaxed.

Finally, the analysis evaluates policies using a unified societal cost framework combining health losses, economic costs, and welfare losses. While such aggregation is necessary for optimization, real-world policymaking also involves distributional, ethical, legal, and political considerations that are not captured in the model. The results should therefore be interpreted as characterizing benchmark cost-minimizing strategies under specified assumptions rather than fully describing all relevant dimensions of pandemic policymaking. In section D.9 in *SI Appendix*, we show that the cost-minimizing strategy may differ from the strategy preferred by a majority of individuals because health and intervention costs are distributed unevenly across age groups.

Despite these limitations, the framework provides a flexible tool for evaluating pandemic preparedness and informing real-time intervention strategies. By integrating epidemiological dynamics, behavioral responses, healthcare constraints, and societal costs, the model allows systematic comparison of alternative strategies across a wide range of plausible future pandemic scenarios. If a new pandemic emerges, an updated country-specific calibration can inform the choice of the intervention path. As part of pandemic preparedness, countries may benefit from investing in both epi–econ modeling capacity and pandemic surveillance systems.

## Data Availability

All computer code used for processing and data analysis is available at: https://github.com/folkehelseinstituttet/covid19_economic_evaluation

## 8 Acknowledgments

We thank seminar participants at presentations at the National Institute of Public Health, the Workshop for Pandemic Preparedness in Stockholm and at the University of Birmingham for valuable feedback, and members of the expert group behind (Holden et al., 2022) for excellent collaboration.

## Funding

1. S. H. has received financial support from The Research Council of Norway, grant no 324615.

## Author contributions

**S.H.**: Conceptualization, Methodology, Writing - Original Draft. **G.R.**: Conceptualization, Methodology, Software, Formal analysis, Writing - Original Draft. **I.H.**: Project administration, Visualization, Writing - Review & Editing.

## Competing interests

There are no competing interests to declare.

## Data and materials availability

All computer code used for processing and data analysis is available at:[73]

## Notes

### Competing Interest Statement

The authors have declared no competing interest.

### Summary of Updates

This version substantially revises the manuscript, including a broad parameter scan of optimal intervention strategies, additional analyses of behavioral responses and pandemic preparedness measures, and substantial revisions to the presentation and discussion.

