## Supplementary information for "Suppression, delayed containment, or no intervention: optimal strategies for pandemic control"

August 19, 2026

**This appendix includes:**

**A Materials and Methods** 1
D.11 Discussion: Effective vaccine timing 62

[*]University of Oslo:
[†]Norwegian Institute of Public Health:
[‡]BI Norwegian Business school:

### A Materials and Methods

#### A.1 Data availability

We use a deterministic age-stratified metapopulation model to describe the spread of a new virus variant in Norway.[1] All the code and data used in this analysis can be found at the GitHub site:[2] https://github.com/folkehelseinstituttet/covid19_economic_evaluation .

#### A.2 The model

In the baseline calibration, we consider the arrival of a novel SARS-CoV-2-like respiratory pathogen, described by two key attributes: an initial reproduction number $R \in [1.2, 3]$, and effective severity $ES \in [1, 20]$, mapped on the existing Norwegian population. The spread of the epidemic is affected by both government interventions like Test-Trace-Isolate-Quarantine (TTIQ) and non-pharmaceutical interventions (NPIs) involving general contact reduction, and voluntary social distancing. To explore the epidemic trajectories, we use a metapopulation SEIR-type model with nine age groups. The aim of the analysis is to explore the optimal government intervention strategy, defined as the intervention strategy that minimizes the total costs from the pandemic and interventions.

##### A.2.1 The epidemiological model

The model follows the natural evolution of the disease and the disease stages for each age group, including vaccination, denoted by the topscript $i, j \in \mathcal{N}$. As a result, our model encompasses 18 sub-populations, $\mathcal{N} = \{1, ..., 18\}$, which are distinguished based on their age and vaccine status. The model is calibrated to the Norwegian population.

The model is defined by the differential equations (1)-(12) where the variables in each equation are described in Table S1. The variable $\Lambda_i$ is the force of infection, described in equation (13). The parameter $import^i$ is the number of infected individuals coming from abroad daily. The description and values of the parameters $D_L$, $D_P$, $D_A$ and $D_I$ are given in Table S2. Further, the description and values of the parameters $f_a^i$, $f_s^i$, $p_{hosp}^i$, $p_{ICU}^i$, $p_{death}^i$, $D_H^i$, $D_{ICU_1}^i$, $D_{ICU_2}^i$ and $D_{ICU_3}^i$ are given in Table S3.

$$\frac{dS^i}{dt} = -\Lambda_i S^i - import^i(t), \tag{1}$$

$$\frac{dE_a^i}{dt} = f_a^i \Lambda_i S^i - \frac{1}{D_L} E_a^i, \tag{2}$$

$$\frac{dE_s^i}{dt} = f_s^i \Lambda_i S^i - \frac{1}{D_L} E_s^i, \tag{3}$$

$$\frac{dP^i}{dt} = \frac{1}{D_L} E_s^i - \frac{1}{D_P} P^i, \tag{4}$$

$$\frac{dA^i}{dt} = \frac{1}{D_L} E_a^i - \frac{1}{D_A} A^i, \tag{5}$$

[1] Our model is similar to other models applied by the Norwegian Institute of Public Health (NIPH) during the Covid-19 pandemic in Norway. See [1, 2] for a detailed description of these models. Similar metapopulation models are used in other studies, for example [3, 4].

[2] The model is implemented in the R-programming language [5] using the odin package [6].

$$\frac{dI^i}{dt} = \frac{1}{D_P}P^i - \frac{1}{D_I}I^i + import^i(t), \tag{6}$$

$$\frac{dH^i}{dt} = p^i_{hosp}(1-p^i_{icu})\frac{1}{D_I}I^i - \frac{1}{D^i_H}H^i, \tag{7}$$

$$\frac{dICU^i_1}{dt} = p^i_{hosp}p^i_{icu}\frac{1}{D_I}I^i - \frac{1}{D^i_{ICU_1}}ICU^i_1, \tag{8}$$

$$\frac{dICU^i_2}{dt} = \frac{1}{D^i_{ICU_1}}ICU^i_1 - \frac{1}{D^i_{ICU_2}}ICU^i_2, \tag{9}$$

$$\frac{dICU^i_3}{dt} = \frac{1}{D^i_{ICU_2}}ICU^i_2 - \frac{1}{D^i_{ICU_3}}ICU^i_3, \tag{10}$$

$$\frac{dR^i}{dt} = (1-p^i_{hosp})(1-p^i_{death})\frac{1}{D_I}I^i + (1-p^i_{death})\frac{1}{D^i_H}H^i + (1-p^i_{death})\frac{1}{D^i_{ICU_3}}ICU^i_3, \tag{11}$$

$$\frac{dD^i}{dt} = (1-p^i_{hosp})p^i_{death}\frac{1}{D_I}I^i + p^i_{death}\frac{1}{D^i_H}H^i + p^i_{death}\frac{1}{D^i_{ICU_3}}ICU^i_3, \tag{12}$$

In the model, the infection process is determined by the force of infection, $\Lambda_i$, given by:

$$\Lambda_i(t) = \sum_j \beta(t)(1-C^i_T(t))sus_i M_{ij}\Big(inf_p P^j(t) + inf_a A^j(t) + inf_s I^j(t)\Big), \tag{13}$$

where the parameter $\beta$ is the overall transmissibility set to get the correct initial reproduction number, $R$. $C^i_T(t)$ is the contact reduction in the population, described in section A.2.2. The parameter $M_{ij}$ is the age-dependent contact-matrix for which we use data from [7], a 2017 POLYMOD-like study from Norway. The parameter $sus_i$ combines the susceptibility effect from changing the susceptibility for symptomatic, $sus^i_s$, and asymptomatic, $sus^i_a$, separately:

$$sus_i = f^i_a sus^i_a + f^i_s sus^i_s. \tag{14}$$

Further, the parameters $inf_p$, $inf_a$ and $inf_s$ are described in Table S2.

##### A.2.2 Contact reduction

Both government interventions and individual responses affect the infectious disease model via the $C^i_T(t)$-parameter. $C^i_T(t)$ is the total change in the contact rate for age group $i$ at time $t$. An individual's contact rate is influenced by government interventions $G(t)$ and the age-specific voluntary distancing $V^i$, as in equation (15):

$$C^i_T(t) = 0.8(G(t) + V(t)^i) - 0.7(G(t) \times V(t)^i). \tag{15}$$

This formulation is chosen to capture that government interventions $G$ and voluntary behavior response $V^i$ to a large extent will overlap, but they may also complement each other. The formulation ensures that $C^i_T = 0.9$, i.e. a 90% reduction in the contact rate, for $G = V^i = 1$, while $C^i_T = 0.8$ if government interventions are at full intensity and no VSD, or vice versa $(G = 1; V^i = 0)$ or $(G = 0; V^i = 1)$. Further details on the model specification and the calibration of costs related to government interventions and voluntary social distancing are provided in Sections B and C.

##### A.2.3 Cost minimization

We define the "optimal strategy" as the time-varying level of interventions, the $G(t)$-path, imposed by the government that minimizes total costs in society arising from the virus itself, individual behavior and governmental interventions. Concretely, we want to find the vector of daily $G$-values such that total costs to society, $L_T$, are minimized:

$$G^*(t) = \arg\min_{G(t)} L_T \tag{16}$$

where

$$\begin{aligned} L_T\big(G(t), \boldsymbol{\theta}\big) &= K_T\big(G(t), V_T(t)\big) + L_{\Delta G}(G(t)) + L_Q(\mathcal{S}(t)) + L_{HC}(\mathcal{S}(t)) \\ &\quad + L_{SICK}(\mathcal{S}(t)) + L_{TTIQ}\big(G(t), \mathcal{S}(t)\big), \\ &\text{s.t.} \quad \big(\mathcal{S}(t), V^i(t)\big) = \mathcal{M}\big(G(t), \boldsymbol{\theta}\big) \end{aligned} \tag{17}$$

Details on the different cost variables within the total costs, $L_T$, are described in section B.

We perform this optimization by the direct method where we parameterize $G(t)$ as a piece-wise constant function and then use a numerical optimization algorithm to find the optimal $G(t)$, see section A.4.

As the model is deterministic, there is no uncertainty regarding how the evolution of the pandemic depends on the path of interventions $G(t)$. Using the infectious disease model $\mathcal{M}$, each strategy $G(t)$ yields a specific disease outcome space $\mathcal{S}(t)$ and an associated level of voluntary social distancing $V^i(t)$ for each age group $i$ in period $t$. $\mathcal{S}(t)$ is a matrix representing the complete disease state, that is, the number of individuals in each disease compartment over time. The vector $\boldsymbol{\theta}$ denotes the full set of model parameters, including the reproduction number $R$, the effective severity $ES$ of the virus variant, and assumptions regarding importation, transmissibility, ICU capacity, and vaccine effectiveness.

##### A.2.4 End of the simulations

In our baseline case, we assume that an effective vaccine or treatment will become available after 9 months (270 days). As a pragmatic way of simulating this event, we set $\mathcal{R}_{\text{eff}} = 0.7$ and include a cool-down period of 150 days to ensure that the full health care costs of individuals infected during the main simulation period are included in the optimization. This is necessary, as many severe outcomes associated with substantial health costs occur several weeks after infection.

| Equation | Variable | Description<br>The number of (...) [stock] |
|---|---|---|
| (1) | $S^i$ | susceptible individuals |
| (2) | $E_a^i$ | exposed, will become asymptomatic, individuals |
| (3) | $E_s^i$ | exposed, will become symptomatic, individuals |
| (4) | $P^i$ | presymptomatic infectious individuals |
| (5) | $A^i$ | asymptomatic infectious individuals |
| (6) | $I^i$ | symptomatic infectious individuals |
| (7) | $H^i$ | hospitalised individuals |
| (8) | $ICU_1^i$ | individuals in hospital, will go to ICU |
| (9) | $ICU_2^i$ | individuals in ICU |
| (10) | $ICU_3^i$ | individuals in post-ICU care |
| (11) | $R^i$ | recovered and immune individuals |
| (12) | $D^i$ | dead individuals |

Table S1: Description of the compartments in the meta-population model

| Variable | Description | Value |
|---|---|---|
| $\beta(t)$ | Overall transmissibility (contact rate) | Set to corresponding $\mathcal{R}$ |
| $inf_s$ | Symptomatic infectiousness | 1 |
| $inf_a$ | Asymptomatic infectiousness | 0.1 |
| $inf_p$ | Presymptomatic infectiousness | 1.3 |
| $D_L$ | Latent period | 3 days |
| $D_P$ | Presymptomatic period | 2 days |
| $D_A$ | Duration asymptomatic | 4.8 days |
| $D_I$ | Duration symptomatic | 4.8 days |

Source: Based on [8] with small modifications for consistency with Norwegian data.

Table S2: Parameters in the meta-population model

| Variable | Description | Value | | | | | | | | |
|---|---|---|---|---|---|---|---|---|---|---|
| | | 0-9 | 10-19 | 20-29 | 30-39 | 40-49 | 50-59 | 60-69 | 70-79 | 80+ |
| | *Susceptibility:* | | | | | | | | | |
| $sus_a^i$ | asymptomatic | 1 | 1 | 1 | 1 | 1 | 1 | 1 | 1 | 1 |
| $sus_s^i$ | symptomatic | 1 | 1 | 1 | 1 | 1 | 1 | 1 | 1 | 1 |
| | *Fraction:* | | | | | | | | | |
| $f_a^i$ | asymptomatic | 0.47 | 0.47 | 0.32 | 0.32 | 0.32 | 0.32 | 0.32 | 0.32 | 0.32 |
| $f_s^i$ | symptomatic | 0.53 | 0.53 | 0.67 | 0.67 | 0.67 | 0.67 | 0.67 | 0.67 | 0.67 |
| | *Risk of:* | | | | | | | | | |
| $p_{hosp}^i$ | hospitalization† | 1.7e-3 | 3e-4 | 5.2e-4 | 8.2e-4 | 1.3e-3 | 2e-3 | 3e-3 | 1e-2 | 3.5e-2 |
| $p_{ICU}^i$ | ICU given hosp† | 0.04 | 0.1 | 0.1 | 0.1 | 0.1 | 0.18 | 0.18 | 0.1 | 0.05 |
| $p_{death}^i$ | death† | 1.8e-8 | 6.7e-8 | 3.4e-7 | 2.3e-6 | 1.5e-6 | 9.9e-5 | 5.6e-4 | 3.7e-3 | 3.7e-3 |
| | *Length of stay (days):* | | | | | | | | | |
| $D_H^i$ | in hospital§ | 1.8 | 3.5 | 3.6 | 3.4 | 4.7 | 5.5 | 6.4 | 6.2 | 6.4 |
| $D_{ICU_1}^i$ | in hospital pre-ICU§ | 1.4 | 1.4 | 1.4 | 2.8 | 3.7 | 3.1 | 3.9 | 4.5 | 4.5 |
| $D_{ICU_2}^i$ | in ICU§ | 3.3 | 3.3 | 3.3 | 3.3 | 9.3 | 9.5 | 16 | 15 | 14 |
| $D_{ICU_3}^i$ | in hospital post-ICU§ | 5 | 5 | 5 | 8.7 | 8.6 | 8.3 | 11 | 11 | 11 |

†Calibrated to ensure the correct number of hospitalizations and deaths during the Omicron wave in Norway see, section A.5.1.
§Estimated from Norwegian data during the Omicron Period (Jan-Oct 2022) with a similar approach as described in [9]

Table S3: Age varying parameters in the meta-population model for severity $ES = 1$.

#### A.3 Summary of empirical inputs

| Model component | Calibration target | Implementation and source |
|---|---|---|
| Epidemiological dynamics | Disease progression and contact patterns | Age-structured metapopulation SEIR model calibrated to Norwegian demographics, contact matrices and epidemiological parameters |
| Base severity ($ES = 1$) | First Omicron wave in vaccinated population | Age-specific hospitalization and mortality rates matched to observed outcomes in Norway. |
| High severity ($ES \approx 15$) | Original Wuhan-like variant in naive population | Age-specific hospitalization and mortality rates matched to observed outcomes in Norway. |
| Health losses | QALYs | Age-specific QALY losses from infection, hospitalization, ICU admission, post-acute sequelae and death based on supporting clinical evidence. |
| Value of health | Monetary value of a QALY | Baseline 1.4 million NOK/QALY (Norwegian VSLY guidance); Sensitivity analysis uses 0.5, 1 and 2 $\times$ baseline. |
| Maximum transmission reduction | Contact reduction | Combined NPIs, TTIQ and VSD reduce contacts by up to 90%, based on Norwegian and Danish evidence. |
| Intervention costs | Economic and welfare costs | GDP losses calibrated to the Norwegian GDP gap (winter 2020–21) plus tax-financing costs; welfare costs from Swedish WTP study. |
| Voluntary social distancing (VSD) | Individuals' behavioral response | Individuals trade-off expected health losses against welfare costs. Parameters reproduce roughly 50% lower peak incidence and 75% lower peak hospitalization without NPIs for $ES = 15$. |
| TTIQ | Cost and effectiveness | Costs based on Norwegian municipal expenditures, including fixed, variable and isolation-related costs. |
| Hospital overcapacity | Capacity-dependent mortality | Hospital model captures postponed treatment and increased mortality under capacity pressure based on empirical evidence. |
| Sickness and quarantine costs | Production, welfare and human-capital losses | Production losses based on Norwegian national accounts during COVID-19; welfare loss based on WTP estimates; school-related human-capital losses from earnings estimates. |
| Vaccination | Timing and effectiveness | Baseline vaccination after 270 days; Sensitivity analysis varies timing (270–1500 days) and effectiveness. |
| Parameter scan | Robustness | Varies transmissibility ($R$), effective severity ($ES$), QALY valuation, vaccination timing and hospital-overcapacity costs, yielding 4 320 parameter combinations. |

Table S4: Overview of main calibration targets and empirical inputs

#### A.4 Running the optimization

The optimal strategy, the $G(t)$-path, is the government choice of contact reduction through Test-Trace-Isolate-Quarantine (TTIQ) and NPIs that minimize societal costs from the pandemic in terms of health, economic and other welfare effects. The optimization presents several challenges due to a large number of parameters, non-convexity of the cost function, the need for derivative-free optimization and the comparatively slow run-time of the model.

The primary numerical difficulty lies in the possibility that $G$ can be different for each day $t$ of the simulation which gives a large number of parameters combined with the fairly slow (~0.1-0.5s on one CPU core) run-time of each simulation. In addition, due to the implementation details we are not able to calculate derivatives of the cost function analytically or through automatic differentiation. We therefore need to either use a derivative-free algorithm or calculate derivatives using finite difference methods. In the testing phase, both of these approaches required similar numbers of function calls to find a minimum, both of which are significantly higher than if we could use a gradient descent-type algorithm with pre-calculated derivatives. The cost function is also not convex and we are searching for the global minimum. Therefore, we use multiple parameterizations, algorithms, and starting points to reduce the risk of selecting a poor local minimum.

Because optimizing a separate intervention level for every day is computationally infeasible, we parameterize using several lower-dimensional representations that allow us to find the optimal solution. To this end, we choose different parameterizations including regularly spaced change points where $G(t)$ changes values and methods with fewer change-points but where the location of the change-point is also optimised.

To find the optimal solution in a computationally efficient way for all the combinations in each scenario, we follow the following procedure, where all of the minimization algorithms are from the NLopt package [10] as implemented in NLoptr [11].

For the methods with regular change points, which primarily capture *Suppression* and *No intervention* strategies, we optimize with multiple start points. One of these start-points is determined by first running a global optimization using the DIRECT-algorithm [12]. The other start points correspond to different types of strategies. From these starting points we use either the SBPLX [10] or the BOBYQA [13] to find the optimal $G(t)$. The separate optimizations are then defined by:

- Starting with a global optimization, then continuing with a local BOBYQA-optimization using 15 and 25 change points.
- Local optimization using BOBYQA starting from 'no interventions' using 15 change points.
- Local optimization using BOBYQA starting from constant $\beta$ which would give $R = 1$ using 15 change points.

To capture the *delayed containment*-strategies we also consider the following three optimizations:

- Local optimization using SBPLX starting from a 'delayed containment'-like solution. To determine the starting point we use the incidence from a run with 'no-interventions' for the same scenario. We then equally space 15 change points between $t_1$ and $t_2$ where $t_1$ is given as the minimum value of $t$ where $inc(t_1) = 0.95 \max(inc)$ and $t_2$ is given as the maximum value where $inc(t_2) = 0.25 \max(inc)$.
- Minimization using 2,3 or 4 change points, where the location of the change points is also optimized with both an SBPLX-algorithm and a BOBYQA-alorithm. In total we have $2n + 1$ variables for $n$ change points.

- Minimization using grid search to find days $t_1$ and $t_2$ such that for $t < t_1, G(t) = 0$ and for $t_1 < t < t_2, G(t) = 1$ and after $t_2$ we set $G(t) = 0.25$. In scenarios with no TTIQ we set the final value to 0 instead. We only consider values for $t_1$ with 20 days of where the peak would have been if there were no interventions and $7 < t_2 - t_1 < 40$.

The reported optimum is the lowest-cost solution obtained across these procedures. The above procedure compares different parameterizations of $G(t)$, and not just different starting points for the algorithm.

#### A.5 Modeling a new variant

Our framework characterizes a new respiratory virus variant by its initial reproduction number and effective severity. In the simulations we use 19 values of R from 1.2 to 3, and 20 values of ES from 1 to 20, in total 380 $(R, ES)$-combinations in each scenario, see Table S5.

| Parameters: | | Values: |
|---|---|---|
| Initial reproduction number | $R$ | $[1.2, 3]$ |
| Effective severity | $ES$ | $[1, 20]$ |

Table S5: Interval for $(R, ES)$-combinations of the virus

For a new variant to spread rapidly, it would need to be significantly immune evasive. Therefore, we assume that the entire population has the same level of immunity and that the biological properties of the virus, along with this level of immunity, result in an initial reproduction number, $R$. We then fix the initial reproduction number by choosing the contact rate, i.e., the value of the $\beta$-parameter which we interpret as the level of contact reducing interventions and behavior. Reproduction numbers are estimated at the start of the simulation by using the next-generation-matrix-method. Note that the implementation of voluntary social distancing (VSD), described in section C, reduces the contact-rate at the start of the simulations in the baseline case.[3] Hence, the initial reproduction numbers are defined conditional on this behavioral response.

##### A.5.1 Effective severity ($ES$)

The effective severity, $ES$, of a new variant is a combination of the biological properties of the virus and the immunity in the population. When calibrating the effective severity we adjust both the transition probability at each disease stage, time in hospital and the lost QALYs for symptomatic, hospitalized, ICU and post acute sequale (for the latter, see section B.1). Hence, we introduce severity-dependent multipliers given in Table S6 and Table S7.

**Calibrate time in hospital and transition probabilities.** We calibrate the effective severity of a new variant such that severity $ES = 1$ approximately corresponds to the first Omicron wave in Norway, when a large majority of the population was vaccinated and/or previously infected.[4] Based on an overall assessment of available data we assume that approximately 3 million out of a population of 5.4 million was infected during the Omicron wave, see e.g [15]. The infection fatality rate (IFR) and infection hospitalization rate (IHR) are calibrated such that, for reproduction number $R = 1.6$ and effective severity $ES = 1$, the model reproduces the observed age-specific mortality and hospitalization rates from the Omicron wave, conditional on the infection share during Omicron. When $ES$ increases, the severity parameters increase as in equation (18)-(21):

$$p^i_{hosp}(ES) = ES \times p^i_{hosp} \tag{18}$$

$$p^i_{death}(ES) = ES \times p^i_{death} \tag{19}$$

$$p^i_{ICU}(ES) = \min\left(1 + \frac{2.8(ES-1)}{15}, 3.8\right) p^i_{ICU} \tag{20}$$

$$D^i_H(ES) = \min\left(1 + \frac{1.3(ES-1)}{15}, 2.3\right) D^i_H \tag{21}$$

[3] In section C.2 we compare baseline with no VSD.

[4] This corresponds to the parameter values in Table S3, where we have calibrated the IHR and IFR with data from the NIPH's risk assessment dated April 29th 2022 [14].

These equations are parameterized such that for an effective severity $ES = 15$, the probability of hospitalization $p^i_{hosp}$ and death $p^i_{death}$ increase 15-fold while ICU-probability $p^i_{ICU}$ increases 3.8-fold and the length of stay in hospital $D^i_H$ increases 2.3-fold. This is an assumption based on the difference in these parameters between the Delta period and the Omicron period from Norwegian Covid-19 data [9]. The difference in ICU-probability is based on the observed change in hospitalizations during the pandemic, from un-vaccinated Delta admissions compared to all Omicron admissions.

| | $D^i_H$ | $p^i_{hosp}$ | $p^i_{ICU}$ | $p^i_{death}$ |
|---|---|---|---|---|
| Severity multipliers from severity level $ES = 1$ to $ES = 15$ | 2.3 | 15 | 3.8 | 15 |

Table S6: Severity multipliers for time in hospital and transition probabilities

Comparing the estimate of the IHR and IFR for Omicron, with estimates from the early phase of the pandemic [16], we find that a severity of approximately 15 describes the original Wuhan variant in an immunologically naive population in Norway. The majority of the reduction in severity is due to improved immunity from vaccination and previous infections.

**Calibrate the QALY loss.** The QALY loss by age group, illness duration, and post-acute sequelae are drawn from research on infections with the Delta variant (autumn 2021) and the first Omicron variant (winter 2022), for both immune and non-immune individuals, see section B.1.1. These estimates were initially produced for the expert commission [17], advising Norwegian authorities during the pandemic. We use these QALY estimates to calibrate our severity scale.

The most severe cases of Covid-19 in Norway were among unvaccinated patients infected with the Delta variant, which we use to calibrate the QALY loss for severity level $ES \geq 15$. The least severe cases occurred among the vaccinated population infected with Omicron in 2022, calibrated to $ES = 1$. The scaling from $ES = 1$ to higher severity levels is shown in Table S7. The multiplicative scale is at its maximum when $ES = 15$, for severity above 15 we assume that the QALY loss remains the same as for severity=15, and only $p^i_{hosp}$, $p^i_{death}$ increase further. Figure S1 illustrates the expected QALY loss per age group for different severity levels.

| | Symptomatic $Q^I_i$ | Hospitalized $Q^H_i$ | ICU $Q^{ICU}_i$ | Death $Q^D_i$ |
|---|---|---|---|---|
| Severity multipliers from severity level $ES = 1$ to $ES = 15$ | 5 | 3 | 1.5 | 1 |

Note: The variables $Q^I_i, Q^H_i, Q^{ICU}_i$ and $Q^D_i$ refer to the total QALYs per person in each age group including long-term effects (post acute sequelae), summarized in Table S8 for an outcome when $ES = 1$.

Table S7: Severity multipliers for the QALY losses

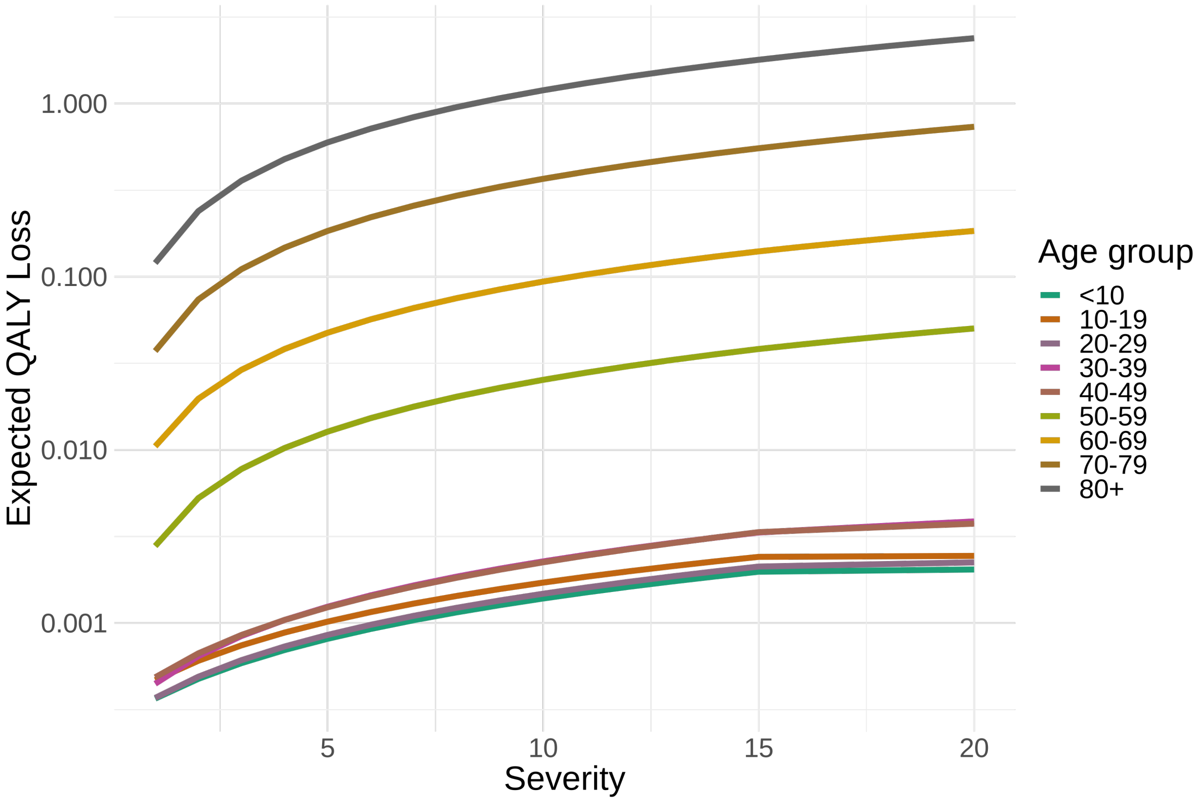

Figure S1: Expected QALY loss per infection per age group for different levels of severity. Note: log scale

### B Calibrated costs

The model is designed to capture the main social costs from a new pandemic in terms of health, economy and other welfare effects. The model identifies four main channels through which a pandemic generates social costs, as illustrated below. The "optimal strategy" is the trajectory of government interventions over time that minimizes the sum of these costs.

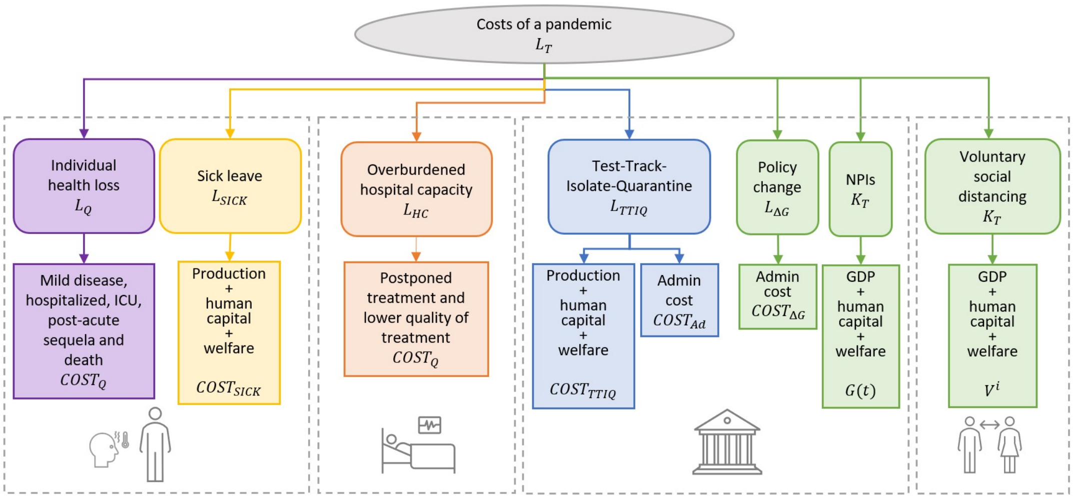

[1.] **Costs of Infections:** This includes the direct health losses associated with symptomatic infection, hospitalization, post-acute sequelae, and death, measured in Quality Adjusted Life Years (QALYs) and monetized. Sickness also results in welfare losses and age-specific economic loss, such as lost productivity during sick leave (ages 0-9 and 20-69) or lost educational opportunities (ages 9-19).

[2.] **Costs of an Overburdened Health System:** An increase in hospitalized Covid-19 patients can lead to postponed elective treatments in general wards, delayed surgeries due to limited ICU capacity, and reduced quality of care, leading to higher morbidity for both Covid-19 and non-Covid-19 patients in intensive care, all measured in QALYs and monetized.

[3.] **Costs of interventions:** Government's TTIQ-regime involves administrative costs for contact tracing and testing, while isolating and quarantining asymptomatic individuals lead to welfare losses and age-dependent economic impacts. Government NPIs result in reduced economic activity, tax financing costs, and decreased welfare.

[4.] **Costs of Voluntary Social Distancing:** When individuals alter their behavior to avoid infection, this leads to similar costs as government NPIs, including reduced economic activity, tax financing costs, and diminished welfare.

$$L_T = K_T + L_{\Delta G} + L_Q + L_{HC} + L_{SICK} + L_{TTIQ} \quad (22)$$

The illustration above corresponds to equation (22). In the following we describe each cost type in detail:

| | | Equation | Section |
|---|---|---|---|
| $K_T$ | Economic cost of contact reduction: NPIs and voluntary social distancing | (29) | B.3 |
| $L_{\Delta G}$ | Economic cost of changing in the intervention level | (30) | B.3.1 |
| $L_Q$ | Health losses from COVID-19 (lost QALYs and life years) | (23) | B.1 |
| $L_{HC}$ | Losses from hospitals operating above capacity (lost QALYs and life years) | (26) | B.2 |
| $L_{SICK}$ | Economic and welfare cost of staying at home when sick, including the lost production when on sick leave | (34) | B.5 |
| $L_{TTIQ}$ | Costs of the TTIQ system (administrative costs of running the program and the economic and welfare costs associated with quarantine and isolation) | (32) | B.4 |

#### B.1 Individual health loss from infection

The health loss from new respiratory viruses will depend on the characteristics of the variant. Since the course of illness for future variants is unknown, our estimates are based on experiences with previous SARS-CoV-2 variants. We assume the same age-severity profile as observed with Covid-19 so far. The total value of the health loss due to infection is given by equation (23), where $f_s^i$ represents the fraction of symptomatic infections in age group $i$, given in Table S3. The variables $\{\tilde{I}_i, \tilde{H}_i, I\tilde{C}U_i, \tilde{D}_i\}$ denote the total number of individuals per age group at the end of the simulation for the four outcomes: infections, hospitalizations, ICU and deaths. The variables $\{Q_i^I, Q_i^H, Q_i^{ICU}, Q_i^D\}$ refer to the total QALY loss per person in each age group, including long-term effects (post acute sequelae), summarized in Table S8 for an outcome when $ES = 1$.

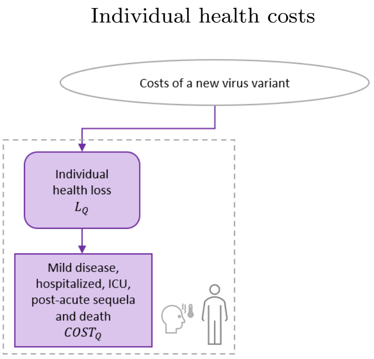

$$L_Q(\mathcal{S}(t)) = COST_Q \sum_i \Big( (f_s^i \tilde{I}_i - \tilde{H}_i - \tilde{D}_i) \times Q_i^I + (\tilde{H}_i - I\tilde{C}U_i) \times Q_i^H + I\tilde{C}U_i \times Q_i^{ICU} + \tilde{D}_i \times Q_i^D \Big), \tag{23}$$

| | Total QALYs, including post sequelae | | | |
|---|---|---|---|---|
| **Age group** | **Symptomatic** $Q_i^I$ | **hospitalization** $Q_i^H$ | **ICU** $Q_i^{ICU}$ | **Death** $Q_i^D$ |
| 0-9 | 0.0008 | 0.0031 | 0.0077 | 66.5 |
| 10-19 | 0.001 | 0.0035 | 0.012 | 57.3 |
| 20-29 | 0.0011 | 0.0046 | 0.0215 | 48.3 |
| 30-39 | 0.0011 | 0.0048 | 0.0246 | 39.6 |
| 40-49 | 0.0013 | 0.0054 | 0.0389 | 31.3 |
| 50-59 | 0.0014 | 0.006 | 0.0395 | 23.5 |
| 60-69 | 0.0087 | 0.0082 | 0.0441 | 15.6 |
| 70-79 | 0.0044 | 0.0111 | 0.0476 | 9.8 |
| 80+ | 0.006 | 0.013 | 0.0361 | 4.9 |

Table S8: Example of the health loss for the infected per model outcome for severity $ES = 1$

**Monetary value:** In order to monetize the health loss, we assume a value of $COST_Q = 1.4$mnNOK per QALY.[5] However, there is no objective valuation of health, and the society's willingness to pay for saving a quality adjusted life year may be higher or lower. Thus, we also consider alternative monetary values of a QALY, cf. Figure 4 in the main paper.

[5]Since the Norwegian health sector does not monetize QALYs, we use the recommended monetary value of health impacts, as suggested by [18], for use in Cost-Benefit Analyses for sectors outside of healthcare. The recommended monetary value of a Statistical Life Year (VSLY) was 1.4 mnNOK in 2021. The VSLY is derived from Norway's official Value of a Statistical Life (VSL), which was 40.35 mnNOK in 2021 [19]. [18] derive the VSLY from the VSL using the formula: $VSL = \sum_{t=0}^{40} \frac{VSLY}{(1+r)^t}$. The official guidance recommends calculating an interval by considering two discount rates: $r = 0$ and $r = 0.031$, and then taking the average of these values. This approach results in a VSLY of 1.39 mnNOK in 2021.

##### B.1.1 Lost QALYs

The direct individual health loss from a new virus variant is determined by reduced life quality for those who fall ill and the loss of life expectancy for those who die. We measure both reduced life quality and lost life expectancy in the health unit QALY (Quality Adjusted Life Years), where QALY=1 equates to one year in perfect health (no loss of QoL). The simulation provides the number of individuals per age group per day per disease stage, from which we can calculate the QALY loss for the population per model outcome.

| Age group | Expected remaining lifetime | QALYs |
|---|---|---|
| 0-9 | 78.17 | 66.5 |
| 10-19 | 68.22 | 57.3 |
| 20-29 | 58.45 | 48.3 |
| 30-39 | 48.69 | 39.6 |
| 40-49 | 39.01 | 31.3 |
| 50-59 | 29.59 | 23.5 |
| 60-69 | 20.77 | 15.6 |
| 70-79 | 12.91 | 9.8 |
| 80+ | 6.67 | 4.9 |

Source: [17]

Table S9: Death

Our QALY estimates in Table S10–S13 are taken from [17] and reflect data from spring 2022. Since then, knowledge about the impact of COVID-19 variants on health-related quality of life and illness duration, including long-term effects, has increased. While the estimates for the duration of long-term effects—especially those not identified at that time — may not precisely match a previous SARS-CoV-2 variant, we believe this uncertainty is acceptable for our purpose. In our analysis these estimates are being used to calibrate projections for possible future virus variants, which will inherently carry more uncertainty, not necessarily related to SARS-CoV-2.

It's important to note that the health-related quality of life losses and durations included in our QALY calculations are average estimates. If a smaller portion of the population (e.g., 5-10%) experiences significantly longer durations of impaired health, such distributional effects will not be captured by these average estimates.

**Mild illness (not hospitalised).** Table S10 outlines the duration (in days) and QALYs per day for mild symptomatic illness at two severity levels: severity level $ES = 1$, calibrated from data on the first Omicron variant in a vaccinated population, and severity level $ES = 15$, based on data from the Delta variant in an unvaccinated population.

| | Omicron in a vaccinated population $ES = 1$ | | Delta in an unvaccinated population $ES = 15$ | |
|---|---|---|---|---|
| Age group | Days | QALY/ day | Days | QALY/ day |
| 0-9 | 2 | 0.1 | 5 | 0.20 |
| 10-19 | 3 | 0.1 | 5 | 0.25 |
| 20-29 | 3 | 0.1 | 5 | 0.25 |
| 30-39 | 3 | 0.1 | 6 | 0.25 |
| 40-49 | 4 | 0.1 | 7 | 0.30 |
| 50-59 | 4 | 0.1 | 9 | 0.30 |
| 60-69 | 4.5 | 0.15 | 11 | 0.30 |
| 70-79 | 5 | 0.2 | 14 | 0.35 |
| 80+ | 5 | 0.25 | 14 | 0.35 |

Source: [17]

Table S10: QALY loss for symptomatic illness (not hospitalised) per age group

For mild illness, we assume symptoms typically resolve within one to two weeks. Recovery time is influenced by age, with older individuals generally taking longer to recover. According to The Zoe Symptom Study, one-third of individuals experienced symptom resolution within three days or less. Symptoms from variants like Omicron typically resolved faster than those from Alpha and Delta variants [20]. Fully vaccinated individuals generally experience milder symptoms and shorter recovery times. By comparison, QALY loss for mild illness from swine flu was estimated at 0.3 for non-hospitalized individuals [21] and about 0.5 for hospitalized cases [22], on a 0-1 scale

**Hospitalised.** Table S11 presents the duration (in days) and the daily QALY loss (on a 0-1 scale) for hospitalized patients at two severity levels, $ES = 1$ and $ES = 15$, calibrated based on the experience with Omicron in a vaccinated population and the Delta variant in an unvaccinated population. We assume that the duration and quality of life loss in the pre-hospitalization and post-discharge phases align with the course of mild illness described in Table S10.

For estimating hospital stay durations by age group, an early study by [23] found that the median length of hospital stays ranged from 5 to 29 days. In the UK, the median hospital stay for Covid-19 patients over the entire pandemic was 7 days, with some fluctuation. Preliminary studies indicate that hospital stays increase with age. [24] also reported a median hospital stay of 7 to 8 days. Norwegian data showed slightly shorter hospital stays [25]. Research indicates that hospitalization durations were approximately 50% shorter for Omicron compared to the Delta variant [26–28].

| | 1. Pre admission | | 2. Hospital stay | | 3. Post discharge | |
|---|---|---|---|---|---|---|
| Age group | Days | QALY/ day | Days | QALY/ day | Days | QALY/ day |
| | Omicron in a vaccinated population $ES = 1$: | | | | | |
| 0-9 | 2 | 0.1 | 1.5 | 0.3 | 2 | 0.1 |
| 10-19 | 3 | 0.1 | 1.5 | 0.3 | 3 | 0.1 |
| 20-29 | 3 | 0.1 | 2.0 | 0.3 | 3 | 0.1 |
| 30-39 | 3 | 0.1 | 2.0 | 0.3 | 3 | 0.1 |
| 40-49 | 4 | 0.1 | 2.0 | 0.4 | 4 | 0.1 |
| 50-59 | 4 | 0.1 | 2.5 | 0.4 | 4 | 0.1 |
| 60-69 | 4.5 | 0.15 | 2.5 | 0.4 | 4.5 | 0.15 |
| 70-79 | 5 | 0.2 | 3.0 | 0.4 | 5 | 0.2 |
| 80+ | 5 | 0.25 | 3.0 | 0.4 | 5 | 0.25 |
| | Delta in an unvaccinated population $ES = 15$: | | | | | |
| 0-9 | 4 | 0.20 | 3 | 0.4 | 4 | 0.20 |
| 10-19 | 5 | 0.25 | 3 | 0.4 | 5 | 0.25 |
| 20-29 | 6 | 0.25 | 4 | 0.4 | 6 | 0.25 |
| 30-39 | 7 | 0.25 | 4 | 0.4 | 7 | 0.25 |
| 40-49 | 8 | 0.30 | 4 | 0.5 | 8 | 0.30 |
| 50-59 | 8 | 0.30 | 5 | 0.5 | 8 | 0.30 |
| 60-69 | 8 | 0.30 | 5 | 0.5 | 8 | 0.30 |
| 70-79 | 8 | 0.35 | 6 | 0.5 | 8 | 0.35 |
| 80+ | 6 | 0.35 | 6 | 0.5 | 6 | 0.35 |

Source: [17]

Table S11: QALY loss when hospitalized (general ward) per age group

**Admitted to the Intensive Care Unit (ICU).** Table S12 presents the duration (in days) and daily QALY loss (on a 0-1 scale) for patients admitted to the ICU at two severity levels: $ES = 1$, calibrated from the experience with Omicron in a vaccinated population, and $ES = 15$, based on data from the Delta variant in an unvaccinated population.

[24] suggested a median time of approximately 2 days from hospital admission to ICU entry. Norwegian data indicated that the length of hospital stay increases with age [25], and that ICU stay durations were about 2 days shorter for vaccinated individuals compared to the unvaccinated [25]. Additionally, [24] reported a median ICU stay of around 12 days. [29] suggested that ICU stays were shorter during the Omicron wave compared to Delta. For a variant similar to Omicron, we estimate hospital stays to be approximately 30% shorter than for a Delta-like variant.

| | 1. Pre admission | | 2. Pre ICU | | 3. ICU | | 4. post ICU | | 5. Post discharge | |
|---|---|---|---|---|---|---|---|---|---|---|
| Age group | Days | QALY/ day | Days | QALY/ day | Days | QALY/ day | Days | QALY/ day | Days | QALY/ day |
| | Omicron in a vaccinated population $ES = 1$: | | | | | | | | | |
| 0-9 | 2 | 0.20 | 1 | 0.4 | 1 | 0.6 | 1 | 0.4 | 2 | 0.20 |
| 10-19 | 3 | 0.25 | 1 | 0.4 | 1 | 0.6 | 1 | 0.4 | 3 | 0.25 |
| 20-29 | 3 | 0.25 | 2 | 0.4 | 2 | 0.6 | 6 | 0.4 | 4 | 0.25 |
| 30-39 | 3 | 0.25 | 2 | 0.4 | 5 | 0.6 | 3 | 0.4 | 5 | 0.25 |
| 40-49 | 4 | 0.30 | 2 | 0.5 | 9 | 0.7 | 6 | 0.5 | 6 | 0.30 |
| 50-59 | 4 | 0.30 | 3 | 0.5 | 8 | 0.7 | 6 | 0.5 | 6 | 0.30 |
| 60-69 | 4.5 | 0.30 | 3 | 0.5 | 9 | 0.7 | 6 | 0.5 | 6 | 0.30 |
| 70-79 | 5 | 0.35 | 3 | 0.5 | 8 | 0.7 | 6 | 0.5 | 6 | 0.35 |
| 80+ | 5 | 0.35 | 3 | 0.5 | 5 | 0.7 | 1 | 0.5 | 4 | 0.35 |
| | Delta in an unvaccinated population $ES = 15$: | | | | | | | | | |
| 0-9 | 4 | 0.20 | 1 | 0.4 | 1 | 0.6 | 2 | 0.4 | 4 | 0.20 |
| 10-19 | 5 | 0.25 | 1 | 0.4 | 1 | 0.6 | 2 | 0.4 | 5 | 0.25 |
| 20-29 | 6 | 0.25 | 2 | 0.4 | 3 | 0.6 | 4 | 0.4 | 6 | 0.25 |
| 30-39 | 7 | 0.25 | 2 | 0.4 | 7 | 0.6 | 4 | 0.4 | 7 | 0.25 |
| 40-49 | 8 | 0.30 | 2 | 0.5 | 13 | 0.7 | 7 | 0.5 | 8 | 0.30 |
| 50-59 | 8 | 0.30 | 3 | 0.5 | 12 | 0.7 | 7 | 0.5 | 8 | 0.30 |
| 60-69 | 8 | 0.30 | 3 | 0.5 | 14 | 0.7 | 6 | 0.5 | 8 | 0.3 |
| 70-79 | 8 | 0.35 | 3 | 0.5 | 12 | 0.7 | 6 | 0.5 | 8 | 0.35 |
| 80+ | 6 | 0.35 | 3 | 0.5 | 7 | 0.7 | 1 | 0.5 | 8 | 0.35 |

Source: [17]

Table S12: QALY loss for admissions to the intensive care unit (ICU) per age group

**Post-acute sequelae ("long covid").** Table S13 outlines the assumptions regarding long-term sequelae following mild illness, hospitalization, or ICU treatment for two severity levels: $ES = 1$, based on Omicron in a vaccinated population, and $ES = 15$, calibrated from the Delta variant in an unvaccinated population. For each health state — mild illness, hospitalization, or ICU treatment — we specify the percentage affected, the duration (in days), and the daily QALY loss (on a 0-1 scale) by age group.

We assume that long-term effects increase with age and disease severity, meaning the highest proportion of post-acute sequelae occurs in the oldest age groups and among the most severely ill. Symptom duration is assumed to taper off after 3 to 5 months for adults, with a shorter duration for younger individuals.

| | After mild illness | | | After hospitalization | | | After ICU | | |
|---|---|---|---|---|---|---|---|---|---|
| Age group | Share | Days | QALY/ day | Share | Days | QALY/ day | Share | Days | QALY/ day |
| | Omicron in a vaccinated population $ES = 1$: | | | | | | | | |
| 0-9 | 4% | 40 | 0.05 | 4% | 70 | 0.1 | 5% | 80 | 0.2 |
| 10-19 | 4% | 30 | 0.05 | 4% | 60 | 0.1 | 8% | 80 | 0.3 |
| 20-29 | 5% | 40 | 0.05 | 8% | 60 | 0.1 | 9% | 80 | 0.3 |
| 30-39 | 5% | 40 | 0.05 | 8% | 70 | 0.1 | 9% | 90 | 0.3 |
| 40-49 | 4% | 45 | 0.05 | 5% | 75 | 0.1 | 6% | 95 | 0.3 |
| 50-59 | 4% | 50 | 0.05 | 5% | 80 | 0.1 | 7% | 100 | 0.3 |
| 60-69 | 5% | 60 | 0.10 | 6% | 90 | 0.12 | 7% | 120 | 0.3 |
| 70-79 | 6% | 70 | 0.10 | 7% | 100 | 0.12 | 8% | 130 | 0.4 |
| 80+ | 7% | 80 | 0.12 | 8% | 110 | 0.12 | 9% | 140 | 0.4 |
| | Delta in an unvaccinated population $ES = 15$: | | | | | | | | |
| 0-9 | 10% | 40 | 0.1 | 10% | 70 | 0.2 | 15% | 80 | 0.2 |
| 10-19 | 10% | 30 | 0.1 | 10% | 60 | 0.2 | 20% | 80 | 0.3 |
| 20-29 | 15% | 40 | 0.1 | 20% | 60 | 0.2 | 25% | 80 | 0.3 |
| 30-39 | 15% | 40 | 0.1 | 20% | 70 | 0.2 | 25% | 90 | 0.3 |
| 40-49 | 20% | 45 | 0.1 | 25% | 75 | 0.2 | 30% | 95 | 0.3 |
| 50-59 | 20% | 50 | 0.1 | 25% | 80 | 0.2 | 35% | 100 | 0.3 |
| 60-69 | 25% | 60 | 0.15 | 30% | 90 | 0.25 | 35% | 120 | 0.3 |
| 70-79 | 30% | 70 | 0.2 | 35% | 100 | 0.3 | 40% | 130 | 0.4 |
| 80+ | 35% | 80 | 0.25 | 40% | 110 | 0.35 | 45% | 140 | 0.4 |

Source: [17], including the following discussion of the literature available:
**Delta variant: Share**
*Age 0-29:* [30] conclude that COVID-19 had a limited impact on healthcare services for children and adolescents in Norway. Preschool-aged children tend to recover more slowly (3-6 months) than primary and secondary students (1-3 months), often due to respiratory issues. [31] found that about a quarter of hospitalized children experience long-term symptoms. For SARS-CoV-2 age is strongly linked to the risk of admission and death. Since long-term effects are associated with disease severity, fewer severe cases in younger groups likely mean fewer long-term effects.
*Age 30-59:* Age is one of the strongest predictors of disease severity, which increases the need for intensive treatment and the risk of long-term consequences. Hence, we assume a gradient, with long-term effects increasing with age and disease severity.
*Age 60-80+:* The highest proportion of long-term effects is expected among the oldest and most severely affected individuals. Post-intensive care syndrome shares similarities with long-COVID, where severity, ICU stay, and pre-existing conditions are predictors of survival. Although initially common, studies suggest that these impacts decrease over time.
**Delta variant: Days and QALYs**
*After mild illness:* [32] found no significant difference in QoL for non-hospitalized patients 1.5 to 6 months after a positive PCR test. Age correlates with symptom severity and duration, which likely results in a longer impact on QoL for older individuals. Symptoms are expected to subside after 3 to 6 months, with faster recovery for younger people. [33] found no increased healthcare use after 2 months. [32] reported QoL was slightly reduced but comparable to the general population (around 0.82/1). [34] found similar QoL for hospitalized patients and controls after 12 months.
*After hospitalization:* [35] and [36] found a greater impact on QoL in hospitalized patients (0.72 and 0.86), compared to[32] (0.82). [33] found little or no increase in healthcare use 4 to 6 months after mild or severe Covid-19, suggesting long-term symptoms are rare beyond 3 months.
*After ICU:* [35] and [36] observed the largest QoL reduction in ICU patients (0.61 and 0.82). Based on a review of long-term effects, many hospitalized patients had symptoms lasting beyond 6 months. [33] also reported little to no increase in healthcare use 4 to 6 months after severe Covid-19, including ICU cases.
**Omicron: Share, Days and QALYs** At the time of [17], no studies had been published on the long-term effects of Omicron infections. Based on [37] we assumed that the risk of long-term effects decreased by 50% for individuals aged 0-39 and by 60% for those over 40 who were vaccinated during the delta wave compared to the unvaccinated. With no data on Omicron, we projected a 25% reduction in risk for individuals aged 0-39 and a 50% reduction for those over 40 following Omicron infections, compared to vaccinated individuals infected with Delta. Given the lower symptom burden associated with Omicron, we also assumed a 50% reduction in the QoL (Quality of Life) impact for both mild and serious cases compared to Delta but kept the duration constant.

Table S13: QALY loss for post-acute sequelae per age group

#### B.2 Cost of overburdened hospitals

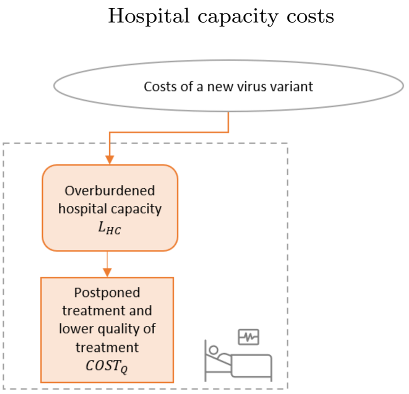

We have developed a simple hospital model to calibrate the consequences of overburdened hospitals and ICUs during the pandemic, and how long elective procedures need to be postponed as a result. Let $B$ denote the maximum number of beds available under normal conditions, for the general ward or at the ICU (we use the same model and notation for the general ward and the ICU). For simplicity, we assume that each day there is a constant number of new non-Covid-19 related admissions, including elective admissions, $EL$, and acute admissions, $AC$. Acute admissions will proceed regardless of capacity, but elective admissions will be accepted only if there is available capacity. The non-Covid-19 admissions are assumed to have an average length of stay of $D_{EL}$ for elective admissions and $D_{AC}$ for acute. Let $\Psi_{C19}$ denote the prevalence of Covid-19 admissions in either hospital or ICU taken from the model above. The available capacity for elective admissions, $Z$, can now be defined by the following equation:

$$Z = B - AC \times D_{AC} - \Psi_{C19} \tag{24}$$

During a pandemic, when the prevalence of acute admissions of Covid-19 and non-Covid-19 patients exceeds capacity, elective admissions will be postponed until the hospital prevalence returns to normal levels. This postponement will result in a health cost for patients who need to wait longer for healthcare.

We track the number of postponed elective admissions daily during the epidemic simulation and admit $Z/D_{EL}$ new patients for elective procedures daily. We then assign a cost in Quality-Adjusted Life Years (QALYs) per day for each day of postponed treatment for the elective patients. This cost is set at $\Omega^{EL} = 0.05$ QALYs per year of postponement patients at the general ward and $\Omega^{AC} = 0.2$ QALYs per year of postponement for ICU admissions.

For ICU admissions, we also consider the reduction in treatment quality when the ICU operates over capacity. At normal capacity, we assume a 20% mortality rate for patients with Covid-19 in ICUs, based on the study by [38]. Based on evidence for [39], we assume a linear effect where an increase in the number of patients equal to 1% of the capacity leads to an increase in mortality by 0.7% of the mortality at full capacity. For example, with a national capacity of 350 beds, treating 450 patients, i.e. 28.6% above capacity, would result in a $0.7\% \times 28.6 = 20\%$ increase in the mortality for all patients admitted during this period. We assume that this increase applies uniformly to Covid-19 and non-Covid-19 patients due to triage considerations.

Hence, each admission to the ICU when it is operating over capacity by $X$ percent is associated with an additional loss of life-years, given by equation (25):

$$\Phi(X) = 10.4(0.2 \times 0.007X) \tag{25}$$

where we assume that the average death in the ICU is equivalent to a loss of 10.4 QALYs, based on our assumption that mortality at the ICU follows a similar exponential increase with age as the overall Covid-19 mortality in Norway, weighted by the age distribution of those who have received ICU treatment for Covid-19 in Norway.[6,7] We also assume that after the total mortality rate at the ICU reaches 75%, it will not increase any further.

Putting this all together we then find that total cost for exceeding hospital capacity:

$$L_{HC} = COST_Q \left( \sigma_H \times \Omega^{EL} + \sigma_{ICU} \times \Omega^{AC} + \sum_t ICU_{inc}(t) \times \Phi\Big( \frac{ICU(t) + ICU_O - \overline{ICU})}{\overline{ICU}} \Big) \right), \quad (26)$$

Where $\sigma_H$ is the number of years of postponed hospital admissions, $\sigma_{ICU}$ is the number of years of postponed ICU treatment. $ICU_{inc}$ is the total incidence of new admissions to ICU including Covid-19 and other reasons. The function $\Phi(X)$ governs the reduction in quality, based on how much the current total prevalence, which is made up of the Covid-19 prevalence $ICU(t)$ and the prevalence from other conditions $ICU_O$, exceeds the capacity $\overline{ICU}$.

For our baseline case, we use the hospital parameters shown in Table S14.

| | Number of acute patients $AC$ | Number of elective patients $EL$ | Length of stay acute patients $D_{AC}$ | Length of stay elective patients $D_{EL}$ | Capacity: number of patients treated at once | QALY loss per year of postponed treatments |
|---|---|---|---|---|---|---|
| General ward | 1600 | 650 | 4 | 5 | 11 000 | $\Omega^{EL} = 0.05$ |
| ICU | 35 | 30 | 4 | 5 | 350 | $\Omega^{AC} = 0.2$ |

Table S14: Parameters for the hospital model

**Monetary value:** In order to monetize the health loss, we assume a baseline value of $COST_Q$ = 1.4mnNOK per QALY, cf. section B.1 above.

[6]Access public data at GitHub from the Norwegian Institute of Public Health (NIPH).
[7]See The Weekly Report from the Norwegian Institute of Public Health (NIPH), [only in Norwegian].

#### B.3 The cost of contact reduction

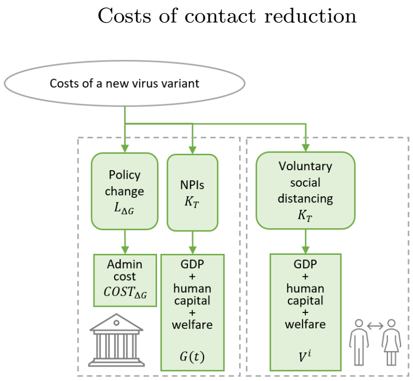

Interventions to contain the disease may involve large costs in the form of reduced economic activity and reduced welfare. As explained in the main paper, the costs from contact reduction from NPIs and social distancing is based on the reduction in mainland GDP, as measured by the output gap calculated by Statistics Norway, of 4.6%, corresponding to 12.7 bnNOK per month (approximately USD 1.2 bn). The tax financing costs of the interventions and fiscal support measures are estimated to correspond to an additional cost of 2 bnNOK per month, cf, [17]. This gives a total of about 15 bnNOK per month in economic costs.

Since our measure of economic costs is based on the reduction in GDP, it does not account for changes in the composition of activities across sectors and firms and within organizations. In cases where regular activities and planned investments were replaced by efforts to manage and adapt to the pandemic and interventions, this approach will underestimate the loss of output and welfare that would have been generated by those regular activities.

Contact reducing interventions will also involve a considerable welfare loss from reduced social activity and contact. As explained in the main paper, the calibration of the welfare loss is based on a choice experiment conducted by [40]. In this experiment, a number of Swedish individuals were asked about the monetary compensation they would require to be willing to participate in a 4-week stay-at home policy, with a maximum of 2 hours per week outside home. The estimated compensation per week was 2000 SEK (Swedish krona) (about USD 200), which corresponds to about 2 000 NOK per week. The welfare loss per person per day is then 285 NOK per day. We assume that this welfare loss applies for the maximum reduction in the contact rate of 90% from interventions and voluntary social distancing. For the whole population of 5.4 mn, including children, the total welfare costs per month is (2000/7)*(365/12)*5.4 $\approx$ 47 bnNOK per month. The total costs per month are thus 15 + 47 = 62 bnNOK per month.

We use a quadratic cost function to capture how the intervention costs depend on the intervention level such that for $G = V = 1$, we reach 15 + 47 = 62 bnNOK per month. To this end, we define an auxiliary function:

$$K(x) = 36.1x + 25.9x^2. \tag{27}$$

Note that $K(1) = 62$, equal to the costs of maximum interventions, while $K(0.5) = 24.5$, implying a convex cost function where 50% of the maximal reduction in the contact rate costs 40% of the total costs for the maximum reduction (as $24.5/62 \approx 0.4$). The overall cost from contact reduction for age group $i$ is:

$$K_T^i = \frac{1}{0.9} f^i \Big(0.8\big(K(V^i) + K(G^*)\big) - \frac{0.7}{62} K(V^i)K(G^*)\Big), \tag{28}$$

Here, $f^i$ is the fraction of the population in age group $i$. Since the cost of TTIQ is included separately we use $G^* = \max\big(0, (G - 0.25)/0.75\big)$ for the costs caused by NPIs. For details on the cost of TTIQ see section B.4 and for details on voluntary social distancing see equation (37) in section C.

The total costs from contact reduction measured in bnNOK is then given by:

$$K_T = \sum_{i=1}^{9} K_T^i. \tag{29}$$

##### B.3.1 Changing intervention levels

We assume that each adjustment of the intervention level is associated with a cost, given by equation (30):

$$L_{\Delta G} = \sum_i K_{change}(\Delta G_\tau), \tag{30}$$

where $\Delta G_\tau = G_{t_2} - G_{t_1}$ are all the non-zero changes in $G(t)$ where $G(t_2) \neq 0$ (we assume that there are no costs when all interventions are removed). The function $K_{change}(\Delta G_\tau)$ is given by equation (31):

$$K_{change}(\Delta G_\tau) = 2 \times COST_{\Delta G} \left( \frac{1}{1 + \exp(-5|\Delta G_\tau|)} - 0.5 \right), \tag{31}$$

where $COST_{\Delta G} = 3$ bnNOK. For large adjustments $G(t)$, this will tend to a maximum cost of 3 bnNOK. This additional cost accounts for expenses related to information, decision-making and implementation associated with changing interventions. Such costs prevent strategies that involve frequent changes in $G(t)$.

#### B.4 The costs of TTIQ

The effect of the Test-Trace-Isolate-Quarantine (TTIQ) system on the pandemic is assumed to come via the effect of $G(t)$ on the contact rate, as given by equation (15). The costs of the TTIQ system on day $t \in \{1, T\}$ depends on the capacity use of the system that day, as measured by the indicator $TTIQ(t) \in [0,1]$, where $TTIQ(t) = G(t)/0.25$ for $G(t) \leq 0.25$ and $TTIQ(t) = 1$ for $G(t) > 0.25$, and on the infection rate.

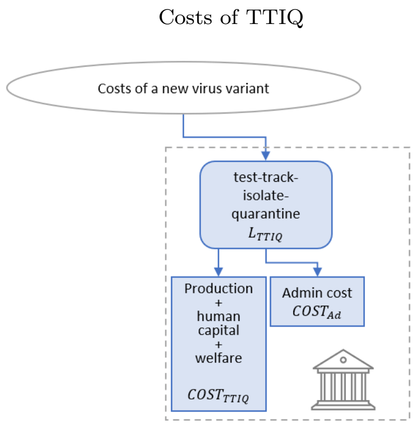

To calculate the costs of the TTIQ system, we assume that when the system works at the maximum, ie. $TTIQ(t) = 1$, the tracking system detects 70% of symptomatic cases and 30% of asymptomatic cases. For each detected case, we assume on average 3 contacts who are quarantined for 7 days. All symptomatic cases experience a productivity loss, a welfare loss and a health loss due to illness, and the detected 70% of them also experience an additional cost due to quarantine while asymptomatic for a total of 7 days, see the right column in Table S15. All discovered asymptomatic cases are isolated for 7 days in the model. The costs of the TTIQ-system from quarantine and isolation are assumed to be proportional to $TTIQ(t)$, the capacity use of the system.

The total cost of TTIQ is given by equation (32), which includes both the economic cost of staying at home due to isolation/quarantine, and the administrative cost of operating the system:

$$L_{TTIQ} = \sum_{t=1}^{T}\sum_{i=1}^{9} TTIQ(t) \times inc_i(t) \times COST^i_{TTIQ} \times \left(f^i_a \rho^i_a D^i_{TTIQ} + f^i_s (D^i_{TTIQ} - D^i_{sick}) + N_c p^i_{det} D^i_{TTIQ}\right)$$
$$+ COST_{Ad}(N_{TTIQ}, \tilde{I}) \sum_{t=1}^{T} \frac{TTIQ(t)}{T} \qquad (32)$$

The variable $inc_i(t)$ is the incidence of new infections per day, where $\tilde{I} = \sum_t \sum_i inc_i(t)$ is the cumulative incidence (total infections). The variable $D^i_{TTIQ}$ is the length of isolation and quarantine, see Table S15, we subtract $D_{sick}$ from $D_{TTIQ}$ since the cost of staying home when ill is accounted for in $L_{sick}$. $\rho^i_a = 0.3$ is the probability of detecting an asymptomatic infection, $COST^i_{TTIQ}$ is the cost per day in quarantine or isolation, $N_c = 3$ is the number of contacts identified during the contact tracing and $p^i_{det} = f^i_s \rho^i_s + f^i_a \rho^i_a$ is the probability of an infection being detected for contact tracing with $\rho^i_s = 0.7$.

There is an administrative cost $COST_{Ad}$ to sustain the organizing of the public TTIQ-system. We assume the administrative part of the contact tracing regime has an initial setup cost of 500 mnNOK and a fixed cost of 2.74 mnNOK per day of operation, $N_{TTIQ}$, and a variable cost of 2 780 NOK per infected person per day, $\tilde{I}$, in terms of tests and personnel performing and analyzing the tests, so that the total administrative costs are:[8]

$$COST_{Ad}(N_{TTIQ}, \tilde{I}) = \text{mnNOK}\,500 + \text{mnNOK}\,2.74 N_{TTIQ} + \text{NOK}\,2780\tilde{I}. \qquad (33)$$

[8]Norwegian municipalities received 3 bnNOK in 2021 as extraordinary payments, earmarked as discretionary funds for infection control and the TTIQ-strategy [41]. We assume half (3/2=1.5 bnNOK) covers fixed costs (where 1/3 = 500 mnNOK is initial setup and 2/3 is daily fixed costs 1000 mnNOK/365 = 2.74 mnNOK per day), while the other half corresponds to variable costs, 2780 NOK per infected.

#### B.5 The costs of sick leave

Individuals with symptomatic illness will be absent - on sick leave - from work, school or other activities irrespective of whether TTIQ is activated. Sick leave creates a loss of production, a loss of human capital and a loss of welfare, depending on the individual's age, which come in addition to the age-dependent health loss in terms of QALYs. These costs apply also if symptomatic illness evolves into more severe outcomes such as hospitalization, ICU and/or post-acute sequale.

The economic and welfare cost of infected individuals staying at home or in hospital is given by:

$$L_{SICK} = \sum_i COST^i_{sick} f^i_s \tilde{I}_i D^i_{sick}, \tag{34}$$

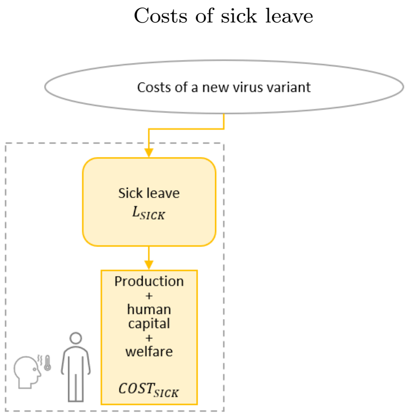

where $f^i_s$ represents the fraction of symptomatic infections in age group $i$, given in Table S3 and the variable $\tilde{I}_i$ denotes the total number of infected individuals per age group at the end of the simulation. $D^i_{sick}$ is the age-dependent duration of the outcome of the infection (mild illness, hospitalization, ICU, post acute sequale). $COST^i_{sick}$ is the individually monetary cost per day of absence, including both economic and welfare costs. If the individual is hospitalized the duration includes the hospital stay. The details of $COST^i_{sick}$ are described in Table S15, and further explained in section B.6.1–B.6.3.

#### B.6 Economic and welfare costs

There is a welfare cost and an economic cost of staying at home when individuals are either sick (on sick leave) or required to stay in quarantine in cases when TTIQ is active, see Table S15. These costs come in addition to the age-dependent health loss when sick.

The 'economic cost' consists of two components, depending on age: A human capital loss denoted $HU^i$ and lost production denoted $Y^i$. In addition to the 'economic cost', individuals experience reduced life satisfaction resulting in a welfare loss, $W^i$.

| | When TTIQ is OFF | | When TTIQ is ON | |
|---|---|---|---|---|
| Age | Economic loss per day when staying at home symptomatic (sick leave) $COST^i_{sick}$ | Length of illness (including hospital) | Economic loss per day when staying at home asymptomatic (quarantine or isolation) $COST^i_{TTIQ}$ | Length of quarantine or isolation (days) $D_{TTIQ}$ |
| 0-9 | $W \times 2 + (1/2 + 1/2 1/2)Y$ | $D^i_{sick}$ | $W \times 2 + (1/2 + 1/2 1/2)Y$ | 7 |
| 10-19 | $W + HU$ | $D^i_{sick}$ | $W + HU$ | 7 |
| 20-29 | $W + Y$ | $D^i_{sick}$ | $W + 1/2 Y$ | 7 |
| 30-39 | $W + Y$ | $D^i_{sick}$ | $W + 1/2 Y$ | 7 |
| 40-49 | $W + Y$ | $D^i_{sick}$ | $W + 1/2 Y$ | 7 |
| 50-59 | $W + Y$ | $D^i_{sick}$ | $W + 1/2 Y$ | 7 |
| 60-69 | $W + Y$ | $D^i_{sick}$ | $W + 1/2 Y$ | 7 |
| 70-79 | $W$ | $D^i_{sick}$ | $W$ | 7 |
| 80+ | $W$ | $D^i_{sick}$ | $W$ | 7 |

Note: Per person per day the welfare loss $W$ is 285 NOK, the human capital loss $HU$ is 570 NOK and the production loss $Y$ is 1500 NOK.

Table S15: The economic and welfare costs of staying at home when either sick or in quarantine, per age group

###### B.6.1 Welfare loss $W$

A loss of welfare arise when there are limitations on personal freedom and choice. As explained above, we assign a monetary value of the welfare loss from staying at home based on [40], leading to an average loss of 285 NOK per person per day.

###### B.6.2 Human capital loss $HU$

During a pandemic, with frequent quarantines and infections, the youth will have fewer days at school. We assume quarantines and infection give a loss of human capital for the age group 10-19 years. For children aged 0-9 we assume that there is not any significant human capital loss.

**Monetary value:** In order to set a value of the human capital loss in a Norwegian setting we use the same approach as [42]. They use data and parameters from the USA to estimate future income loss for children in the USA due to half a year of school closure because of Covid-19, finding a lifetime income loss of 0.95% for children between 4-14 years old measured in lost income over the course of their lives [42].

According to [43], the average lifetime income in Norway is 11 mnNOK, of which a loss of 0.95% corresponds to 104,500 NOK per person for half a year of school closure. 104 500 NOK / 182.5 days = NOK 572.6 ≈ NOK 570 per day per person.

###### B.6.3 Production loss $Y$

**Remote work:** We denote $Y$ as the average production loss per day for a person who is not able to work due to sickness or quarantine. We assume that asymptomatic individuals in the age groups 20-69 years can still work while in quarantine, if they have the opportunity to work remotely. We assume half of them do not have this option, implying an average loss per person equal to $^1/_2 Y$.[9]

**Child care:** For the youngest age group, we assume there will be both a production loss due to childcare needs and a welfare loss for both the child and caregiver when staying home during a pandemic. When children aged 0-9 years must stay home due to symptomatic infection, isolation, or asymptomatic but in quarantine, a caregiver is also required to stay at home. We assume all caregivers are part of the workforce, and one caregiver per child.

Furthermore, we assume that half of these caregivers are unable to work remotely. For the half who can work from home, we assume that balancing childcare with remote work enables only half a day of productive work. Altogether, when young children are required to stay home this results in the following production loss per child:

[9]There is no formal registration of remote work in Norway. The assumption that half the working force can work from home is based on several surveys: [44] found that 39 percent of jobs in Norway could be performed from home. The Work Reserach Institute (AFI) mapped the prevalence of remote work during the pandemic in Norway and found that 50 percent of employees had the opportunity for remote work in March 2020, while in February 2021, 52 percent said the same [45]. According to a consumer survey by Opinion [46] nearly 60 percent of Norwegian employees had remote work arrangements March 2020. Surveys conducted by [47] showed that close to 56 percent was working remotely in April 2020 in and around Oslo.

$$\text{Production loss per day per caregiver} = (\underbrace{1/2}_{\substack{\text{Unable}\\ \text{to work}\\ \text{remote}}} + \underbrace{1/2 \, 1/2}_{\substack{\text{work}\\ \text{remote}\\ \text{half}\\ \text{speed}}})Y \tag{35}$$

**Monetary value:** From the national accounts of Norway, Total hours worked for employees and self-employed (million workhours) in 2021 was 4 034 million hours [48]. For simplicity we assume that all production is done by the age group 20-69 years, which is a population of 3 467 thousand individuals [49]. The number of hours worked per day per person in the age group 20-69 years old $= 4\,034/(365 \times 3\,467) =$ 3.2hours:

$$\frac{\text{All hours worked in the economy}}{\text{All individuals in working age}} = \text{hours worked per person in working age} \tag{36}$$

According to the national accounts, the average wage cost per hour worked was 487 NOK for Mainland Norway in 2021 [48]. The average production loss per day due to illness (symptomatic) for individuals in the age group 20-69 years old is assumed to be 487 NOK * 3.2 = 1 552 NOK. Rounded to 1 500 NOK (2021) per person per day.

##### B.6.4 Teaching and schools

Several studies document significant human capital losses from digital home-schooling, particularly for students with fewer resources at home, see e.g. [50], [51], and [42]. In Norway, however, digital teaching and school closures were relatively limited during the pandemic.[10] The Norwegian expert group [17] examined National Test results from 2020 and 2021 and found no evidence of significant learning loss compared to pre-pandemic years; average test scores remained stable over time.[11] Similarly, [52] find no significant learning loss using Danish national test data.

Overall, this suggests limited human capital loss from the pandemic in Norway. This aligns with the fact that schools largely remained open and indicates that the widely used *traffic light model*[12] in schools and preschools has not had substantial effects on human capital. Accordingly, we do not include costs from learning loss due to school closures, except for losses due to absence associated with quarantine through TTIQ and illness. Nevertheless, the long-term effects of school closures and digital learning remain uncertain, underscoring the importance of further research for future pandemics.

#### B.7 Costs by intervention level

For illustration, Figure S2 shows how the different types of costs depend on the level of interventions $G(t)$. In the simulations for Figure S2, $G$ is not chosen optimally, but is kept constant throughout the epidemic at the level indicated on the horizontal axis. The initial reproduction number is $R = 1.8$, and the three panels show results for three different severity levels: 1, 7 and 15.

The line 'Contact Reduction' shows the economic and welfare costs from contact reduction caused by government interventions and VSD. This line is downward-sloping for $G < 0.25$, indicating that a

[10] In Norway, schools and preschools were closed during the first wave of the pandemic in spring 2020 but began to reopen in late April 2020.

[11] Stable test results may reflect limited learning loss, successful compensation by schools, the nature of the tests, or limitations in year-to-year comparability.

[12] Introduced in 2020 to allow flexible adjustment of school measures to local infection levels [53].

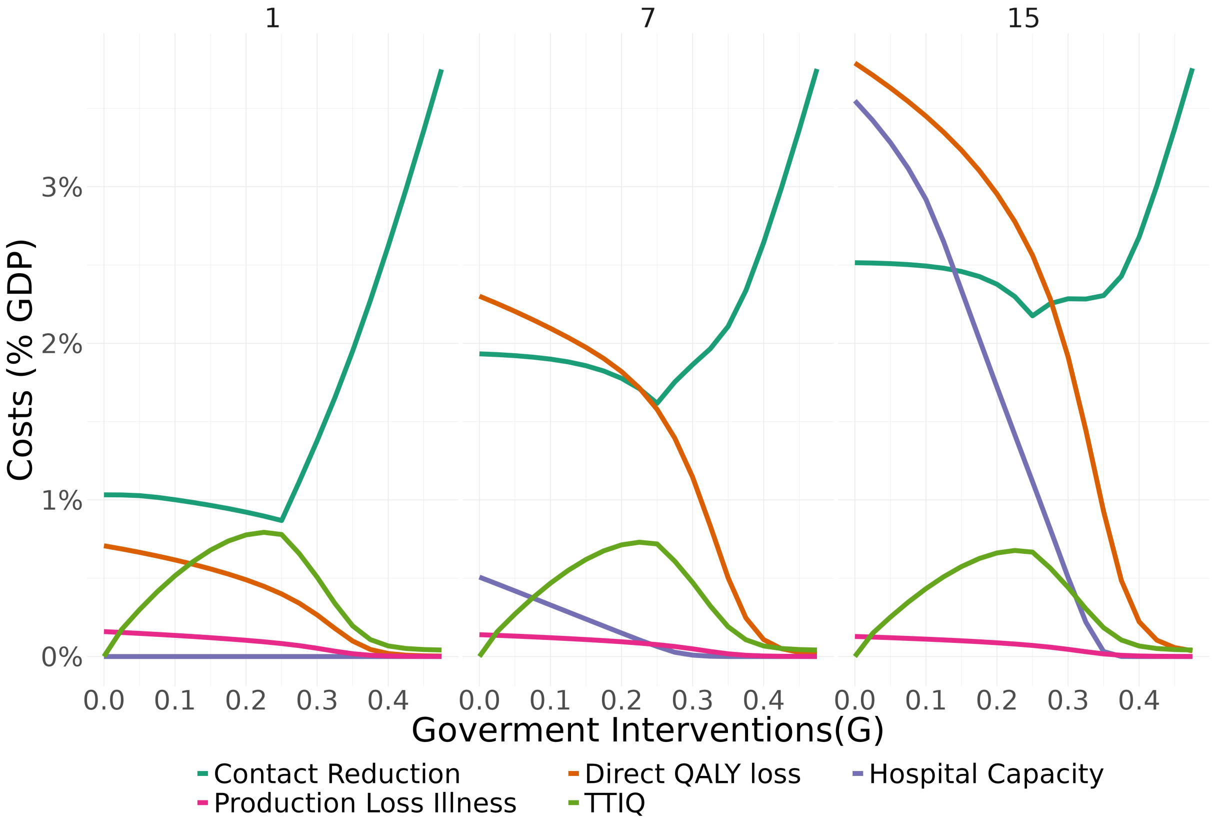

Figure S2: **The baseline case: Cost groups.** The figure shows how the various types of costs depend on the intervention level $G(t) \in [0, 1]$, assuming that the intervention level is kept constant throughout the pandemic, for $R = 1.8$ and severity levels $ES = \{1, 7, 15\}$. For the scenarios shown here, interventions $\approx 0.5$ are sufficient to reach $R = 1$.

higher $G$ in this interval leads to lower costs from contact reduction. This pattern arises because in the interval $G \in [0, 0.25]$, a higher $G$ represents stronger TTIQ, which reduces infections and thereby reduces VSD in the model. For $G > 0.25$, contact-reducing NPIs are imposed, and the costs of contact reduction increase sharply with $G$. 'Direct QALY Loss' shows the monetized health loss from the disease, $L_Q$ in equation [17], which is decreasing in $G$ because interventions reduce the number of infections. 'Hospital Capacity', $L_{HC}$, is the excess health costs associated with higher mortality rates in overloaded hospitals. From the high-severity panel (right), we see that these costs may be large for high severity combined with a low level of interventions $G(t)$. 'TTIQ' shows the costs from Test-Trace-Isolate-Quarantine, $L_{TTIQ}$ in equation [17], which are increasing in $G$ in the interval $[0, 0.25]$, when TTIQ is used to the full extent. Higher $G$, above 0.25, indicates other contact-reducing NPIs, which reduce the number of infections and thus also reduce the costs from TTIQ. 'Production Loss Illness', $L_{SICK}$, is the economic and welfare cost for infected individuals, which is only a small share of the total costs.

### C Voluntary social distancing

#### C.1 Method

We assume that individuals adjust their contact rates by weighing the health risk from the disease against the welfare costs of staying at home, which we refer to as voluntary social distancing (VSD). Specifically, individuals compare the welfare cost of staying at home over a 60-day period with the expected health cost associated with maintaining normal social activity. The health loss is based on the contemporaneous probability of infection multiplied by the age-specific QALY loss from infection. The parameters are chosen so that VSD reduces peak incidence by approximately 50% and peak hospital admissions by approximately 75% in a SARS-CoV-2-like epidemic ($ES = 15$) without government intervention, cf. figures S3.B and S3.C.

Let $p^i_{inf}$ denote the approximate risk of infection per day, given by the contemporaneous infection rate in the age-group, given by $p^i_{inf} = \frac{E^i_s + E^i_a}{D_L}/S^i$ where $E^i_s$ and $E^i_a$ are the number of exposed individuals (who will become symptomatic and asymptomatic, respectively) and $D_L$ is the latent period, while $S^i$ is the number of susceptible individuals. Assuming a constant per-day probability of being infected, the probability of being infected within the 60-day period ($\tau = 60$) is $\big(1 - (1 - p^i_{inf})^\tau\big)$.

The average welfare cost of staying at home, $W^i$, is set to 285 NOK per day per person, based on the results from a choice-experiment in Sweden during the early stage of the pandemic [40]. As the experiment shows considerable heterogeneity across individuals, we assume that $W^i$ is uniformly distributed in the population between zero and $W_{\max}$. Hence, the costs are uniformly distributed from 0 to 570 NOK per day.

We use a logistic transformation to ensure a gradual response from the relative utility of social distancing (i.e. expected health cost relative to the welfare costs from social distancing) to the actual choice taken by the individual. The idea is to capture that individuals may abstain from actions involving high risk, yet they may take some risk in other situations.

For an individual with welfare costs of staying at home $W^i = 285$ NOK/day and an assumed duration of $\tau = 60$ days, we assume that the reduction in contact behaviour for an individual in age-group $i$ is given by (time is omitted):

$$V^i(W) = \Gamma\Big(a \times \big(COST_Q \times E^i_{QALY} \times \big(1 - (1 - p^i_{inf})^\tau\big) - W\tau\big)\Big), \tag{37}$$

where $\Gamma(x) = \exp(x)/(1 + \exp(x))$. The expected health cost from infection, $COST_Q \times E^i_{QALY}$, denotes the monetary value of the age- and scenario-specific expected QALY loss (on a 0-1 scale) if infected. The parameter $a = 4/(9W\tau) = 0.0005$ bnNOK$^{-1}$ is to smooth the transition between no and full voluntary social distancing and is set such that if the expected health cost is 30% higher than the cost from avoiding social contact, the individual will reduce his/her social contact corresponding to a reduction in $V^i$ by 75%.

The amount of voluntary social distancing (VSD) in each age group is then given by:

$$V^i_T = \frac{1}{W_{\max}\tau}\int_0^{W_{\max}} V^i(W)dW = \frac{\log(\exp(aC) + \exp(aW\tau))}{a} + \frac{\log(\exp(aC) + 1)}{W_{\max}\tau} \tag{38}$$

Where $C = COST_Q \times E^i_{QALY} \times \left(1 - (1 - p^i_{inf})^{\tau}\right)$. And the total amount of VSD is:

$$V_T = \sum_{i=1}^{9} V_T^i. \tag{39}$$

#### C.2 No VSD and Altruism

To what extent individuals adjust behavior during a pandemic to reduce their health risk has important implications for the optimal strategy and associated costs. VSD depends on information available to the public, including guidance from health and political authorities. [54] argue that the sharp decline in consumption early in the pandemic in Portugal reflected pessimistic beliefs about case-fatality rates.

| **Specification** | $E^i_{QALY}$ |
|---|---|
| Scenario: Baseline | $E^i_{individual}$ |
| Scenario: No VSD | — |
| Scenario: Altruism | $E^i_{individual}+$ $E^i_{contacts}$ |

Table S16: VSD cases

In this subsection we compare our baseline VSD-assumption with a scenario where there is no VSD and a scenario where individuals take into account both their own health risk as well as the health risk they may cause by infecting their contacts. The latter we refer to as Altruism. Thus, in total we consider three different specifications for the expected QALY loss in the individual trade-off, $E^i_{QALY}$ in equation (38), see table Table S16. See Figures 6 and 7 in Main paper for the results.

In the specification labeled Altruism, the expected QALY loss, $E^i_{QALY}$, is replaced by a combination of the individual's own expected health loss, $E^i_{individual}$, and the expected health loss of the average person infected by someone from age group $i$, denoted $E^i_{contacts}$. The latter is defined as the expected contact-weighted average health loss of individuals in age groups with whom group $i$ interacts:

$$E^i_{contacts} = \frac{\sum_j E^j_{individual} M_{ij}}{\sum_j M_{ij}},$$

To illustrate the effect of the three specifications, Figure S3.A shows implied VSD across age groups during an unconstrained epidemic, for three severity levels, $ES = \{1, 9, 15\}$. With VSD, the oldest groups select maximum distancing at the epidemic peak. However, in baseline, when individuals only take into account their own health risk, younger groups undertake little social distancing. If individuals fully account for both their own and their contacts' health costs (altruism), distancing also increases among younger age groups. The effect is nevertheless limited, partly because interactions are concentrated among similarly low-risk age groups.

Figure S3.B and S3.C show the effect of varying the assumptions about voluntary social distancing on incidence rates and hospitalizations during a pandemic without interventions. VSD gives a clear reduction in incidence rates, in particular for high severity levels. However, the reduction is substantially larger for hospital admissions, reflecting stronger distancing among older age groups, who face higher risks of severe outcomes and hospitalization. We also observe that altruism further amplifies the reduction in infections and hospitalizations, particularly at high severity levels.

Table S17 reports the share of simulations in Figures 6 and 7 in the main paper where total costs increase or decrease relative to baseline.

Figure S4 shows the change in infection rates among individuals above 60 years under altruism relative to baseline. The blue color indicates that altruism leads to fewer infections among older age groups in

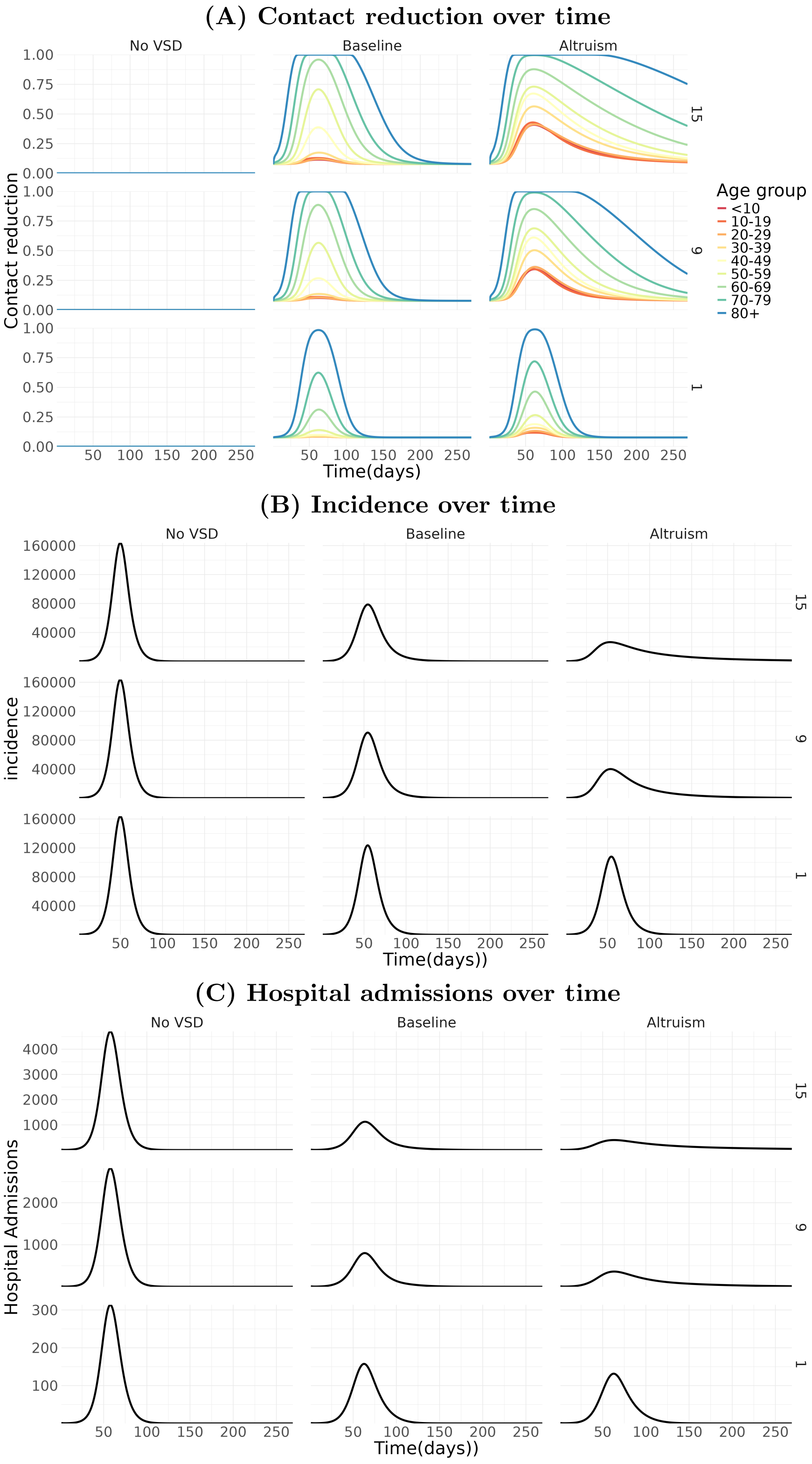

Figure S3: Two alternative specifications of voluntary social distancing: 'No VSD' and 'Altruism' compared to the baseline case. For $R = 2$ and $ES = 1, 7, 15$ in rows. Trajectory with no governmental interventions $G(t) = 0$.

| VSD type | $s_Q \times COST_Q$ | With increased cost relative to baseline VSD | With decreased cost relative to baseline VSD |
|---|---|---|---|
| No VSD | $0.5 \times COST_Q$ | 609 (42%) | 831 (58%) |
| No VSD | $1.0 \times COST_Q$ | 683 (47%) | 757 (53%) |
| No VSD | $2.0 \times COST_Q$ | 860 (60%) | 580 (40%) |
| Altruism | $0.5 \times COST_Q$ | 1424 (99%) | 16 (1%) |
| Altruism | $1.0 \times COST_Q$ | 1195 (83%) | 245 (17%) |
| Altruism | $2.0 \times COST_Q$ | 706 (49%) | 734 (51%) |

Table S17: Number (share) of simulations where total costs $L_T$ increased or decreased for No VSD and Altruism

scenarios where the health costs from the pandemic are low, due to low severity and/or low QALY-value. In these scenarios, the optimal policy is *No/negligible intervention* implying substantial epidemic overshoot, and altruism reduces infections by mitigating the overflow. In scenarios with greater health costs, associated with higher severity and/or high QALY-value, the red color shows that altruism leads to higher infection rates among individuals above 60. In many scenarios, the main reason is that altruism leads to a change of strategy to less interventions, from *Delayed containment* to *No/negligible intervention*, implying higher overall infection rates. However, in some scenarios, eg. for baseline QALY-value, so = 1 and 1000 days, $ES > 15$ and $R > 1.6$, the optimal strategy is *Delayed containment* under both altruism and baseline. In these scenarios, altruism leads to higher infection rates for a given strategy, by shifting infections from younger groups to older groups.

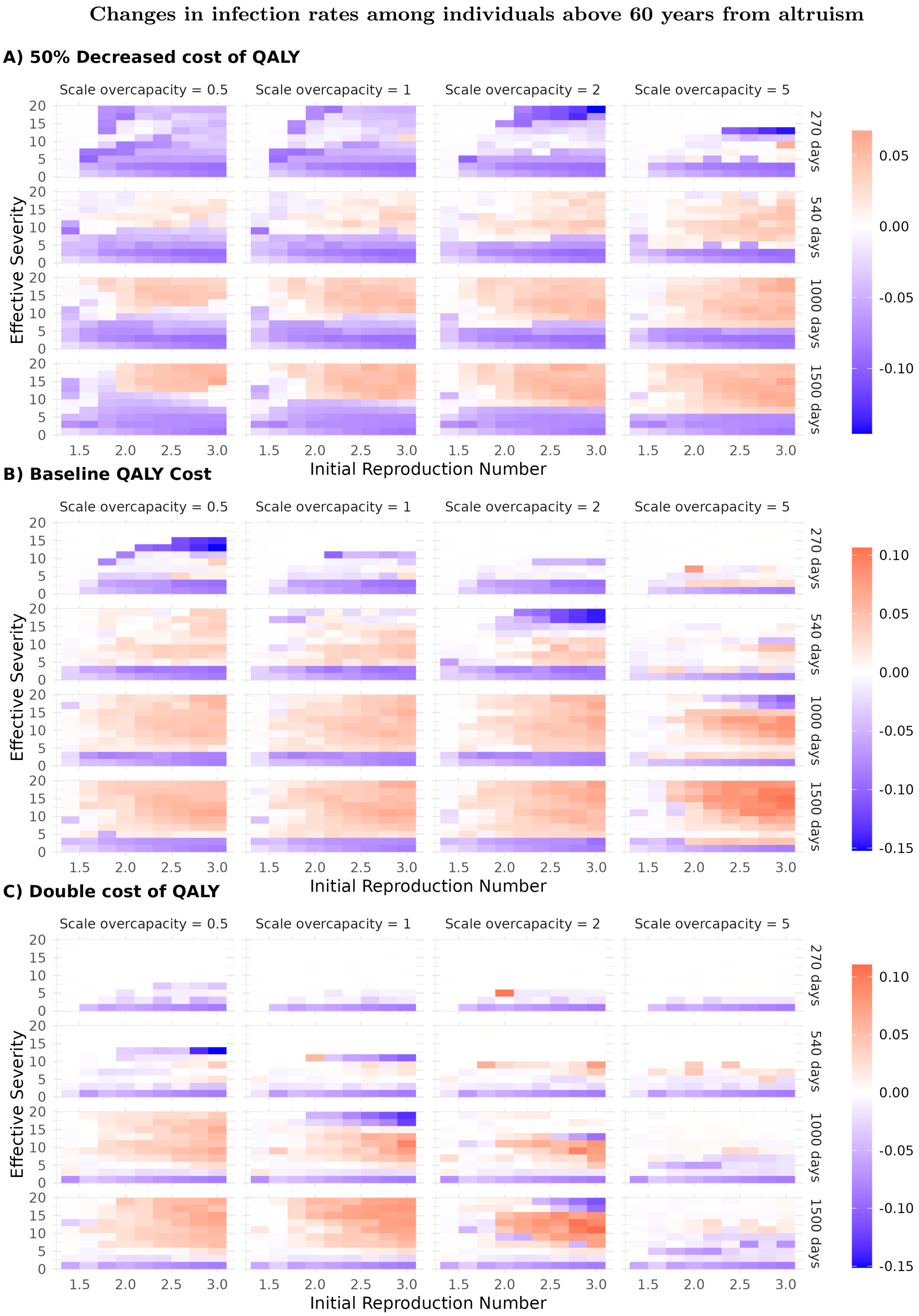

Figure S4: **Change in infection rates above 60 years from altruism.** The figure shows the change in infection rate for individuals above 60 years under the optimal strategy with altruism compared to the baseline VSD assumptions. Here a change of 0.1 (red color) indicates that an additional 10% of individuals above 60 years were infected in the scenarios with altruism.

### D Supporting figures and results

#### D.1 Categorizing optimal solutions

##### D.1.1 The categorization rule

After identifying the $G(t)$-trajectories that minimize social costs, we construct a qualitative categorization rule to label the optimal strategies, which is applied consistently across all cases. The classification presented in the main paper, and used to classify strategies in Figure 4, is based on characterizing each $G(t)$-trajectory using up to six quantitative variables. The first three variables characterize the policy dimension of the optimal strategy (intervention intensity and timing), whereas the remaining three capture epidemiological outcomes in terms of immunity, incidence and infections:

| | | | |
|---|---|---|---|
| $ARI$ | Average Relative Intervention | Mean intervention level $\overline{G(t)} = \frac{1}{T}\sum_{t=0}^{T} G(t)$ relative to a $G(t)$-trajectory set to ensure $R = 1$ | $ARI = \frac{\overline{G(t)}\|_{\text{opt}}}{\overline{G(t)}\|_{\text{force } R=1}}$ |
| $MINT$ | Max Intervention level | Maximum intervention level over time | $\max_t G(t)$ |
| $t$ | Timing | Days until first intervention is implemented | $G(t) = 0$ until day $t$ |
| $RMI$ | Relative Max Incidence | Maximal incidence in the optimal strategy relative to no intervention | $\frac{\max_t inc(t)\|_{\text{opt}}}{\max_t inc(t)\|_{\text{Noi.}}}$ |
| $FHI$ | Fraction of Herd Immunity | Fraction infected relative to the herd immunity threshold $\tilde{I}_{\text{Herd}}$ | $\tilde{I}/\tilde{I}_{\text{Herd}}$ |
| $IR$ | Infection ratio | Fraction infected relative to no interventions $\tilde{I}_{\text{Noi.}}$ | $\tilde{I}/\tilde{I}_{\text{Noi.}}$ |

See Table S18 for a summary of the rules we apply to label strategies. The rules are chosen with the aim of obtaining clear categories where strategies with similar features are grouped together. As will become apparent from Figures S5, S9 and S10 below, the large majority of optimal strategies fall in distinct clusters according to a combination of intervention strategy and pandemic outcome. However, for some parameter combinations there are also a limited set of borderline cases where the distinction between strategy types is blurred, in particular between suppression and mitigation. In these few cases, the exact thresholds used to classify categories may affect the classification.

| **Strategy name:** | **Rule:** | | | | | |
|---|---|---|---|---|---|---|
| | $ARI$ | $MINT$ | Intervention start | $RMI$ | $FHI$ | $IR$ |
| **No intervention** | $= 0$ | | | | | |
| **Suppression** | $> 0.65$ | | | $< 0.3$ | | |
| **Delayed containment** | | $\geq 0.3$ | on/after day 15 | | | |
| **TTIQ-only** | | $\approx 0.25$* | on/after day 15 | | | |
| **Negligible intervention** | | | | | | $\geq 0.95$ |
| **Flatten-the-curve** | | | | $< 0.5$ | $> 0.6$ | |

*Notes:* *Due to approximations in the numerical optimization procedure this is set in the interval $\leq 0.255$ and $\geq 0.245$

Table S18: The rule for categorizing optimal $G(t)$-trajectories

We use the label *No intervention* for trajectories without government intervention ($ARI = 0$). *Suppression* refers to trajectories with high intervention intensity ($ARI > 0.65$) that keep infections low ($RMI < 0.3$). *Delayed containment* refers to trajectories where interventions are introduced no earlier than day 15 with sufficiently high intensity ($MINT \geq 0.3$). *TTIQ-only* refers to trajectories where interventions are introduced no earlier than day 15, and interventions, if used, are TTIQ at full level ($G = 0.25$). *Negligible intervention* refers to trajectories where some intervention may be present, but where reduction in the total number of infections is less than 5 % compared to *No intervention* ($IR \geq 0.95$), implying that the epidemiological effect of the interventions is negligible. *Flatten-the-curve* refers to trajectories where peak incidence is substantially reduced relative to *No intervention* ($RMI < 0.5$), while a large share of the population is still infected ($FHI > 0.6$). The classification hierarchy follows table S18, implying that if a trajectory satisfies the criteria of two categories, it is classified as the category listed first in S18. Strategies that do not fall into any of the other categories are grouped in *Other*. Figure S5 shows the distribution of combinations of $ARI$, $RMI$ and $FHI$ in the baseline calibration presented in Figure 2 in the main paper. The observed clusters are used to inform the classification criteria presented in table S18.

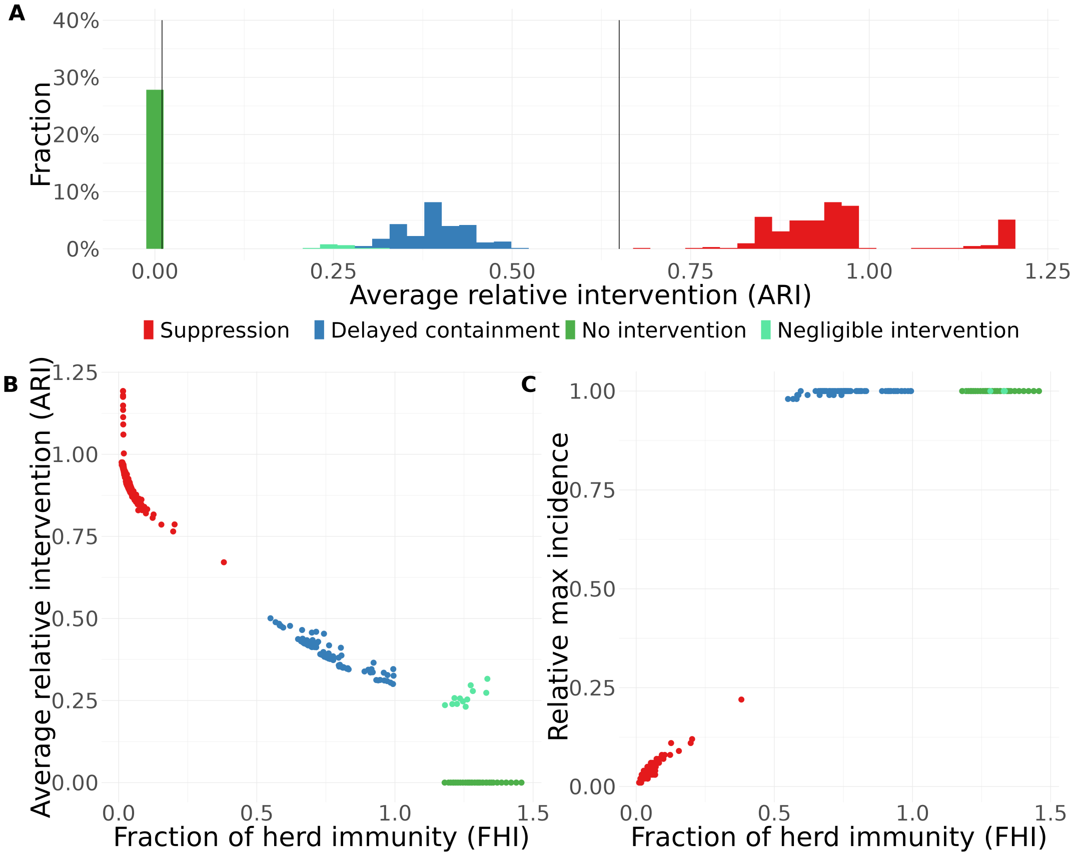

Figure S5: **Baseline case.** Each dot corresponds to the optimal strategy for one of the 380 $(R, ES)$-combinations in the baseline calibration in Figure 2 in the main paper, where $R \in [1.2, 3]$ and $ES \in [1, 20]$. ($19 \times 20 = 380$ combinations)

Figure S6 shows the incidence curves for the same optimal strategies in the baseline calibration in Figure 2 in the main paper, further emphasizing the qualitative difference between different groups of optimal strategies.

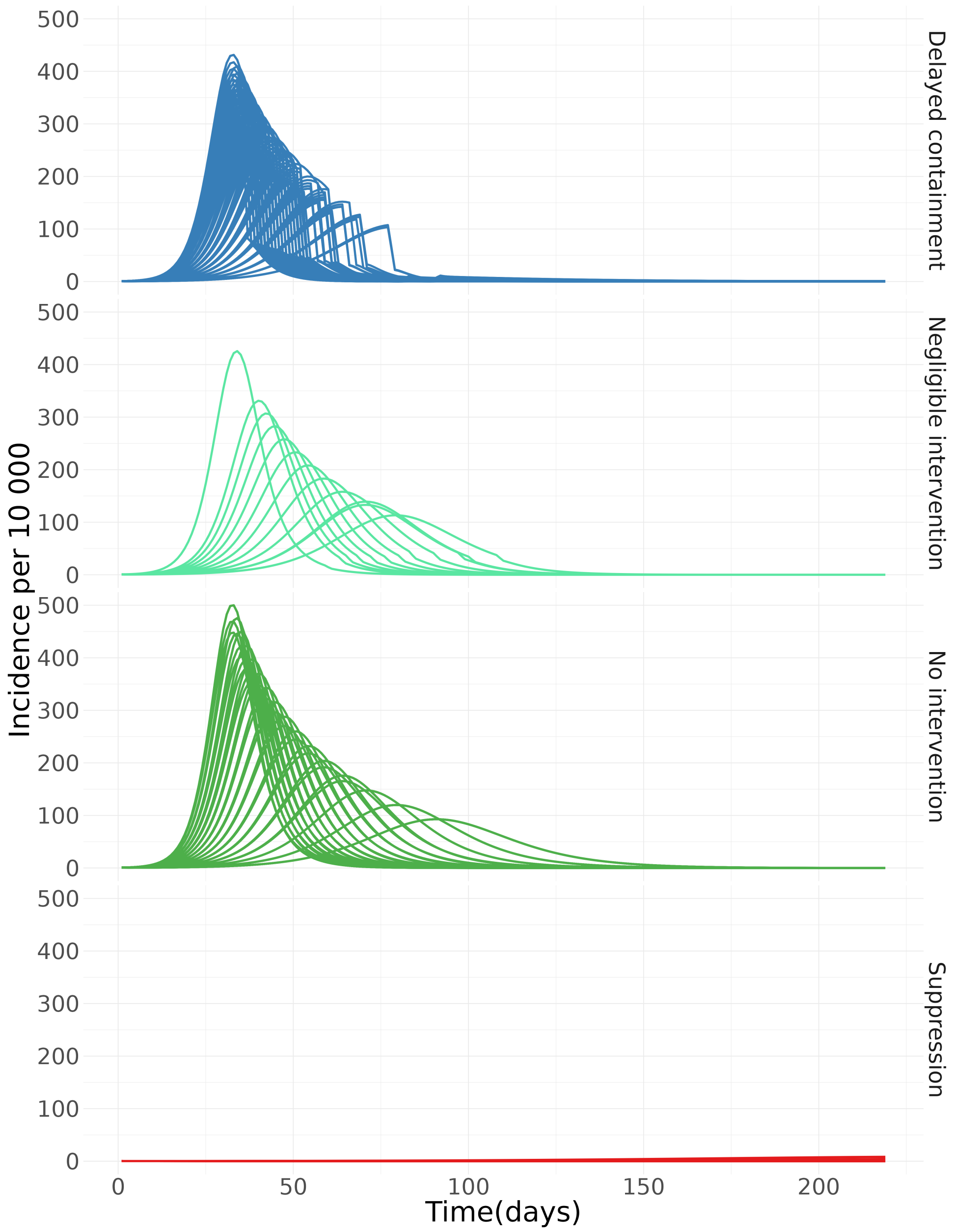

Figure S6: **Incidence of infections** sorted into strategy types in the baseline calibration. Each line represents one of the 380 $(R, ES)$-combination, where $R \in [1.2, 3]$ and $ES \in [1, 20]$. $(19 \times 20 = 380$ combinations)

##### D.1.2 Key properties

In this section, we present alternative visualizations of the strategies presented in Figure 4 in the main paper (Figures S7–S10), to explore key properties of the strategy types. All figures are based on the parameter combinations in Table S19, as in Figure 4.

| Parameter | Values |
|---|---|
| Initial reproduction number | $R \in \{1.4, 1.6, 1.8, 2.0, 2.2, 2.4, 2.6, 2.8, 3.0\}$ |
| Effective severity | $ES \in \{1, 3, 5, 7, 9, 11, 13, 15, 17, 19\}$ |
| Length until effective vaccine | $T \in \{270, 540, 1000, 1500\}$ days |
| Scaling of baseline QALY costs $s_Q \times COST_Q$ | $s_Q \in \{0.5, 1, 2\}$ |
| Scaling of overcapacity costs $s_o \times L_{HC}$ | $s_o \in \{0.5, 1, 2, 5\}$ |

Table S19: Parameter values for broad parameter scan

Figure S7 shows the numbers of each of the optimal strategy types in Figure 4 in the main paper, and the key numbers are summarized in table S20.

| $s_Q \times COST_Q$ | *No intervention* | *Negligible intervention* | *Suppression* | *Delayed containment* | *TTIQ-only* | *Flatten-the-curve* | *Other* |
|---|---|---|---|---|---|---|---|
| $0.5 \times COST_Q$ | 266 (18%) | 250 (17%) | 123 (9%) | 734 (51%) | 53 (4%) | 2 (0%) | 12 (1%) |
| $1.0 \times COST_Q$ | 178 (12%) | 53 (4%) | 305 (21%) | 876 (61%) | 22 (2%) | 7 (1%) | 9 (1%) |
| $2.0 \times COST_Q$ | 120 (8%) | 16 (1%) | 636 (44%) | 623 (43%) | 5 (0%) | 25 (2%) | 15 (1%) |

Table S20: Number, and share in percentage, of optimal strategies in figure 4 in the main paper

Figures S8-S11 show key properties of the optimal strategies.

Figure S8 shows how the Fraction of herd immunity (FHI) and the Relative max incidence (RMI) depend on the different parameter values, using the same parameter combinations as Figure 4 in the main paper, implying that the upper part Figure S8.A shows results for QALY cost equal to 50% of the baseline value, while the 4x4 panels exhibit different scales of excess costs of overcapacity (horisontally) and time until vaccination (vertically).

Consider first the top left panel of Figure S8.A, where the overcapacity costs in the health sector is half of the baseline case (scale overcapacity = 0.5), while effective vaccination is after 270 days as in the baseline case. The red dots in the bottom left corner represent $(R, ES)$-combinations where *suppression* is the optimal strategy. We observe that the relative max incidence RMI is below 0.25 while the fraction infected relative to herd immunity FHI is below 0.5, indicating that strict interventions are used to keep the epidemic down. The blue dots at the top represent $(R, ES)$-combinations where *Delayed containment* is the optimal strategy. RMI is close to one, as no interventions are used until close to the peak of the epidemic. However, FHI varies from 0.5 to 1, reflecting that strict interventions are used to stop the epidemic, followed by TTIQ to control the epidemic even for an infection rate below herd immunity. The green dots at the top right corner represent $(R, ES)$-combinations where the optimal strategy is *No intervention*. RMI is equal to one and FHI is well above one, reflecting epidemic overshoot. The light green dots indicate a *Negligible intervention* which reduces epidemic overshoot, but does not prevent it, as indicated by FHI being above one.

Looking more broadly at the results in Figure S8, the same distinct categories prevail in the large majority of the panels, except that suppression is not optimal with long time until vaccination and low excess costs of overcapacity. However, for higher monetary costs of a QALY, in particular for double costs, the picture is less clear-cut in the lower right corner with max overcapacity costs and/or max time to vaccination. As we have seen, long time until vaccination makes suppression very costly, and the delayed containment strategy is less attractive for high overcapacity costs. Thus, in scenarios with long time until vaccination and high overcapacity costs (lower right corner), *Flatten-the-curve* is optimal for some combinations (orange dots), where intermediate interventions are used to prevent high infection rates, yet the interventions are not sufficiently strict to prevent that a large part of the population is infected. The purple dots indicate $(R, ES)$-combinations where the optimal strategy does not fall into any of the three main categories. For short time until vaccination, most or all of the *Other* strategies seem to be variants of delayed containment strategies that do not meet our exact pre-defined criteria. For delayed vaccination, many of the *Other* categories seem to be strategies that lie between suppression and flatten-the-curve. Most resemble suppression, but with lower intervention intensity.

Figure S9 shows the average relative intervention ($ARI$) and the fraction of herd immunity ($FHI$) for the optimal strategies. For the *Suppression* strategies, there is some variation in average relative intervention ($ARI$) and fraction of herd immunity ($FHI$), but they all keep $FHI$ below 0.5. The *Delayed containment* strategies have lower $ARI$ and higher $FHI$, but $FHI$ is still kept below 1. *No/negligible intervention* involve lower intervention and pandemic overshoot, resulting in $FHI$ above 1. For high overcapacity costs and/or double QALY-cost, there are also some intermediate *Flatten-the-curve* or *Other* strategies.

Figure S10 shows the infection ratio (IR), defined as the fraction infected under the optimal strategy relative to the fraction infected under no interventions, and the max intervention level for the optimal strategies. While there is considerable variation across suppression strategies, they are nevertheless qualitatively different from the *Delayed containment* strategies and the *No/negligible intervention* strategies. In Figure S11 we show the density of FHI values for the different strategies. The peaks of the distributions reflect that the strategies cluster in distinct categories.

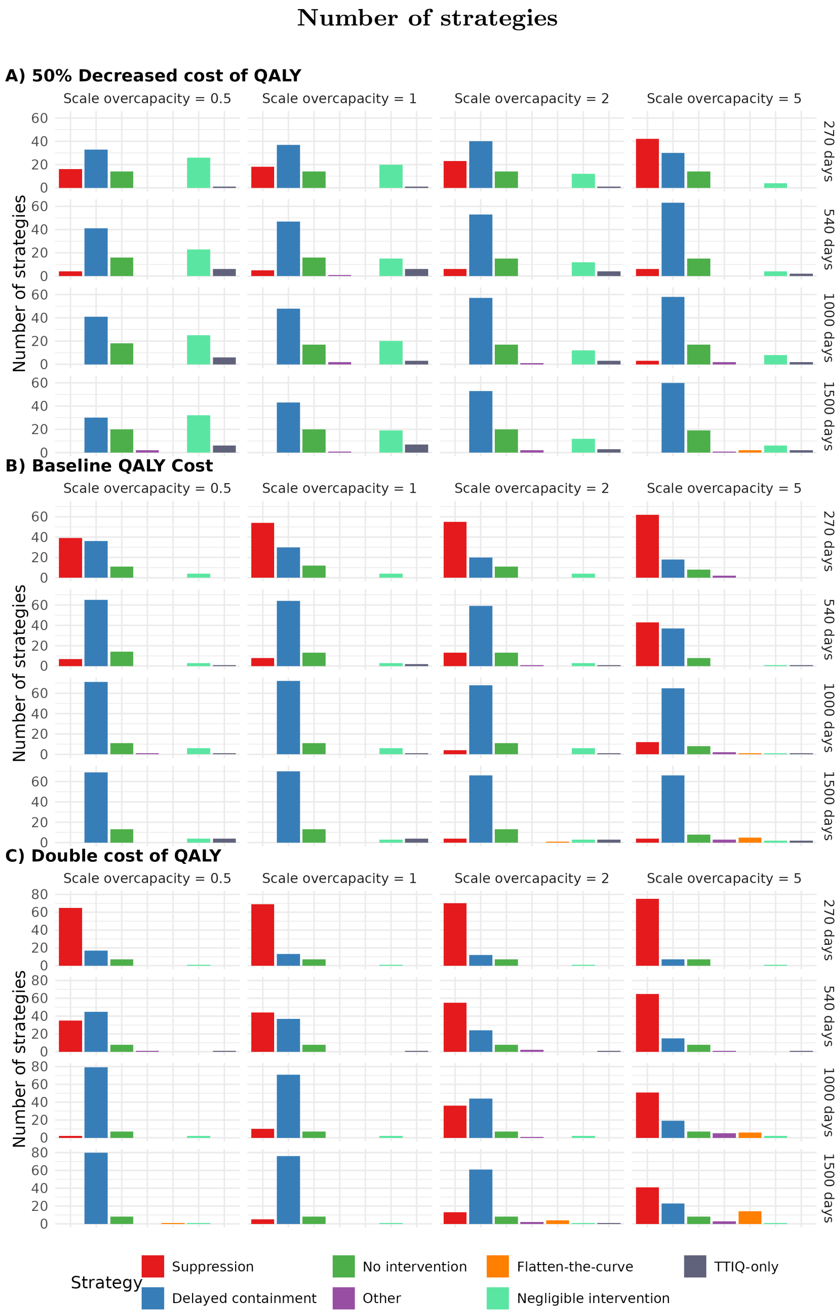

Figure S7: **Number of Strategies.** The number of various types of optimal strategies in Figure 4 in main paper, where strategy types follow the rule in Table S18. Parameter combinations as in Table S19.

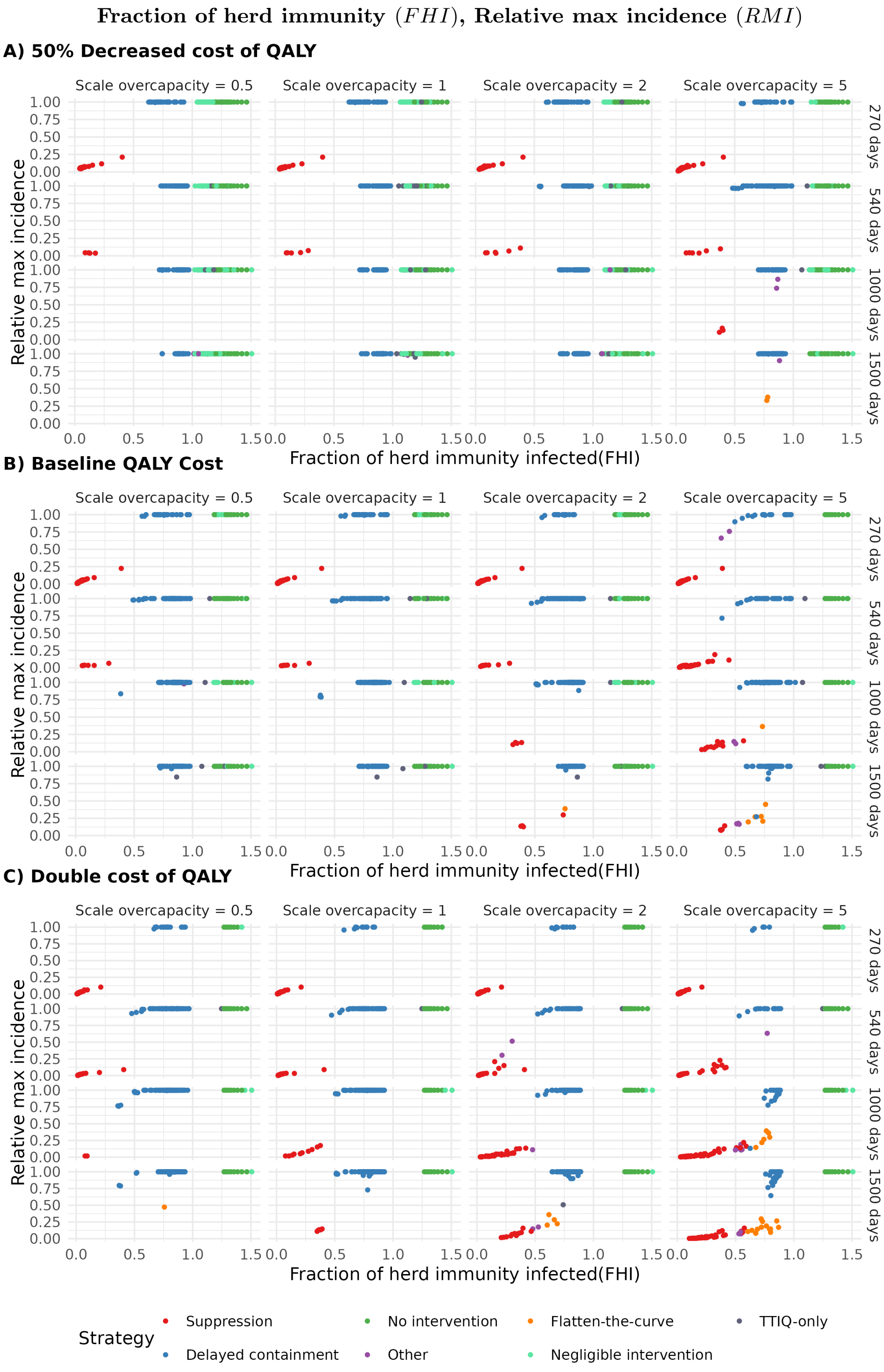

Figure S8: **Parameter scan.** Fraction of herd immunity $(FHI)$, and Relative Max Incidence $(RMI)$ for the optimal strategies presented in Figure 4 in main paper. Dots are classified into strategy types following the rule in Table S18, and parameter combinations as in Table S19.

#### Optimal strategies: $FHI$ and $ARI$

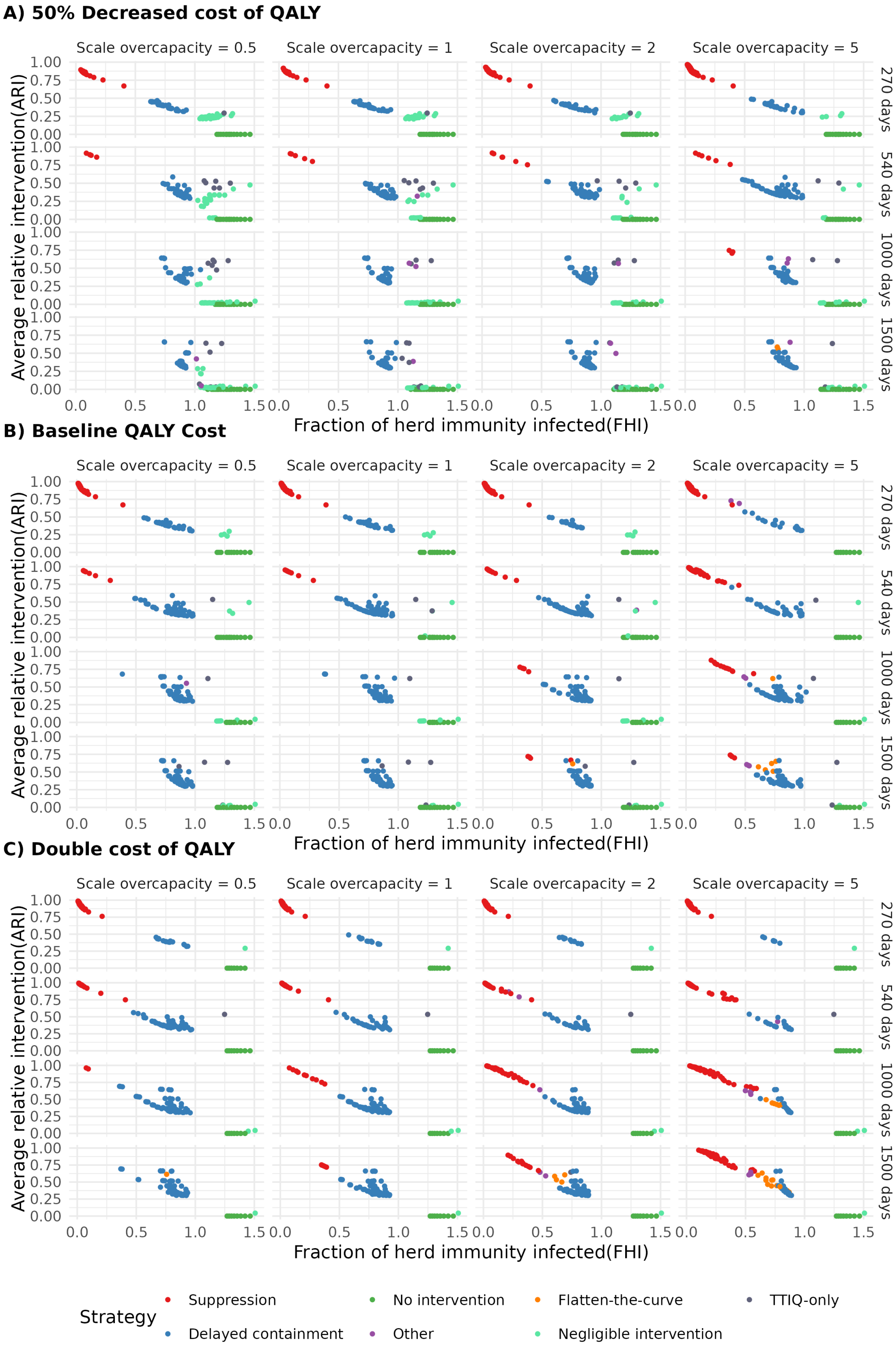

Figure S9: **Parameter scan.** Fraction of herd immunity ($FHI$) and Average relative intervention level ($ARI$) for the optimal strategies presented in Figure 4 in main paper. Dots are classified into strategy types following the rule in Table S18 and parameter combinations as in Table S19.

**Optimal strategies: $IR$ and $MINT$**

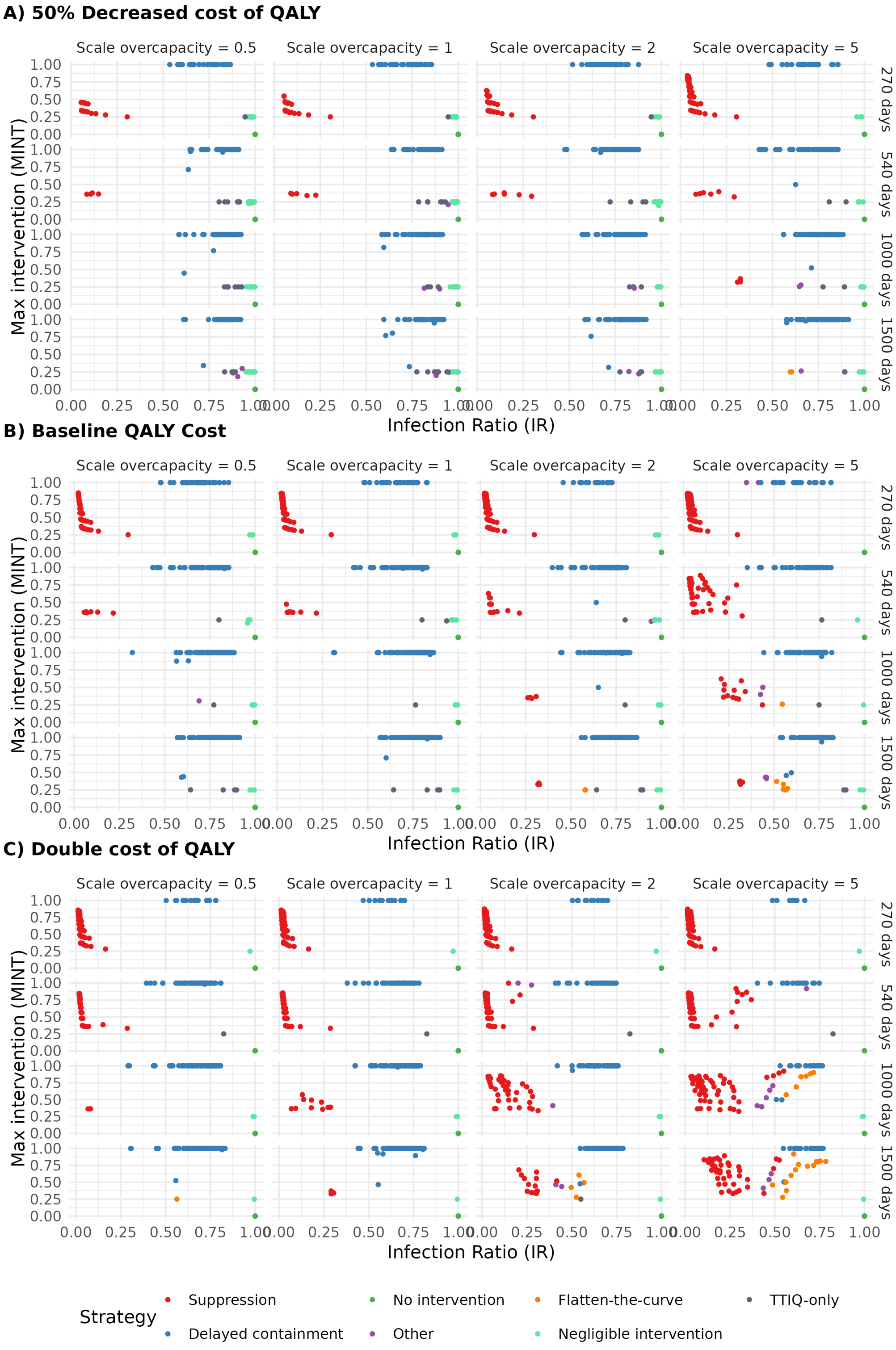

Figure S10: **Parameter scan.** Infection ratio ($IR$) and max intervention level $MINT$ for the optimal strategies in Figure 4 in main paper. Dots are classified into strategy types following the rule in Table S18, and parameter combinations as in Table S19.

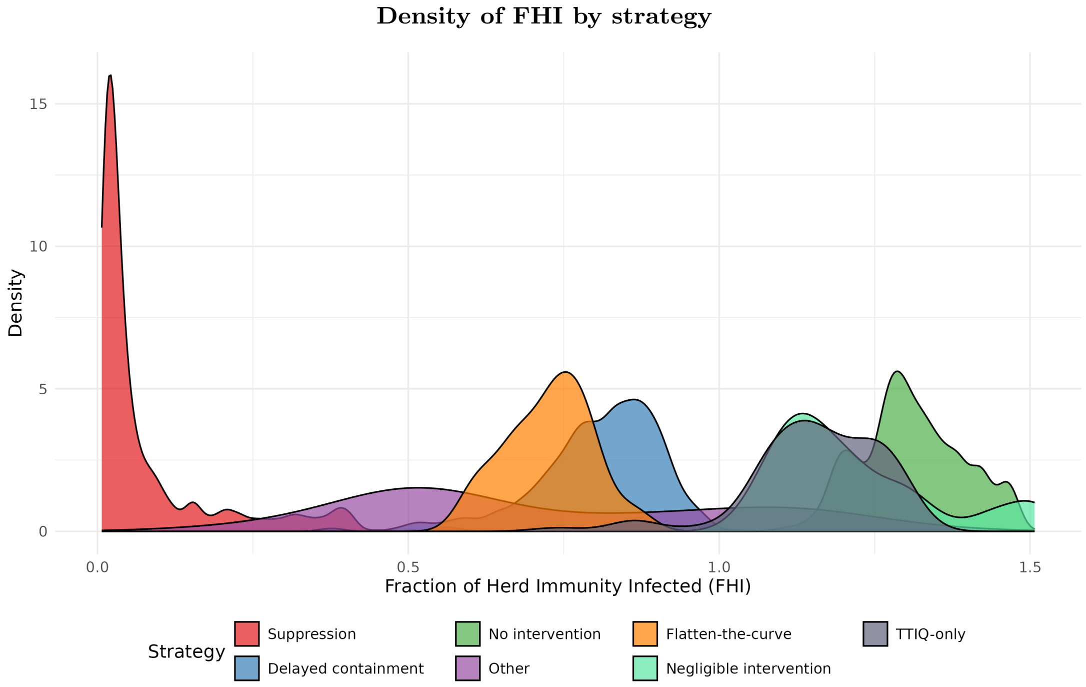

Figure S11: **Parameter scan.** Density of Fraction of Herd Immunity Infected(FHI) by strategy type for all strategies in Table S19. Note that the area for each strategy is set equal even if the number of cases of each strategy varies as seen in S7.

#### D.2 Examples of strategies

In this subsection we present examples of the strategy types, to illustrate some of the key characteristics. Each of figures S12–S17 illustrates a specific $(R, ES)$-combination in order to depict four trajectories: the optimal strategy (*Optimal*), a strategy without any interventions (*No Intervention*), an optimal suppression strategy, ie. the optimal strategy under the restriction that $ARI > 0.65$ and $RMI < 0.3$ (*Suppression*). The optimal *Delayed containment* strategy is derived under the conditions presented in section D.4 below, to ensure that the strategy is in line with the key properties of the delayed containment strategy.

Each figure has a 4×4 panel showing: (t-left) the amount of government interventions, (t-right) incidence of new infections, (b-left) prevalence in ICU and (b-right) cumulative fraction infected.

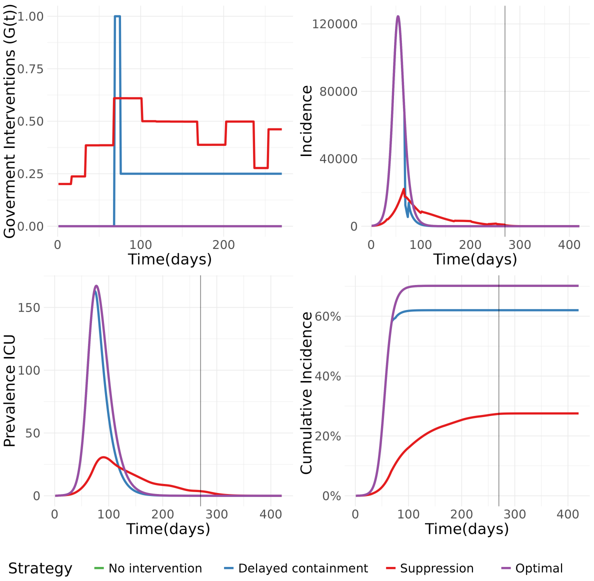

Figure S12: Combination: $R = 2$ and $ES = 1$ in baseline. Here *No intervention* is the optimal strategy. The shaded vertical line marks the introduction of the effective vaccine.

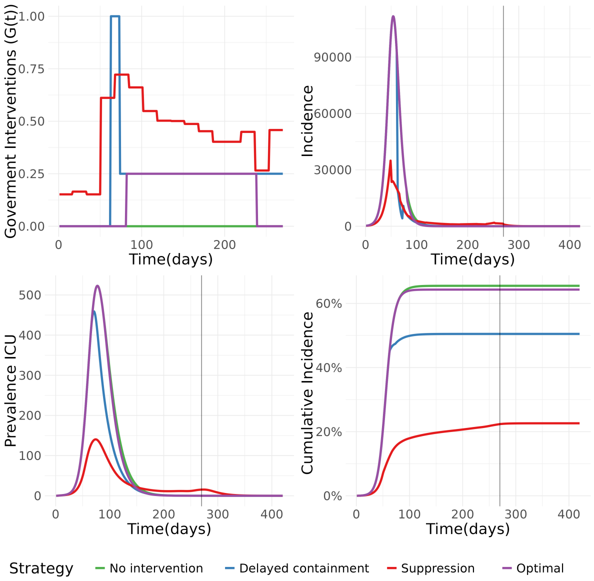

Figure S13: Combination: $R = 2$ and $ES = 3$ in baseline. Here *Negligible intervention* is the optimal strategy. The shaded vertical line marks the introduction of the effective vaccine.

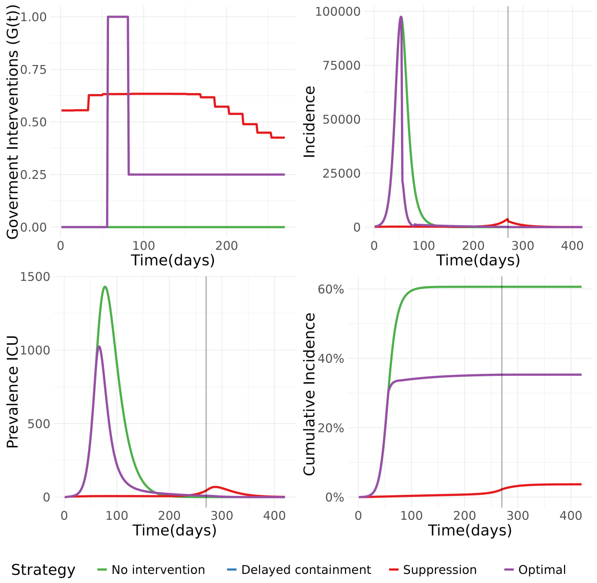

Figure S14: Combination: $R = 2.5$ and $ES = 7$ in baseline. Here *Delayed containment* is the optimal strategy. The shaded vertical line marks the introduction of the effective vaccine.

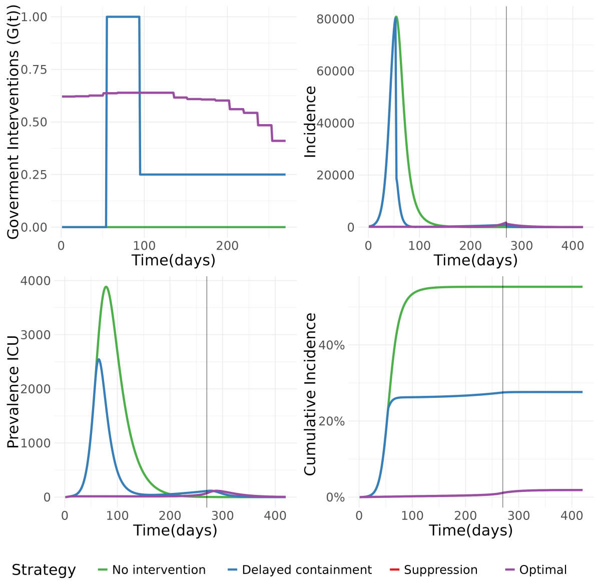

Figure S15: Combination: $R = 2$ and $ES = 15$ in baseline. Here *Suppression* is the optimal strategy. The shaded vertical line marks the introduction of the effective vaccine.

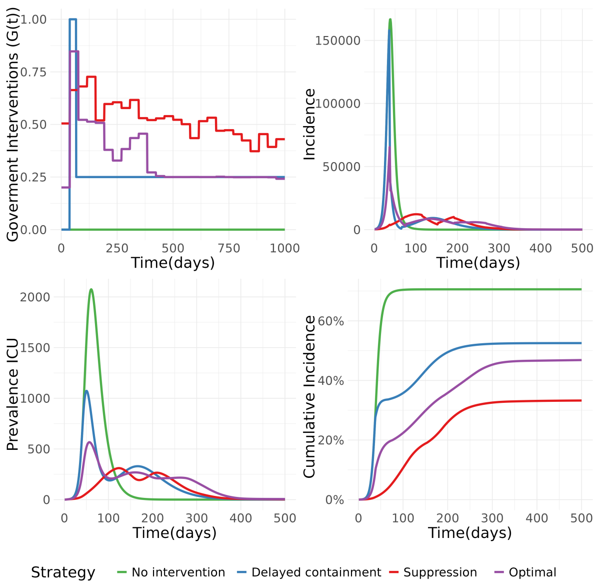

Figure S16: Combination: $R = 2.6$ and $ES = 7$ in a case with 1000 days to an effective vaccine, $2 \times COST_Q$ and $5 \times L_{HC}$. Here the optimal strategy is in the *Other* category. Note that we only show the first 500 days of the epidemic to highlight the interesting features.

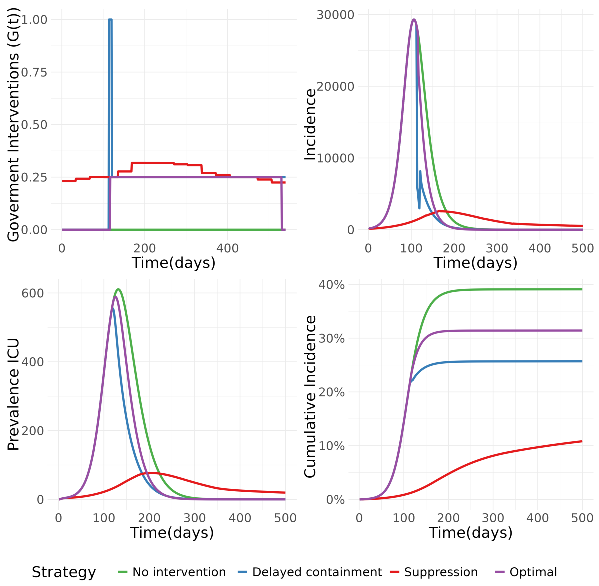

Figure S17: Combination: $R = 1.4$ and $ES = 7$ in a case with 540 days to an effective vaccine, $0.5 \times COST_Q$ and $0.5 \times L_{HC}$. Here *TTIQ-only* is the optimal strategy.

#### D.3 Relative cost of interventions

Figures S18-S19 explore what fraction of the total costs $K_T$ are due to interventions. Intervention costs are the total costs from contact reduction caused by government interventions and voluntary social distancing.

Figure S18 shows that under suppression, intervention costs are usually above 90 % of the total costs, but it may also be lower, in the interval 75-90 % for low R, when TTIQ is sufficient to contain the pandemic. In a very few cases intervention costs may also be below 75 % of total costs under suppression (for double QALY-cost, scale overcapacity = 5, vaccination after 1500 days, low $R$ and high $ES$). As intervention costs include costs associated with voluntary social distancing, these costs are also a large part of the total costs under no or negligible intervention. For high R, intervention costs including VSD constitute typically about 25-50 % of total costs under no/negligible intervention, and for lower $R$, intervention costs could be above 50 % of total costs. When *Delayed containment* is optimal there is larger variation in the fraction of intervention costs to total costs, varying from around 20 % to above 60 %.

Figure S19 shows the infection ratio ($IR$), defined as the fraction infected relative to the fraction infected with no interventions, and the intervention costs as a fraction of total costs, for the optimal strategies. *Suppression* strategies are omitted to facilitate a focus on the other strategies. *No/negligible intervention* have per definition almost no impact on the relative fraction of infected, but as noted above, intervention costs, including VSD, may still be a substantial part of total costs. *Delayed containment* leads to much larger reduction in the relative fraction of infected, with considerable variation depending on the parameter values.

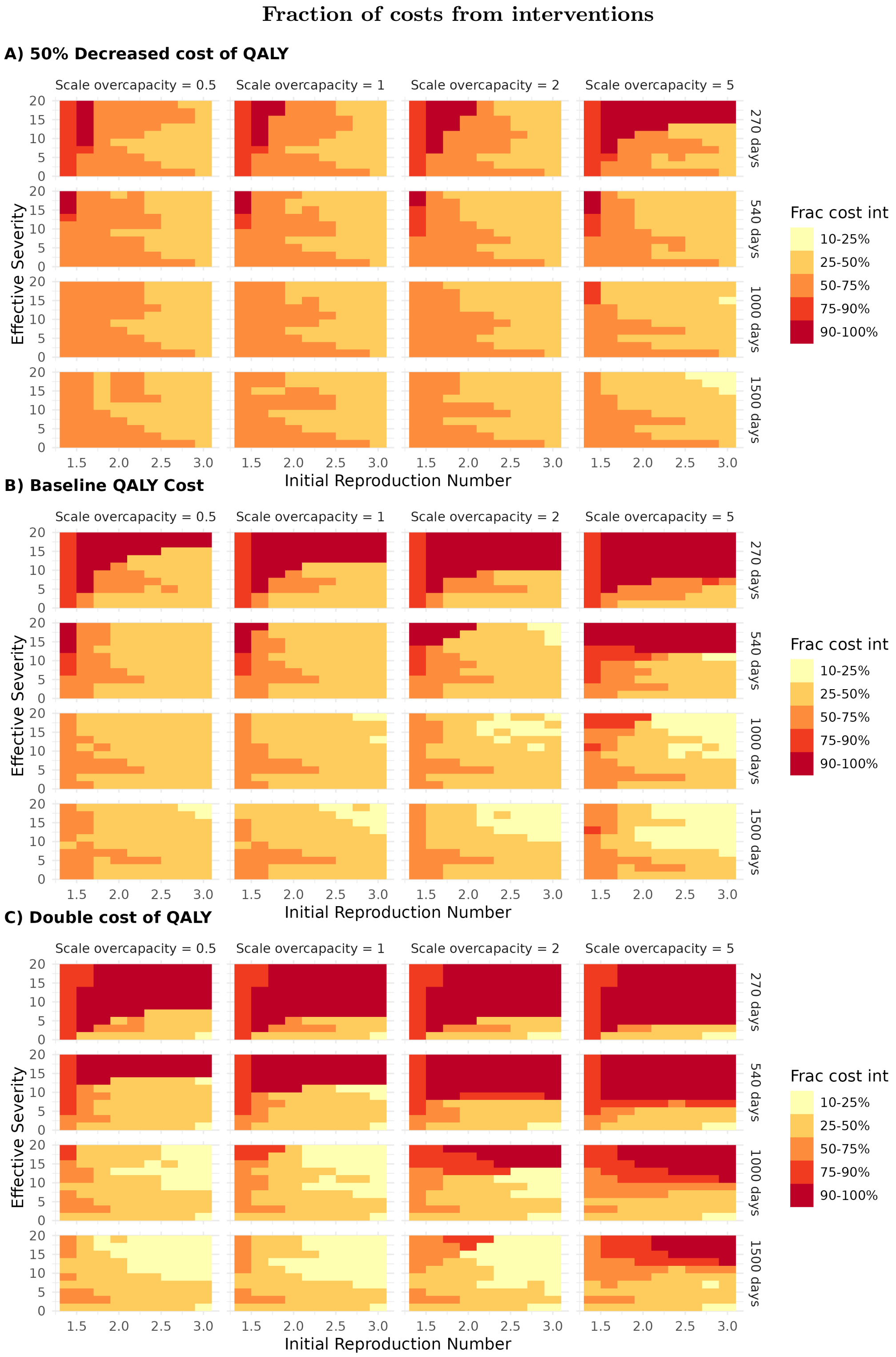

Figure S18: **Fraction of total costs due to interventions** for the optimal strategies in Figure 4 in the main paper. Parameter combinations in Table S19.

#### Optimal strategy: Fraction of costs from interventions and $(IR)$ $\tilde{I}/\tilde{I}_{\mathrm{Noi.}}$

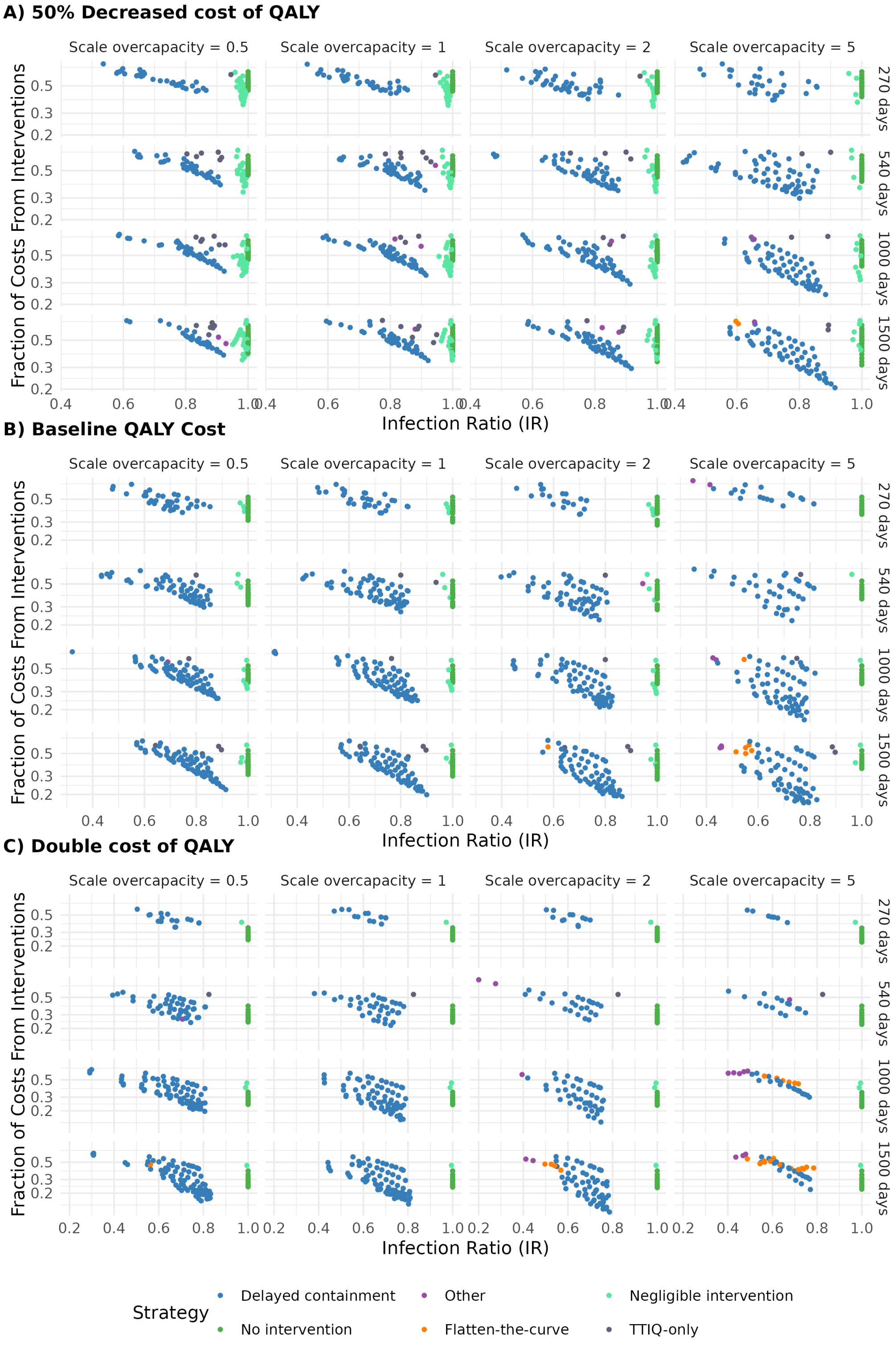

Figure S19: **Parameter space scan.** Infection ratio and the intervention costs as a fraction of total costs, for the optimal strategies in Figure 4 in the main paper. Suppression strategies are not included. Dots are classified into strategy types following the rule in Table S18. Parameter combinations as in Table S19.

#### D.4 Baseline case: Detailed output

Table S21 provides detailed output on economic losses and incidence for selected virus variants in the baseline case. For each $(R, ES)$-combination, the table compares the outcomes of five $G(t)$ trajectories, where all are optimal strategies under different restrictions, ensuring that they fall into the appropriate strategy categories:

1. Optimal: strategy from the optimization.
2. Suppression: $ARI > 0.65$.
3. No intervention: $G(t) = 0$.
4. Delayed containment: Grid search over delayed containment strategies, see Section A.4. Strict intervention, $G(t) = 1$, must start within 20 days of the uncontrolled peak and last 7–40 days.[13]
5. ICU cap: subject to the constraint $ICU \leq \overline{ICU}$.

| $R$ | $ES$ | $G(t)$ | $L_T$ † | $L_Q$ | $L_{HC}$ | $L_{\Delta G}$ | $L_{SICK}$ | $K_T$ | $L_{TTIQ}$ | ARI | FHI | hosp | ICU | deaths | $ICU^{peak}$ $/\overline{ICU}$ § |
|---|---|---|---|---|---|---|---|---|---|---|---|---|---|---|---|
| | | | measured in bnNOK | | | | | | | | | per 100 000 | | | |
| 1.6 | 1 | Optimal | 65 | 25 | 0 | 0 | 6 | 34 | 0 | 0.00 | 146 | 92 | 9 | 30 | 0.30 |
| 1.6 | 1 | Suppression | 98 | 9 | 0 | 6 | 2 | 63 | 18 | 0.72 | 44 | 32 | 3 | 12 | 0.04 |
| 1.6 | 1 | No int. | 65 | 25 | 0 | 0 | 6 | 34 | 0 | 0.00 | 146 | 92 | 9 | 30 | 0.30 |
| 1.6 | 1 | Delay. cont. | 75 | 20 | 0 | 6 | 5 | 41 | 4 | 0.39 | 119 | 72 | 7 | 23 | 0.29 |
| 1.6 | 1 | ICU cap | 65 | 25 | 0 | 0 | 6 | 34 | 0 | 0.00 | 146 | 92 | 9 | 30 | 0.30 |
| 1.6 | 7 | Optimal | 118 | 12 | 0 | 4 | 0 | 97 | 4 | 0.88 | 6 | 42 | 8 | 15 | 0.16 |
| 1.6 | 7 | Suppression | 118 | 12 | 0 | 4 | 0 | 97 | 4 | 0.88 | 6 | 42 | 8 | 15 | 0.16 |
| 1.6 | 7 | No int. | 173 | 82 | 22 | 0 | 5 | 64 | 0 | 0.00 | 128 | 424 | 96 | 105 | 2.63 |
| 1.6 | 7 | Delay. cont. | 132 | 32 | 3 | 6 | 2 | 86 | 4 | 0.56 | 44 | 154 | 34 | 42 | 1.39 |
| 1.6 | 7 | ICU cap | 118 | 12 | 0 | 4 | 0 | 97 | 4 | 0.87 | 6 | 41 | 8 | 15 | 0.17 |
| 1.6 | 15 | Optimal | 126 | 14 | 0 | 4 | 0 | 105 | 3 | 0.91 | 3 | 47 | 16 | 17 | 0.32 |
| 1.6 | 15 | Suppression | 126 | 14 | 0 | 4 | 0 | 105 | 3 | 0.91 | 3 | 47 | 16 | 17 | 0.32 |
| 1.6 | 15 | No int. | 353 | 135 | 130 | 0 | 5 | 83 | 0 | 0.00 | 116 | 759 | 292 | 178 | 7.07 |
| 1.6 | 15 | Delay. cont. | 177 | 44 | 13 | 6 | 1 | 110 | 4 | 0.65 | 27 | 212 | 80 | 59 | 2.58 |
| 1.6 | 15 | ICU cap | 126 | 14 | 0 | 4 | 0 | 105 | 3 | 0.91 | 3 | 47 | 16 | 17 | 0.32 |
| 2.0 | 1 | Optimal | 70 | 31 | 0 | 0 | 7 | 32 | 0 | 0.00 | 138 | 112 | 11 | 34 | 0.48 |
| 2.0 | 1 | Suppression | 167 | 17 | 0 | 5 | 4 | 110 | 32 | 0.69 | 67 | 60 | 6 | 22 | 0.12 |
| 2.0 | 1 | No int. | 70 | 31 | 0 | 0 | 7 | 32 | 0 | 0.00 | 138 | 112 | 11 | 34 | 0.48 |
| 2.0 | 1 | Delay. cont. | 81 | 26 | 0 | 6 | 7 | 39 | 4 | 0.33 | 122 | 95 | 10 | 28 | 0.47 |
| 2.0 | 1 | ICU cap | 70 | 31 | 0 | 0 | 7 | 32 | 0 | 0.00 | 138 | 112 | 11 | 34 | 0.48 |
| 2.0 | 7 | Optimal | 168 | 55 | 14 | 6 | 4 | 84 | 5 | 0.43 | 70 | 292 | 64 | 74 | 2.93 |
| 2.0 | 7 | Suppression | 209 | 13 | 0 | 5 | 0 | 187 | 4 | 0.92 | 4 | 44 | 9 | 16 | 0.18 |
| 2.0 | 7 | No int. | 206 | 99 | 40 | 0 | 6 | 60 | 0 | 0.00 | 120 | 522 | 116 | 133 | 4.09 |
| 2.0 | 7 | Delay. cont. | 168 | 55 | 14 | 6 | 4 | 84 | 5 | 0.43 | 70 | 292 | 64 | 74 | 2.93 |
| 2.0 | 7 | ICU cap | 209 | 13 | 0 | 6 | 0 | 186 | 4 | 0.92 | 5 | 45 | 9 | 16 | 0.20 |
| 2.0 | 15 | Optimal | 217 | 15 | 0 | 5 | 0 | 194 | 3 | 0.94 | 2 | 52 | 18 | 19 | 0.37 |
| 2.0 | 15 | Suppression | 217 | 15 | 0 | 5 | 0 | 194 | 3 | 0.94 | 2 | 52 | 18 | 19 | 0.37 |
| 2.0 | 15 | No int. | 417 | 169 | 161 | 0 | 6 | 81 | 0 | 0.00 | 109 | 954 | 355 | 237 | 11.10 |
| 2.0 | 15 | Delay. cont. | 276 | 80 | 67 | 6 | 3 | 116 | 4 | 0.49 | 54 | 456 | 168 | 111 | 7.27 |
| 2.0 | 15 | ICU cap | 217 | 14 | 0 | 5 | 0 | 195 | 3 | 0.94 | 2 | 50 | 17 | 18 | 0.36 |
| 2.6 | 1 | Optimal | 74 | 36 | 0 | 0 | 9 | 29 | 0 | 0.00 | 131 | 130 | 14 | 39 | 0.68 |
| 2.6 | 1 | Suppression | 233 | 22 | 0 | 7 | 5 | 159 | 41 | 0.68 | 74 | 79 | 8 | 27 | 0.22 |
| 2.6 | 1 | No int. | 74 | 36 | 0 | 0 | 9 | 29 | 0 | 0.00 | 131 | 130 | 14 | 39 | 0.68 |
| 2.6 | 1 | Delay. cont. | 86 | 32 | 0 | 6 | 8 | 36 | 4 | 0.29 | 121 | 116 | 12 | 34 | 0.65 |
| 2.6 | 1 | ICU cap | 74 | 36 | 0 | 0 | 9 | 29 | 0 | 0.00 | 131 | 130 | 14 | 39 | 0.68 |
| 2.6 | 7 | Optimal | 211 | 74 | 29 | 6 | 5 | 91 | 6 | 0.38 | 77 | 397 | 85 | 102 | 4.35 |
| 2.6 | 7 | Suppression | 299 | 58 | 13 | 12 | 4 | 182 | 31 | 0.68 | 53 | 295 | 65 | 78 | 2.27 |
| 2.6 | 7 | No int. | 244 | 123 | 58 | 0 | 7 | 56 | 0 | 0.00 | 113 | 638 | 139 | 176 | 5.93 |
| 2.6 | 7 | Delay. cont. | 211 | 74 | 29 | 6 | 5 | 91 | 6 | 0.38 | 77 | 397 | 85 | 102 | 4.35 |
| 2.6 | 7 | ICU cap | 308 | 34 | 2 | 9 | 2 | 247 | 14 | 0.84 | 24 | 159 | 34 | 48 | 1.00 |
| 2.6 | 15 | Optimal | 318 | 14 | 0 | 5 | 0 | 297 | 2 | 0.97 | 2 | 48 | 17 | 18 | 0.31 |
| 2.6 | 15 | Suppression | 318 | 14 | 0 | 5 | 0 | 297 | 2 | 0.97 | 2 | 48 | 17 | 18 | 0.31 |
| 2.6 | 15 | No int. | 497 | 219 | 195 | 0 | 7 | 77 | 0 | 0.00 | 104 | 1192 | 431 | 326 | 16.62 |
| 2.6 | 15 | Delay. cont. | 368 | 128 | 112 | 6 | 5 | 113 | 5 | 0.41 | 70 | 727 | 258 | 184 | 12.43 |
| 2.6 | 15 | ICU cap | 321 | 16 | 0 | 7 | 0 | 296 | 3 | 0.96 | 2 | 58 | 20 | 21 | 0.35 |

† From equation (17): $L_T = L_Q + L_{HC} + L_{\Delta G} + L_{SICK} + K_T + L_{TTIQ}$
§ Peak number of patients at ICU, relative to max capacity $\overline{ICU}$.

Table S21: Baseline case: Detailed output for virus variants with $R = \{1.6, 2.0, 2.6\}$ and $ES = \{1, 7, 15\}$

[13] We use a more restrictive rule than in section D.1.1 to ensure that the strategy exhibit the key features of a *Delayed containment*-strategy.

#### D.5 Overcapacity costs (Fig S20 )

To illustrate the implications of our assumptions concerning excess costs from hospital overcapacity, Figure S20 shows the overcapacity cost, $L_{HC}$, relative the sum of all other costs in terms of health, economy and welfare ($K_T, L_{\Delta G}$,$L_Q$,$L_{SICK}$), under the assumption that no interventions are implemented. Under the baseline assumptions, overcapacity costs may exceed 50 % of the other cost components at high severity levels. When the scale parameter for overcapacity is set to $s_o = 2$, overcapacity costs may be greater than the other costs for baseline QALY cost and high severity, and more than three times other costs for double QALY cost and high severity. For scale of overcapacity costs $s_o = 5$, overcapacity costs may be more than seven times the other costs for double QALY cost and high severity.

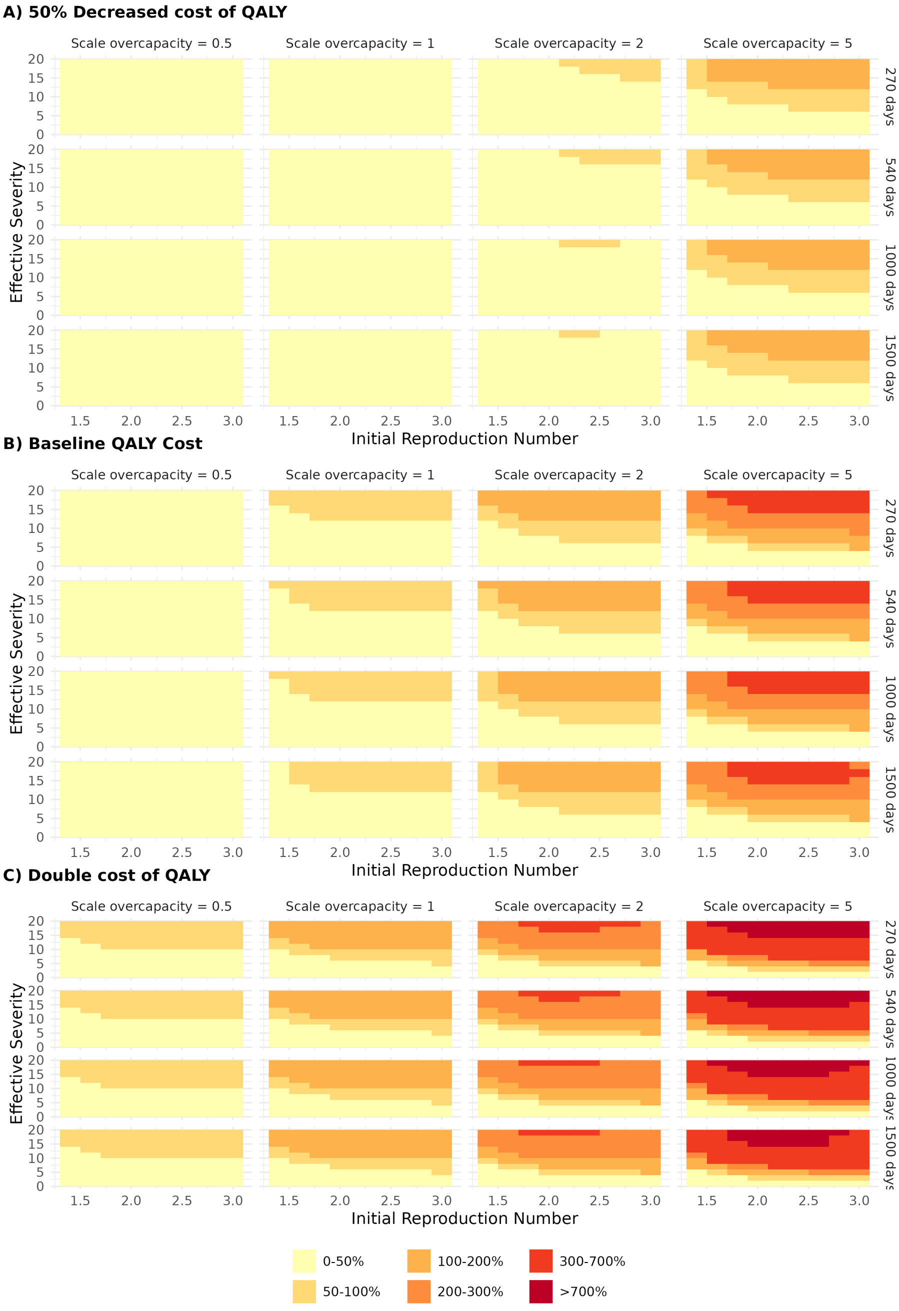

Figure S20: The figure shows the excess costs coming from overcapacity in hospitals, relative to all other costs. Parameter combinations as in Table S19.

#### D.6 Optimal strategy by severity (Fig S21)

In this subsection we illustrate the mechanics of the model by displaying how the different strategy types depend on the effective severity ($ES$) of the virus. To do this, we calculate the optimal intervention policy conditional on each strategy type as defined in section D.4.

The left panel shows total costs ($L_T$), the mid panel the average relative intervention ($ARI$) and the right panel the fraction of infected over the duration of the pandemic.

Consistent with the results in Figure 2.A in main paper, we observe that the unconditional optimal strategy varies from *No intervention* for low severity, *Delayed containment* for medium severity and *Suppression* for high severity. Note that the contingent optimal strategies imply that interventions and infections change smoothly with severity, while the unconditional optimal strategy involves clear jumps in intervention level and infection outcome when there is a change in the type of strategy. Note also that the ICU-cap strategy involves much higher costs than the optimal strategy, except for low severity when a *No interventions* strategy do not exceed hospital capacity, thus formally satisfying the criteria for the ICU-cap.

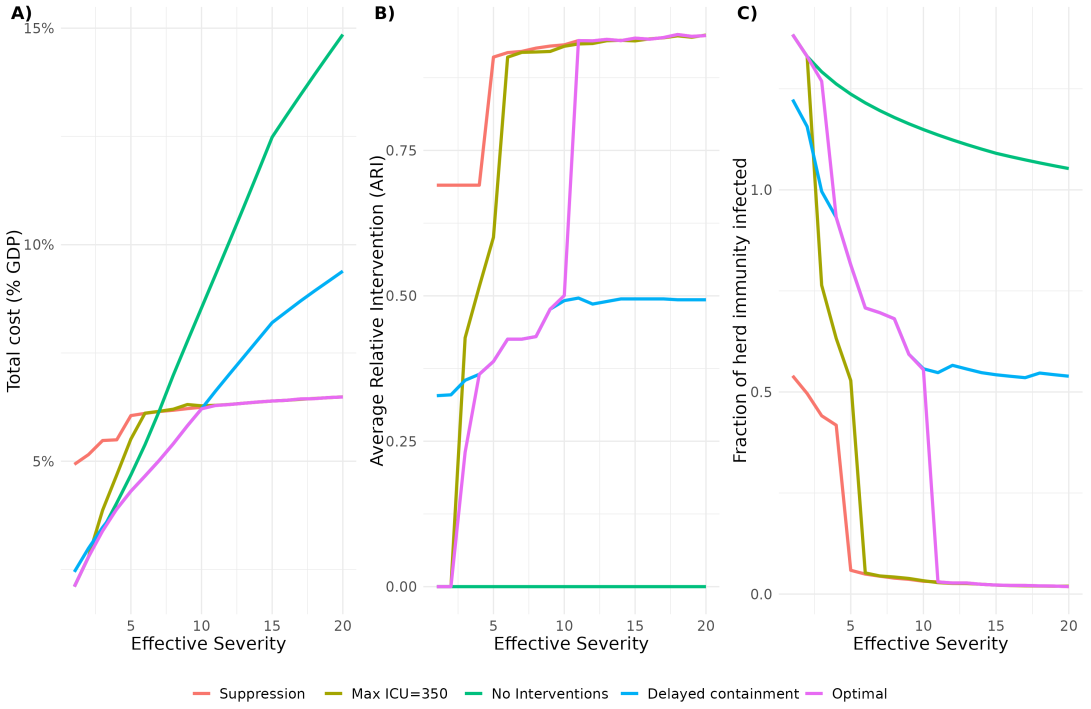

Figure S21: **The effect of increasing severity on the various strategy types.** The lines represent optimal strategies conditional on the type of strategy. Suppression requires that the average relative intervention is above 0.65 and the RMI below 0.3. Delayed containment is the optimal delayed containment strategy as defined by in Section D.4. The purple lines indicate the unconditional optimal strategy. **Panel A:** Total costs **Panel B:** Average relative intervention (ARI) **Panel C:** Fraction infected relative to the herd immunity level (FHI)

#### D.7 No TTIQ (Fig S22)

To explore the robustness of our results under the broad parameter scan presented in Figure 4 in the main paper, Figure S22 displays the optimal strategy if Test-Trace-Isolate-Quarantine (TTIQ) is not available.

This raises the costs of interventions, in particular for low initial values of $R$ where TTIQ would have been sufficient to keep the effective $R$ below unity. For low QALY-costs, this makes no intervention the dominant strategy choice for low to moderate severity, while *Delayed containment* remains optimal in most cases with high severity. For baseline ($1 \times COST_Q$) and double QALY cost ($2 \times COST_Q$), the optimal strategy remains the same as when TTIQ is feasible in the large majority of cases.

Thus, removing the possibility of TTIQ does not affect the key results that the optimal strategy is typically either *Suppression*, *Delayed containment* or *No/negligible intervention*, and there are only a few cases of *Flatten-the-curve* for long duration and high overcapacity costs.

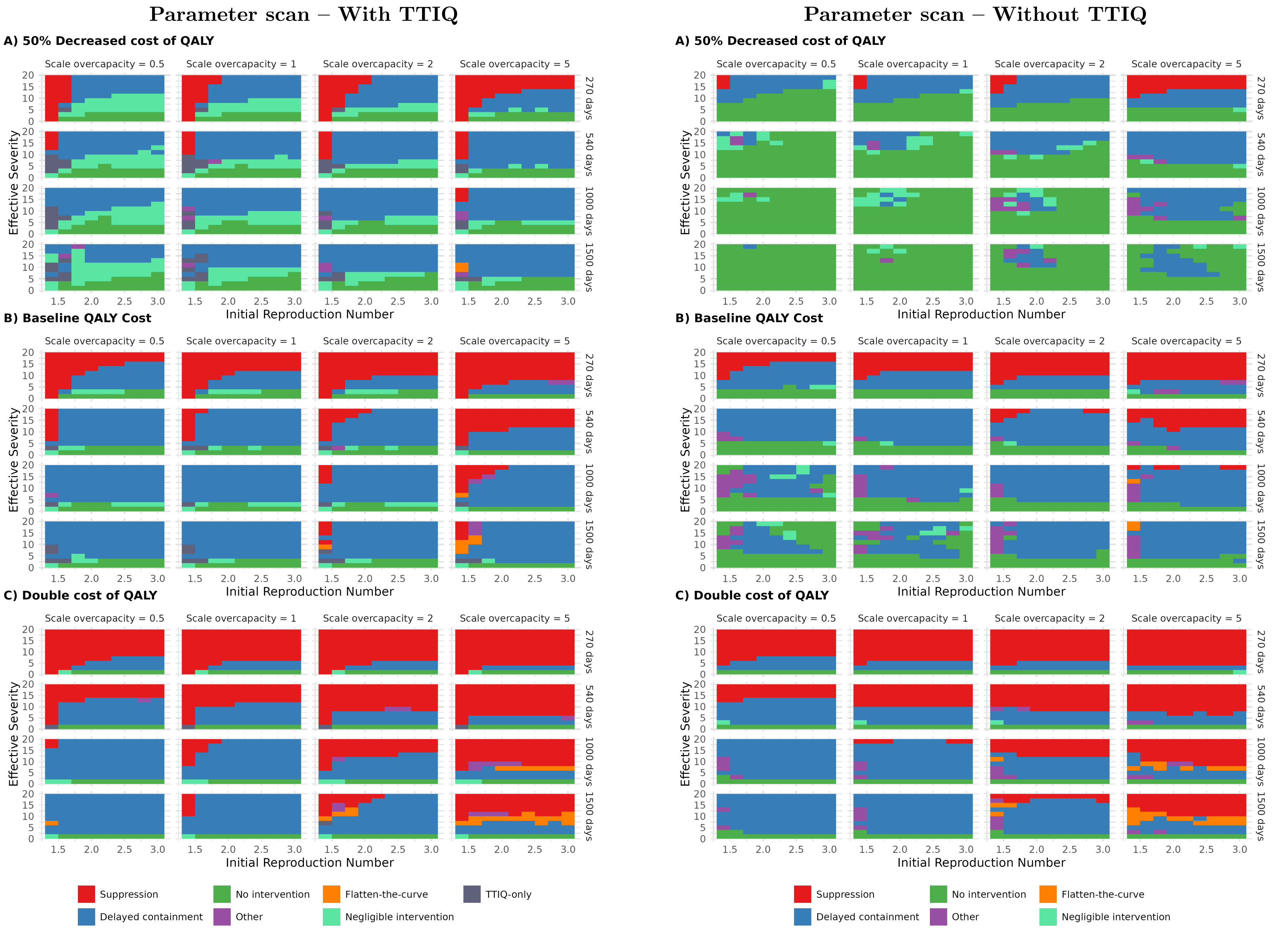

Figure S22: **Sensitivity analysis: NO TTIQ** Optimal strategies in the broad parameter scan, under different assumptions: With TTIQ (left) is identical to Figure 4 in the main paper; without TTIQ (right). **A:** 50% decrease cost of QALY. **B:** Baseline QALY cost. **C:** Double costs of QALY

#### D.8 Policy mistakes (Fig S23-S25)

In this subsection we consider the increase in costs from choosing a strategy that is not the cost-minimizing strategy. Choosing a policy which is not the cost-minimizing may be an error - a policy mistake - or it could be the result of a deliberate choice based on a motivation that is not captured by our loss function or optimization procedure. Irrespective of the reason, it is useful to have information about the consequences for total societal costs from choosing a specific strategy. The figures may also be useful if there is uncertainty about the properties of the virus or the society's preferences or capacity to handle the pandemic.

Figures S23, S24 and S25 show the additional cost of always choosing either *Delayed containment*, *Suppression* or *No intervention* across all the cases in the parameter scan.

Figure S23 shows the increase in total costs from always choosing *Delayed containment*, relative to the costs under the optimal strategy. Light yellow indicates that *Delayed containment* is optimal. We observe that in the majority of scenarios, *Delayed containment* is either optimal or it involves an increase in costs of less than 10 %. For low severity and/or low R, total costs are low under *No/Negligible intervention*, and the relative cost increase from *Delayed containment* is greater. For scenarios with high severity and short time to vaccination (270 or 540 days), in particular for high value of a QALY, *Delayed containment* may involve much greater increase in total costs, even more than 600 % increase in some scenarios indicated by dark red color.

Figures S24 and S25 show the increase in costs from always choosing *Suppression* and *No intervention*, respectively. These strategies lead to a larger increase in relative costs for a larger set of scenarios. In particular, this applies to *Suppression*, which involves a stark relative increase in total costs for low severity levels and low QALY-value, while *No intervention* results in greater relative increase in costs for high severity and high QALY-value. Note however that under optimal policy, total costs are strongly increasing in severity and the QALY-value, implying that *Suppression* leads to larger relative increase when total costs are low, while *No intervention* leads to larger relative increase when total costs are high. Comparing the effect of policy mistakes on the absolute increase in total costs might therefore give a different picture. A further study of the costs of policy mistakes should therefore consider the absolute increase in total costs.

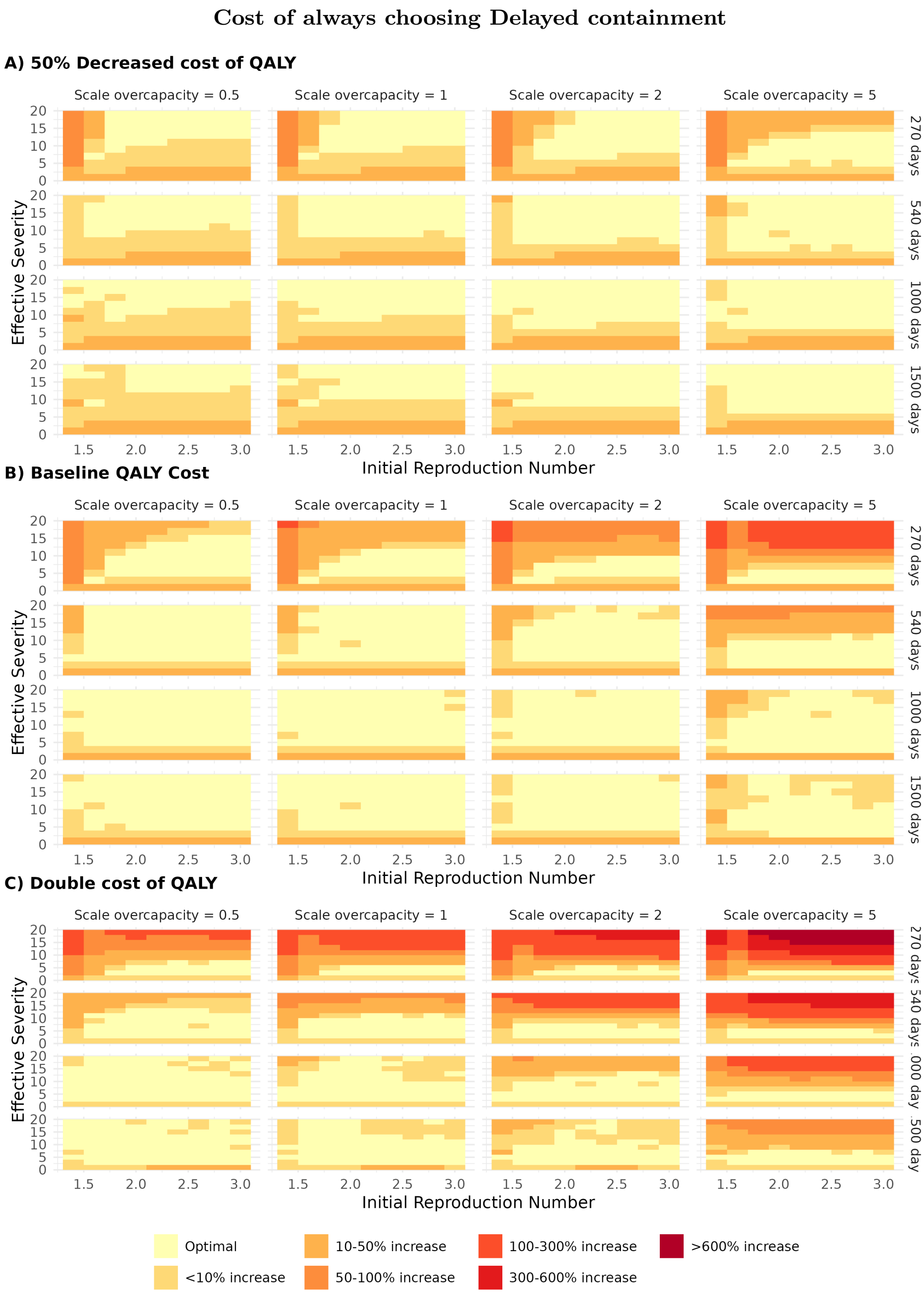

Figure S23: **Relative increase in costs** from choosing the optimal *Delayed containment* strategy, as defined in Section D.4, compared to the globally optimal strategy.

### Cost of always choosing Suppression

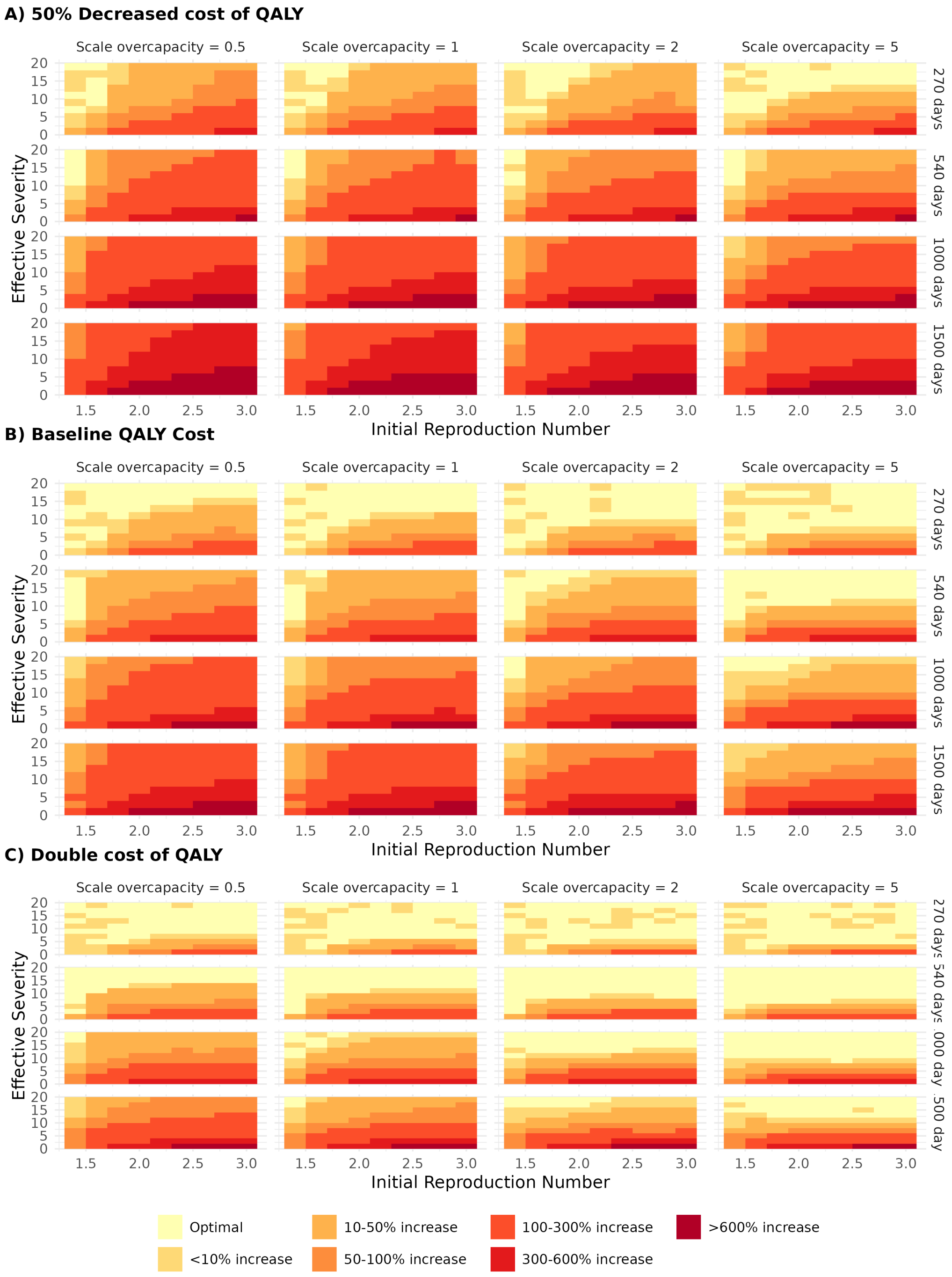

Figure S24: **Relative increase in costs** from choosing the optimal *Suppression* strategy, as defined in Section D.4, compared to the globally optimal strategy.

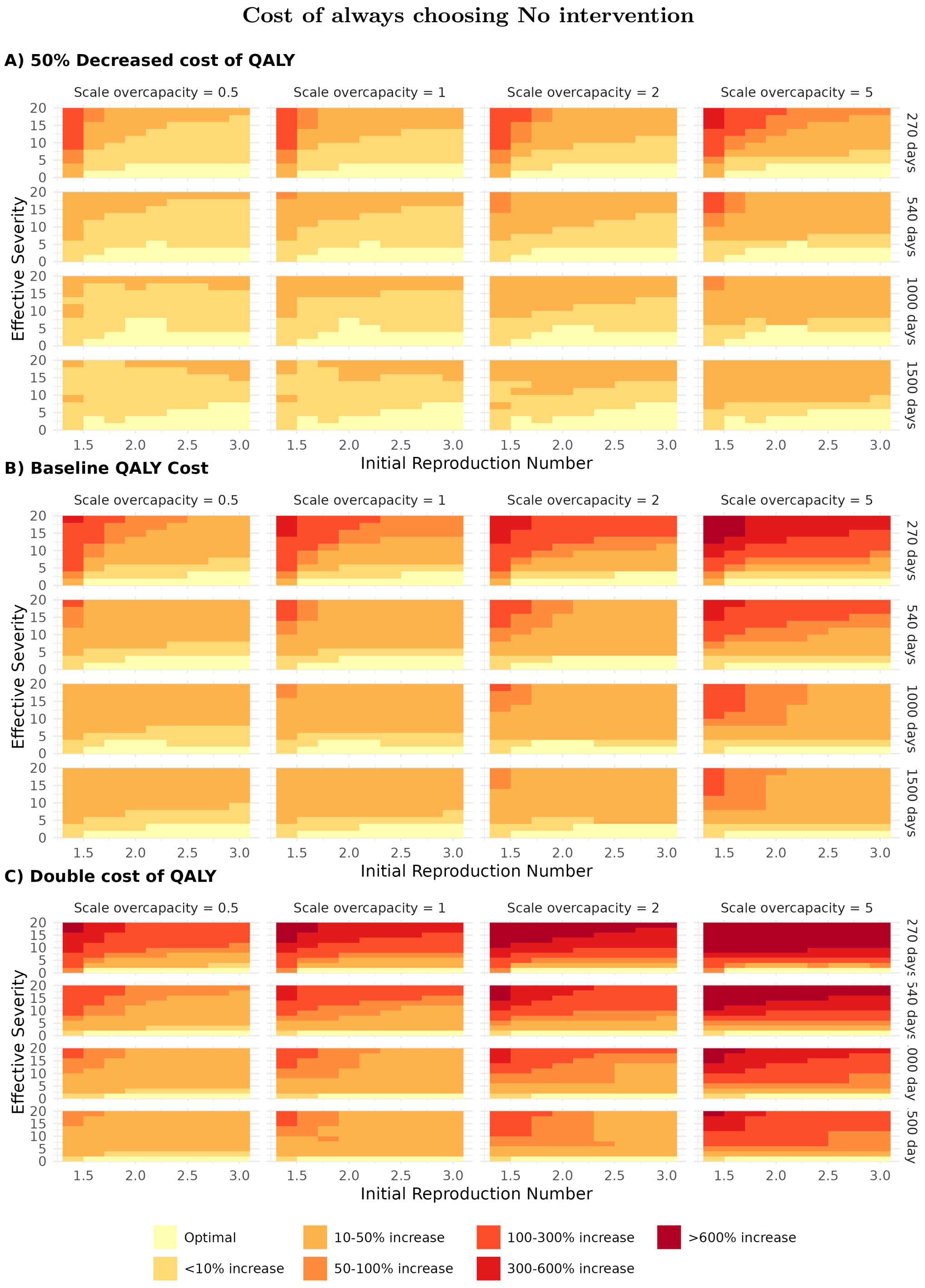

Figure S25: **Relative increase in costs** from choosing the *No intervention* strategy, as defined in Section D.4, compared to the globally optimal strategy

.

##### D.8.1 Strict ICU cap (Fig S26)

We have seen that *flatten-the-curve* policies are optimal only under narrow circumstances, in scenarios with long time until vaccination and very high excess costs of overwhelmed hospitals. However, the experience from the Covid-19 pandemic suggests that *flatten-the-curve* strategies are more common in practice, as protection of the health sector has been a major motivation for imposing strict interventions. This may reflect that an overburdened health sector, where media report on patients dying in crowded hospitals, might involve substantial political costs which are not included in our simulations.

The simulations in this paper show that *flatten-the-curve* policies may come with high costs. Figure S26 presents the increase in total societal costs from imposing a strict ICU constraint, where interventions are sufficiently strict to keep the number of patients below the ICU limit at all times.

The additional costs from imposing a strict limit on ICU capacity vary sharply depending on the parameter values. For the baseline case, with scale of overcapacity equal to 1 and 270 days until vaccination, total costs may increase by up to 81 % for high R-values and low severity. For low QALY-cost ($0.5 \times COST_Q$), delayed vaccination, low overcapacity cost ($0.5 \times L_{HC}$) and high R-values, imposing a strict ICU limit may increase total societal costs by a multiple of 4 to 7 times. For high QALY-cost ($2 \times COST_Q$) and high overcapacity costs ($5 \times L_{HC}$), the relative increase is smaller. However, as the total costs are very high under these circumstances, the absolute increase in total costs from imposing a strict ICU limit may still be very high.

#### Imposing a strict ICU cap

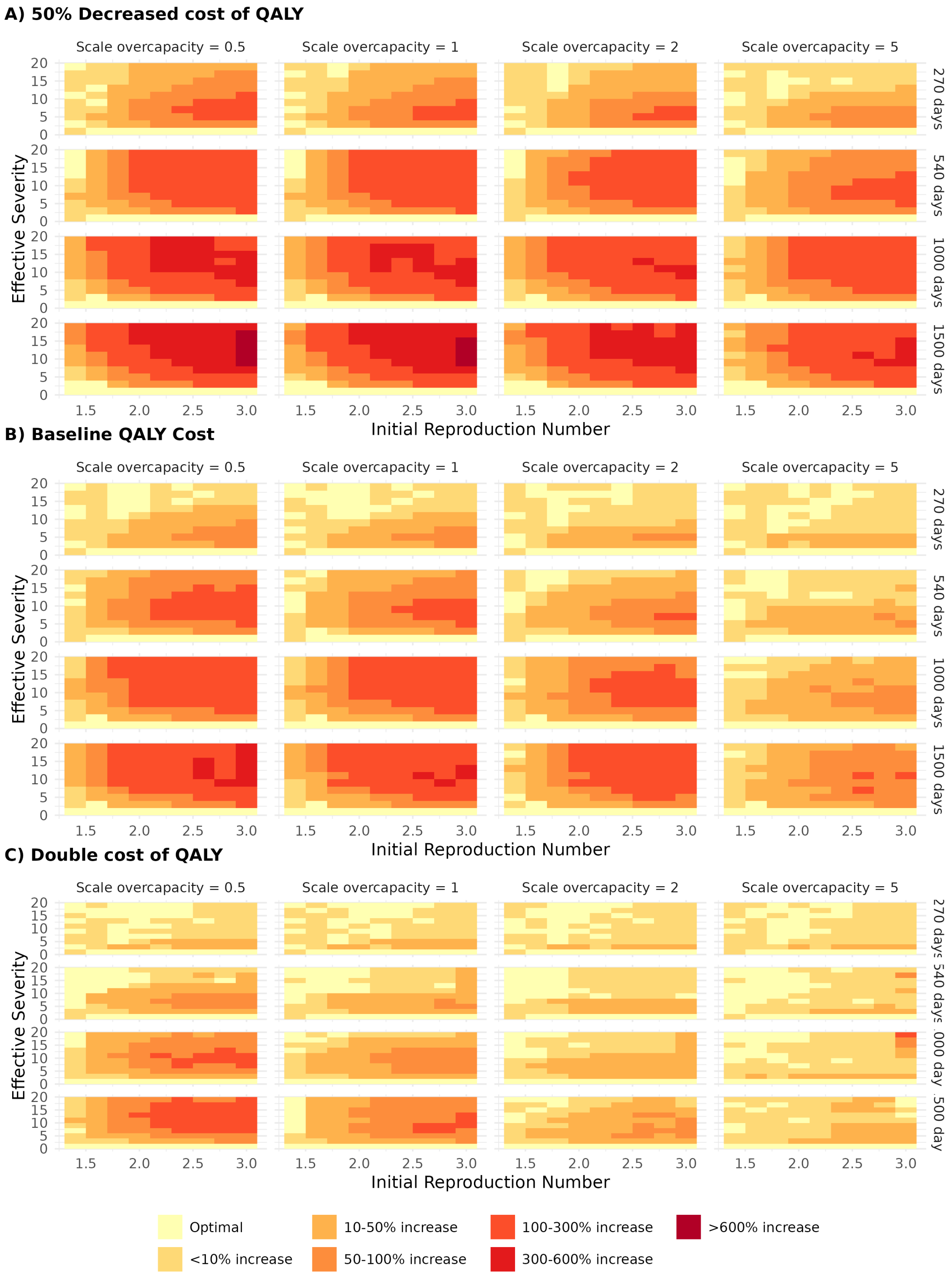

Figure S26: **Increase in costs from a ICU constraint.** Relative increase in costs of imposing a strict limit on 350 patients in the ICU for all parameters in the parameter space scan, compared to the costs under the optimal strategy.

#### D.9 Distributional effects (Fig S27)

We define the optimal strategy as the one that minimizes total societal costs. However, because Covid-19 risks have a high age-gradient, strategy choice has important distributional implications. This is illustrated in Figures S27.A and B, which show total costs by age group and per person within each age group for different strategy types, assuming that $R = 2.4$ and $ES = 12$. The difference across age reflects that under *Suppression*, infections are rare, so costs vary little across age groups aside from the residual health risk borne disproportionally by older individuals. Under *Delayed containment* or *No intervention*, higher infection rates impose much larger health costs and VSD-costs on older groups, while younger groups benefit from fewer and/or shorter contact-reducing interventions.

To highlight a potential policy dilemma, we focus on an $(R, ES)$-combination where *Suppression* is optimal and other strategies yield higher total costs. However, the unequal distribution of health costs under *Delayed containment* implies that a majority of the voters (here, those in the age from 18 to 50), would privately prefer *Delayed containment* to *Suppression*. In such cases, individual preferences leading to *Delayed containment* may conflict with the utilitarian optimum where total costs are minimized under *Suppression*.

Given the structure of our model, it is difficult to examine other types of distributional consequences, like differences across groups depending on socio-economic or health-related characteristics. Empirically, both disease burdens and intervention costs were unevenly distributed across groups [55, 56]. Exposure also varied by occupation and sector, and the economic incidence was also correspondingly heterogeneous [57].

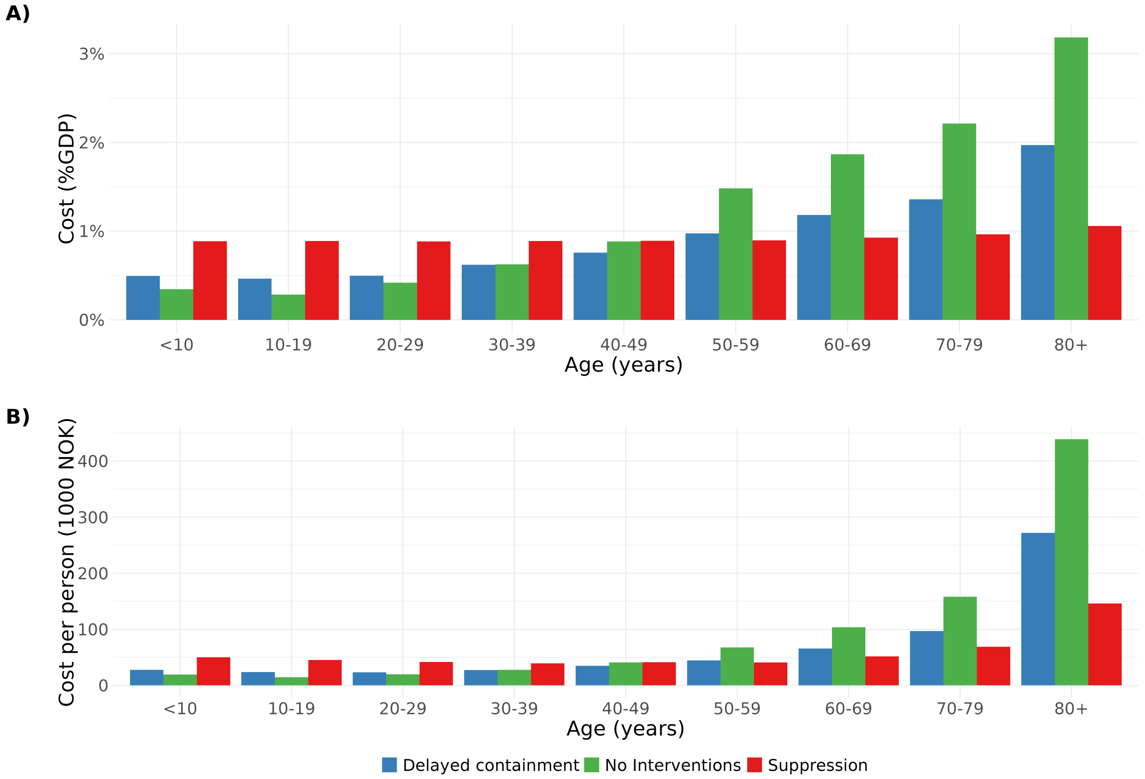

Figure S27: **Distributional effects. Panel A:**Costs per age group and **Panel B:** costs per person in each age group in baseline with an initial reproduction number of 2.4 and a severity of 12. The overall costs of the suppression strategy at 281 bnNOK is lower than the costs of the delayed containment strategy at 283 bnNOK.

#### D.10 Policy scenarios

##### D.10.1 Evaluate policies (Fig 8)

This subsection explains the assumptions in Figure 8 in main paper, relative to the baseline calibration. (For comparison, the solid black line indicates the optimal strategy boundary in the baseline case as displayed in Figure 2.A in main paper). Table S22 presents an overview of how the assumptions are altered in each case, relative to the assumptions in the baseline scenario which is stated in the first row.

| Case: | | Initial cases | Beta | Daily import | Weak vax | ICU cap. | TTIQ | VSD | Time | $COST_Q$ |
|---|---|---|---|---|---|---|---|---|---|---|
| **Figure 2 main paper:** | | | | | | | | | | |
| A-D | Baseline | 900 | – | 1 | No | 350 | Yes | Individual | 9 mo. | 1.4 mnNOK |
| **Figure 8 main paper:** | | | | | | | | | | |
| A-B | Generic reduction | | 10% red | | | | | | | |
| C-D | Border control: Off | | | 210 | | | | | | |
| E-F | Increase ICU capacity | | | | | 450¶ | | | | |
| G-H | Weak vaccine | | | | Yes† | | | | | |
| I-J | Early vaccine 1 | | | | | | | | 100 d. | |
| K-L | Early vaccine 2 | | | | | | | | 200 d. | |

¶ Corresponds to an 29% capacity increase
†In the scenario with an imperfect vaccine we adjust susceptibility and severity for vaccinated individuals as in Table S23.

Table S22: Overview of the different scenarios: Changed assumption compared to baseline

###### Ventilation and face masks

Figures 8.A and 8.B show the effect of a generic reduction in the transmission of the virus of 10%, eg. from increased indoor ventilation at schools or public buildings, or the use of face masks. The left panel shows that this change makes *Suppression* more attractive, implying that it is used for a larger set of $(R, ES)$-combinations. The right panel illustrates why, by displaying how the reduction in cost measured as percent of annual GDP depends on the virus characteristics. Reduced transmission of the virus involves a considerable cost reduction for *Suppression* as it reduces the need for costly interventions, in particular for low to moderate initial reproduction numbers. However, if *No intervention* is optimal, there is little or no gain from a generic reduction in transmission. This illustrates that the gain from specific interventions depends crucially on the chosen strategy, and thus must be evaluated along the chosen policy path.

###### Border control

Figures 8.C and 8.D show the effect of increasing the daily inflow from abroad of infected individuals from 1 in baseline to 210, corresponding to about 40 per million people. This makes *Suppression* less attractive, as stricter interventions are required to keep the effective reproduction number sufficiently below one to offset imported infections. As a result, total costs can rise substantially for virus variants with high severity (right panel). In contrast, when *No intervention* is optimal, additional imports have little effect on total costs, because they have limited effect on cumulative infections once widespread transmission occurs. This illustrates that strict border controls can be valuable as part of a suppression strategy, while the effect is likely to be much smaller without other contact-reducing interventions. This is consistent with findings in the Covid-19 literature [58] that travel restrictions may reduce transmission primarily when combined with measures reducing transmission rates in the community. [58] concludes that if border control measures are to be adopted in future outbreaks, they should be seen as part of a broader strategy that includes other NPIs.

During Covid-19, policies did not always follow this principle. A notable example is the U.S., where federal region-based travel restrictions for non-citizens with recent presence in the Schengen Area, the United Kingdom, Ireland, and Brazil, remained in place until modifications in January 2021 [59], in spite of very high domestic transmissions [60] and with large variation in domestic NPIs across states [61]. Similarly, in England most domestic restrictions were lifted in July 2021, but restrictions on international travels were continued [62].

###### ICU capacity

Figures 8.E and 8.F show the effects of a 29% increase in the ICU capacity, from 350 to 450 beds. Higher ICU capacity reduces the health costs associated with hospital overload, thereby reducing the costs of allowing incidence to rise. This yields large cost reductions for high-transmissibility, high-severity variants for which *Delayed containment* is optimal. In contrast, for variants where *Suppression* is optimal, incidence remains low and ICU capacity is not binding, so the gain is negligible. Thus, increasing ICU capacity shifts the relative attractiveness toward *No intervention* and *Delayed containment*, and away from *Suppression*. Quantifying the gain from increased ICU capacity is useful when evaluating costly investments in hospital capacity.

###### Partially effective vaccine

In Figure 8 panel (G-H) in main paper, we include vaccination with a weak, imperfect and leaky vaccine. The purpose of this scenario is to illustrate the benefits of vaccination when there is a moderate vaccine effect. To implement the scenario we adjust the assumptions about susceptibility and severity for the vaccinated people, these assumptions are listed in Table S23.

| **Type** | **Value** |
|---|---|
| Protection: symp infection | 30% |
| Protection: asymp infection | 20% |
| Protection: hosp | 40% |
| Protection: death | 40% |
| Protection: ICU given hosp | 40% |
| Reduction in hospital stay | 15% |

Table S23: Weak vaccine

Figures 8.G and 8.H show the outcomes under a partially effective vaccine. This reduces health costs substantially for variants with high transmissibility and moderate-to-high severity.

**Vaccine timing**

Figures 8.I and 8.J show outcomes with effective vaccination after 100 days, while Figures 8.K and 8.L show outcomes for vaccination after 200 days. We observe that shorter time to vaccination makes *Suppression* more attractive, because costly interventions are required for a shorter period. For high transmission and high severity, the gains from faster vaccination can be large, up to 7 % of annual GDP in our simulations for $R = 3$ and $ES = 20$ with vaccination at 100 days.

#### D.11 Discussion: Effective vaccine timing

For tractability of the analysis, we have assumed full certainty about the timing of the effective vaccination. However, we conjecture that for the optimal type of strategy, essentially the same results will apply under reasonable assumptions, even if the decision maker only knows the expected time until vaccination, as long as new information does not lead to a change of strategy.

The argument is as follows: We want to find the strategy that minimizes the expected cost $\mathbb{E}[K]$ when the time to an effective vaccine is a stochastic variable, with a probability distribution $p(T)$. We assume that the strategy is chosen at $t = 0$, and there is no further information about when the vaccine arrives which will change the decision for $t > 0$. With constant strategy, the costs of *Suppression* are essentially constant per month, and thus proportional to the duration until vaccination. Thus, the costs are of the form $K_S = sT$, where $s > 0$ are the costs per time unit under *Suppression.* For the *No intervention* and *Delayed containment* strategies, the bulk of the costs of comes during a limited time period with high health costs and for *Delayed containment* also intervention costs, and then small costs per time unit afterwards. Thus, the costs are of the form $K_N = k_N + nT$ where $k_N > 0$ are the health and intervention costs during this period and $n > 0$ are the much lower costs per time unit afterwards.

Under these assumptions, with linear cost functions depending on the time of vaccination $T$, the expected costs are equal to the costs with the expected time of vaccination, $\mathbb{E}[K_S] = s\,\mathbb{E}[T]$ and $\mathbb{E}[K_N] = k_N + n\,\mathbb{E}[T]$ and therefore the optimal strategy will be given by the optimal strategy for $T = \mathbb{E}[T]$.

If there is a possibility that new information may lead to a change of strategy, this may also affect the initial choice of strategy. For example, if there initially is huge uncertainty about the expected time of effective vaccination, it might be optimal to choose *Suppression* for a limited period until there is more information about the expected arrival of an effective vaccine. If there is positive news indicating an early vaccine arrival, *Suppression* can be prolonged, while bad news may lead to a change to *Delayed containment* or *No intervention.*

[5] R Core Team. *R: A Language and Environment for Statistical Computing.* R Foundation for Statistical Computing, Vienna, Austria, 2022.

[6] Rich FitzJohn. odin: ODE Generation and Integration, 2022. R package version 1.4.4.

[7] Lamprini Veneti, Bjarne Robberstad, Anneke Steens, Frode Forland, Brita A. Winje, Didrik F. Vestrheim, Christopher I Jarvis, Amy Gimma, W. John Edmunds, Kevin Van Zandvoort, and Birgitte Freiesleben de Blasio. Social contact patterns during the early COVID-19 pandemic in Norway: insights from a panel study, April to September 2020. *BMC Public Health*, 24:1438, 2024.

[8] Luca Ferretti, Chris Wymant, Michelle Kendall, Lele Zhao, Anel Nurtay, Lucie Abeler-Dörner, Michael Parker, David Bonsall, and Christophe Fraser. Quantifying SARS-CoV-2 transmission suggests epidemic control with digital contact tracing. *Science*, 368, 5 2020.

[9] Jeanette Stålcrantz, Anja Bråthen Kristoffersen, Håkon Bøås, Lamprini Veneti, Elina Seppälä, Nina Aasand, Olav Hungnes, Reidar Kvåle, Karoline Bragstad, Eirik Alnes Buanes, and Robert Whittaker. Milder disease trajectory among COVID-19 patients hospitalised with the SARS-CoV-2 omicron variant compared with the delta variant in Norway. *Scandinavian Journal of Public Health*, 50:676–682, 7 2022.

[10] T. Rowan. Functional stability analysis of numerical algorithms. Dissertation, doctor of philosophy, The University of Texas at Austin, 1990.

[11] Steven G. Johnson. The NLopt nonlinear-optimization package, 2022.

[12] D R Jones, C D Perttunen, and B E Stuckman. Lipschitzian optimization without the Lipschitz constant. *Journal of Optimization Theory and Applications*, 79:157–181, 1993.

[13] M.J.D. Powell. The BOBYQA algorithm for bound constrained optimization without derivatives. Cambridge na report, University of Cambridge, 2009.

[14] Norwegian Institute of Public Health. Risiko ved COVID-19 epidemien og influensaepidemien i Norge. Risk assesment, NIPH, 2022. Accessed: 2022-29-04.

[15] Alfonso Diz-Lois Palomares, Birgitte Freiesleben de Blasio, Louis Yat Hin Chan, Francesco Di Ruscio, and Jørgen E. Midtbø. Combining an agent-based model with gaussian process emulation to model the emergence of the sars-cov-2 omicron variant in norway. *Epidemics*, 54:100895, 2026.

[16] Gro Tunheim, Gunnar Øyvind Isaksson Rø, Trung Tran, Anne-Marte Bakken Kran, Jan Terje Andersen, Eline Benno Vaage, Anette Kolderup, John Torgils Vaage, Fridtjof Lund-Johansen, and Olav Hungnes. Trends in seroprevalence of SARS-CoV-2 and infection fatality rate in the Norwegian population through the first year of the COVID-19 pandemic. *Influenza and Other Respiratory Viruses*, 16:204–212, 3 2022.

[17] Steinar Holden, Thomas von Brasch, Oona Dunlop, Karen Walseth Hara, Ingrid Hjort, Katrine Vellesen Løken, Simen Markussen, Hans Olav Melberg, Anne Spurkland, and Styrk Fjærtoft Vik. Samfunnsøkonomiske vurderinger av smitteverntiltak: Rapport fra ekspertgruppe på oppdrag fra Helse- og omsorgsdepartementet. Ministry of Health and Care Services (Norway), 2022.

[18] Hdir. Virkninger på helse og livskvalitet i utredninger og samfunnsøkonomiske analyser: Temaveileder til utredningsinstruksen. Nasjonal veileder, The Norwegian Directorate of Health, 2024.

[19] DFØ. Verdien av et statistisk liv (vsl) [value of a statistical life], 2024. Accessed: 2023-15-12.

[20] Cristina Menni, Ana M Valdes, Lorenzo Polidori, Michela Antonelli, Satya Penamakuri, Ana Nogal, Panayiotis Louca, Anna May, Jane C Figueiredo, Christina Hu, Erika Molteni, Liane Canas, Marc F Österdahl, Marc Modat, Carole H Sudre, Ben Fox, Alexander Hammers, Jonathan Wolf, Joan Capdevila, Andrew T Chan, Sean P David, Claire J Steves, Sebastien Ourselin, and Tim D Spector. Symptom prevalence, duration, and risk of hospital admission in individuals infected with SARS-CoV-2 during periods of omicron and delta variant dominance: a prospective observational study from the zoe COVID study. *Lancet*, 399:1618–1624, 4 2022.

[28] André Peralta-Santos, Eduardo Freire Rodrigues, Joana Moreno, Vasco Ricoca, Pedro Casaca, Eugenia Fernandes, João Paulo Gomes, Rita Ferreira, Joana Isidro, Miguel Pinto, Vítor Borges, Luís Vieira, Sílvia Duarte, Carlos Sousa, José Pedro Almeida, Luís Menezes, Bibiana I Ferreira, Ana Matias, Ana Pelerito, Samanta Freire, Teresa Grilo, Cláudia Medeiros Borges, Vera Moutinho, Andreia Leite, Irina Kislaya, Ana Paula Rodrigues, Pedro Pinto Leite, and Baltazar Nunes. Omicron (BA.1) SARS-CoV-2 variant is associated with reduced risk of hospitalization and length of stay compared with Delta (B.1.617.2). *medRxiv*, 1 2022.

[29] A Danielle Iuliano, Joan M Brunkard, Tegan K Boehmer, Elisha Peterson, Stacey Adjei, Alison M Binder, Stacy Cobb, Philip Graff, Pauline Hidalgo, Mark J Panaggio, Jeanette J Rainey, Preetika Rao, Karl Soetebier, Susan Wacaster, ChinEn Ai, Vikas Gupta, Noelle-Angelique M Molinari, and Matthew D Ritchey. Trends in disease severity and health care utilization during the early Omicron variants period compared with previous SARS-CoV-2 high transmission periods — U.S. December 2020–January 2022. *MMWR Morb Mortal Wkly Rep*, 71:146–152, 2022. January 25, 2022.

[30] Karin Magnusson, Katrine Damgaard Skyrud, Pål Suren, Margrethe Greve-Isdahl, Ketil Størdal, Doris Tove Kristoffersen, and Kjetil Telle. Healthcare use in 700000 children and adolescents for six months after COVID-19: before and after register based cohort study. *BMJ*, 376:e066809, 2022.

[31] Ismail M Osmanov, Ekaterina Spiridonova, Polina Bobkova, Aysylu Gamirova, Anastasia Shikhaleva, Margarita Andreeva, Oleg Blyuss, Yasmin El-Taravi, Audrey DunnGalvin, Pasquale Comberiati,

Diego G Peroni, Christian Apfelbacher, Jon Genuneit, Lyudmila Mazankova, Alexandra Miroshina, Evgeniya Chistyakova, Elmira Samitova, Svetlana Borzakova, Elena Bondarenko, Anatoliy A Korsunskiy, Irina Konova, Sarah Wulf Hanson, Gail Carson, Louise Sigfrid, Janet T Scott, Matthew Greenhawt, Elizabeth A Whittaker, Elena Garralda, Olivia V Swann, Danilo Buonsenso, Dasha E Nicholls, Frances Simpson, Christina Jones, Malcolm G Semple, John O Warner, Theo Vos, Piero Olliaro, Daniel Munblit, and the Sechenov StopCOVID Research Team. Risk factors for post-COVID-19 condition in previously hospitalised children using the isaric global follow-up protocol: a prospective cohort study. *The European Respiratory Journal*, 59:2101341, 2022.

[39] Francesco Castagna, Xiaonan Xue, Omar Saeed, Rachna Kataria, Yoram A Puius, Snehal R Patel, Mario J Garcia, Andrew D Racine, Daniel B Sims, and Ulrich P Jorde. Hospital bed occupancy rate

is an independent risk factor for COVID-19 inpatient mortality: A pandemic epicentre cohort study. *BMJ open*, 12:e058171, 2 2022.

[40] O. Andersson, P. Campos-Mercade, F. Carlsson, F. Schneider, and E. Wengström. The impact of stay-at-home policies on individual welfare. *The Scandinavian Journal of Economics*, 124:340–362, 2022.

[41] Regjeringen.no. Ekstra overføringer til kommunesektoren under koronapandemien for 2021, 2022. Accessed: 2022-20-12.

[42] Nicola Fuchs-Schündeln. COVID-induced school closures in the US and Germany: Long-term distributional effects. *Economic Policy*, 37:609–639, 2022.

[43] Manudeep Bhuller, Magne Mogstad, and Kjell G Salvanes. Life-cycle earnings, education premiums, and internal rates of return. *Journal of Labor Economics*, 35:993–1030, 4 2017.

[44] Henning Holgersen, Zhiyang Jia, and Simen Svenkerud. Who and how many can work from home? Evidence from task descriptions. *Journal for Labour Market Research*, 55, 2021.

[55] C. Berchet, J. Bijlholt, and M. Ando. Socio-economic and ethnic health inequalities in COVID-19 outcomes across OECD countries. OECD Health WP No. 153, 2023.

[56] Amy Finkelstein, Geoffrey Kocks, Maria Polyakova, and Victoria Udalova. Heterogeneity in damages from a pandemic. Forthcoming. The Review of Economics and Statistics, 2024.

[57] Adam Altmejd, Olof Östergren, Evelina Björkegren, and Torsten Persson. Inequality and COVID-19 in Sweden: Relative risks of nine bad life events, by four social gradients, in pandemic vs. prepandemic years. *Proceedings of the National Academy of Sciences of the United States of America*, 120:e2303640120, 2023.

[58] Karen Ann Grépin, John Aston, and Jacob Burns. Effectiveness of international border control measures during the COVID-19 pandemic: a narrative synthesis of published systematic reviews. *Philosophical Transactions of the Royal Society A: Mathematical, Physical and Engineering Sciences*, 381(2257):20230134, 2023.

[59] United States Executive Office of the President. Proclamation 10138: Terminating suspensions of entry into the united states of aliens who have been physically present in the schengen area, the united kingdom, the republic of ireland, and the federative republic of brazil. U.S. Presidential Proclamation, published in the Federal Register, January 18., 2021.

[60] Centers for Disease Control and Prevention. Covid data tracker weekly review. CDC COVID Data Tracker, February 12., 2021.

[61] Christopher Adolph, Kenya Amano, Bree Bang-Jensen, Nancy Fullman, and John Wilkerson. Pandemic politics: Timing state-level social distancing responses to COVID-19. *Journal of Health Politics, Policy and Law*, 46(2):211–233, 2021.

[62] UK Parliament, House of Commons Library. Travelling abroad under COVID-19 restrictions. Insight. August 19., August 2021.
